# Evidence-guided AI regularization for suicidal ideation prediction in pediatric bipolar disorder

**DOI:** 10.64898/2026.06.18.26355841

**Authors:** Hammza Jabbar Abd Sattar Hamoudi, Mon-Ju Wu, Marsal Sanches, Giovana B. Zunta-Soares, Cesar A. Soutullo, Jair C. Soares, Benson Mwangi

## Abstract

**Background:** Suicide prediction models in psychiatry often rely on purely data-driven feature selection, which can produce unstable and clinically opaque predictor sets in modest-sized samples. We developed Evidence-Based AI LASSO (EBAL), an evidence-guided regularization framework that incorporates curated clinical evidence into feature-specific penalty factors for interpretable prediction.

**Methods:** Baseline data from 136 youth with confirmed bipolar spectrum disorder in the Greater Houston Area Bipolar Registry were analyzed using 20 candidate clinical predictors. Forty higher-level evidence documents on suicidality and related predictor domains were curated through a structured evidence synthesis workflow and indexed as an auditable evidence corpus. An open-weight large language model assigned feature-specific penalty factors using a prespecified scoring rubric, and these penalties were used to fit a weighted LASSO model. EBAL was compared with a standard evidence-agnostic LASSO using nested leave-one-out cross-validation.

**Results:** For suicidal ideation, EBAL achieved an AUROC of 0.768, balanced accuracy of 0.757, sensitivity of 0.758, and specificity of 0.757. The standard LASSO achieved an AUROC of 0.760 and balanced accuracy of 0.715. EBAL improved balanced accuracy (+0.042, p=0.010) and Matthews correlation coefficient (+0.079, p=0.010), while retaining fewer stable predictors than standard LASSO (11/20 vs 18/20). The strongest positive predictors were current depressed mood, duration of mood disorder illness, and comorbid generalized anxiety disorder. For suicidal behavior, both models performed near chance and retained all candidate predictors.

**Limitations:** The study was cross-sectional, single-site, and modest in sample size, with no external validation cohort.

**Conclusions:** EBAL produced a sparser and more clinically coherent model for suicidal ideation in pediatric bipolar disorder, but did not improve prediction of suicidal behavior. These findings support evidence-guided regularization as a transparent strategy for aligning psychiatric prediction models with prior clinical knowledge while preserving interpretability.

## Introduction

Compared with the broader child and adolescent population, youth with bipolar disorder report suicidal ideation at markedly elevated rates. However, clinicians still lack interpretable tools for identifying which patients are most likely to report suicidal thoughts (Hauser et al., 2013; Nock et al., 2013). The risk domains implicated in pediatric bipolar suicidality are clinically familiar and span; depressive symptom severity, mixed mood states, psychiatric comorbidity, family dysfunction, and psychosocial adversity (Axelson et al., 2006; Birmaher et al., 2006; Goldstein et al., 2009; Hauser et al., 2013). Nevertheless, these Individual risk factors have not yielded clinically reliable prediction on their own, a limitation underscored by five decades of suicide-risk research showing that prediction of suicidal thoughts and behaviors remains only marginally better than chance (Franklin et al., 2017). This gap is not simply a problem of insufficient algorithms but it reflects a deeper limitation in model development, where predictions rely on whatever data is available rather than formally integrating prior clinical knowledge.

Machine learning was expected to close this gap by integrating many variables simultaneously, and multivariable approaches have been increasingly applied to suicidality outcomes in psychiatric populations over the past decade (Belsher et al., 2019; Pigoni et al., 2024; Seyedsalehi et al., 2025; Spittal et al., 2025). However, systematic reviews indicate that most suicide prediction models are primarily data-driven, with limited integration of established domain knowledge. Furthermore, many of these models remain vulnerable to overfitting, unstable feature selection, and incomplete reporting (Belsher et al., 2019; Collins et al., 2024; Moons et al., 2025; Mwangi et al., 2014; Pigoni et al., 2024; Seyedsalehi et al., 2025; Spittal et al., 2025; Steyerberg et al., 2010). Feature selection remains a true methodological bottleneck in pediatric psychiatry, acting not as a neutral preprocessing step but rather as the critical juncture where a model either privileges clinically plausible risk domains or locks onto unstable, sample-specific correlations within a modest dataset (Mwangi et al., 2014; Nogueira and Brown, 2016). Among the available strategies for selecting relevant predictors from a larger candidate set, embedded methods are particularly well-suited to clinical prediction because they perform variable selection as part of the model fitting process rather than as a separate preprocessing step (Mwangi et al., 2014). The Least Absolute Shrinkage and Selection Operator (LASSO) is one of the most widely adopted embedded method and achieves sparsity by adding an L1 penalty to the regression objective function, by forcing coefficients of redundant predictors to zero during model estimation (Tibshirani, 1996). This simultaneous feature selection and model estimation process yields interpretable models that clinicians can interpret, a property that is especially valuable in psychiatric risk prediction (Mwangi et al., 2014; Tibshirani, 1996). Therefore, in this study we selected LASSO as it naturally handles the small-sample, large-predictor setting - also known as the “small-n-large-p” problem that characterizes most pediatric psychiatric datasets while producing a sparse model for clinical interpretability and scrutiny. The standard LASSO, however, applies a uniform penalty across all coefficients, treating every candidate predictor as equally likely to be irrelevant before the data are observed (E. Zhang et al., 2025). It offers no mechanism for encoding prior knowledge about which variables carry stronger clinical or empirical support. When predictors are correlated, as they frequently are in psychiatric data, LASSO also tends to retain one variable from a correlated group and discard the rest, which can make selected feature sets unstable across samples (Nogueira and Brown, 2016; Tai and Pan, 2007).

LASSO already supports feature-specific penalty weights, where each predictor can receive a different penalty factor that reflects prior beliefs about its relevance. Indeed, weighted variants such as the adaptive LASSO have demonstrated improved feature selection properties when reliable prior information is available (Tai and Pan, 2007; Zou, 2006). The practical challenge though, is that assigning these weights manually becomes infeasible as the number of candidate predictors grows, and expert opinion alone is difficult to elicit in a systematic, reproducible, and scalable way (E. Zhang et al., 2025). Modern large language models (LLMs) offer a scalable solution to this challenge, but their optimal role in clinical prediction may not be as end-to-end classifiers. Instead, they are better positioned as translators converting unstructured biomedical evidence into the structured regularization weights already supported by the LASSO penalty (Thirunavukarasu et al., 2023). Building on this principle, LLM-LASSO was recently proposed as a framework that uses LLM-derived relevance scores to assign feature-specific penalty factors during model fitting by relaxing the penalty for highly relevant predictors while aggressively penalizing those with weak evidence (E. Zhang et al., 2025). This concept directly addresses the evidence-agnostic limitation of standard LASSO by replacing the uniform penalty with one informed by prior domain knowledge. However, this strategy depends critically on the quality of the text supplied to the model. For example, if the source corpus is broad, noisy, or insufficiently screened, the resulting penalty weights risk encoding literature artifacts rather than clinically valid evidence (E. Zhang et al., 2025).

The solution to this input-quality problem lies in the principles of evidence-based medicine. Therefore, we developed an evidence-guided LLM-LASSO pipeline, termed Evidence-Based AI LASSO (EBAL), for suicidal ideation prediction in pediatric bipolar disorder. Rather than allowing the LLM to draw on unfiltered text, the pipeline begins with a manual and structured evidence synthesis workflow guided by evidence-based practice principles. This framework includes a PICOS-framed research question, systematic database searching, and expert-conducted screening that curates methodologically screened literature relevant to suicidal ideation risk (da Costa Santos et al., 2007; Murad et al., 2016; Page et al., 2021; Sackett et al., 1996). Subsequently, only the resulting curated corpus is supplied to a LLM that translates the synthesized evidence into feature-level relevance weights mapped to candidate variables in the clinical dataset. These weights parameterize a weighted LASSO so that predictors backed by strong prior evidence are less penalized while on the other hand, those lacking empirical support are increasingly penalized (Tai and Pan, 2007; Tibshirani, 1996; E. Zhang et al., 2025). Therefore, the core innovation is that evidence synthesis becomes model regularization and prior clinical evidence is not merely cited after model development as interpretive context but directly incorporated into the penalty structure before estimation. This approach reframes clinical prediction as a knowledge-constrained learning problem. To evaluate the clinical utility of this framework, we tested whether this evidence-guided regularization strategy could produce a sparse, clinically coherent predictor set for suicidal ideation in pediatric bipolar disorder. We further evaluated whether embedding domain knowledge directly into the LASSO penalty structure improved classification performance relative to standard, evidence-agnostic LASSO while preserving interpretability for psychiatric research and clinical translation.

## Methods

### Study design, setting, and participants

This was a cross-sectional analysis of baseline clinical data from the Greater Houston Area Bipolar Registry (GHABR), an ongoing longitudinal cohort study at the Center of Excellence on Mood Disorders - The University of Texas Health Science Center at Houston (UTHealth). The design, procedures, and rationale of the GHABR have previously been described in detail (Diaz et al., 2021). Briefly, the registry enrolls children and adolescents aged 6 to 17 years who meet DSM-5 diagnostic criteria for bipolar disorder type I, type II, or other specified bipolar and related disorder, as well as unaffected offspring of a parent with bipolar disorder, including healthy controls without personal or family psychiatric history. The GHABR participants are recruited from the Harris County Psychiatric Center, the UTHealth child and adolescent outpatient psychiatry clinic, referrals from other healthcare providers, and self-referrals. All psychiatric diagnoses are confirmed using the Mini International Neuropsychiatric Interview for Children and Adolescents, English version 7.0.2 for DSM-5 (MINI-KID) (Sheehan et al., 2010), and enrollment is monitored by a board-certified psychiatrist who reviews each diagnosis obtained through the standardized interview (Diaz et al., 2021). Individuals with a history of autism spectrum disorder, a primary substance use disorder, intellectual disability, severe neurological conditions affecting cognitive status (i.e epilepsy or traumatic brain injury), schizophrenia, or uncontrolled severe medical illness are excluded from the registry. The present analysis was restricted to registry participants with a confirmed bipolar spectrum disorder diagnosis. The analytic sample comprised 136 participants and full demographic and clinical characteristics are summarized in Table 2.

### Outcomes

The primary outcome was suicidal ideation, derived from the MINI-KID suicidality module item B1a. This item assesses intentional self-endangerment and asks whether the respondent planned or expected to hurt themselves on purpose or put themselves in a position where they could be harmed. Responses were coded as a binary variable, with 1 indicating a report of suicidal ideation and 0 indicating no report. This coding was independent of the suicidal behavior outcome, so that any participant who reported ideation on item B1a was counted as positive regardless of behavior status. In the analytic sample, 99 of 136 participants (72.8%) were positive for suicidal ideation and 37 (27.2%) were negative.

As a secondary outcome, we examined suicidal behavior using MINI-KID suicidality module item B1c, which screens for self-harm behavior within the suicidality module’s binary yes/no assessment framework. Responses were coded as a separate binary variable, with 1 indicating a report of suicidal behavior and 0 indicating no report. This coding was independent of the suicidal ideation outcome, so that a participant positive for behavior on item B1c was not necessarily positive for ideation on item B1a. In the analytic sample, 52 of 136 participants (38.2%) were positive for suicidal behavior and 84 (61.8%) were negative.

A descriptive partition of the cohort into mutually exclusive groups of no suicidality, suicidal ideation only, and suicidal behavior is provided in Table 2.

### Candidate predictors

Candidate predictors were derived from the GHABR baseline assessment battery and spanned six clinically relevant domains for pediatric bipolar disorder and suicidality risk assessment. These included current mood state (depressed, euthymic, or manic), illness chronicity (duration of mood disorder), psychiatric comorbidities (generalized anxiety disorder, personality disorder, agoraphobia, and attention-deficit/hyperactivity disorder), current pharmacotherapy (antidepressant and lithium use), service utilization (total hospitalizations and mania-related hospitalization history), physical health indicators (obesity and severe obesity), neurobehavioral measures (parent-reported Behavioral Inhibition System and Behavioral Activation System scales), and sociodemographic and perinatal variables (years of education, paternal education, Hispanic or Latino ethnicity, and birthweight below 5.5 pounds). After matching candidate predictors between the feature specification file and the clinical dataset, 20 variables were retained for modeling. The complete variable dictionary and full candidate predictor list are provided in Supplementary Table S1.

Seven of the 20 predictors contained missing values. The variables with the highest rates of missingness were total number of hospitalizations (15.4%) and duration of mood disorder (14.0%). Parent-reported BIS/BAS scores were each missing in 2.9% of cases, antidepressant use in 2.2%, lithium use in 1.5%, and comorbid personality disorder in 0.7%. Missing values were imputed using the column median prior to modeling. A complete summary of missing data by variable is provided in Supplementary Table S1.

### Evidence synthesis

To construct the domain knowledge corpus used for feature weighting, we performed a systematic evidence synthesis workflow aligned with Cochrane guidance and PRISMA reporting principles (Higgins et al., 2022; Page et al., 2021). In keeping with evidence hierarchies in evidence-based practice, we restricted eligible sources to higher-level evidence documents, specifically systematic reviews, meta-analyses, network meta-analyses, umbrella reviews, and clinical practice guidelines (Murad et al., 2016). A prespecified PICOS framework guided the review question and eligibility structure (da Costa Santos et al., 2007). The primary population of interest was youth with bipolar spectrum disorder. Because higher-level evidence specific to pediatric bipolar suicidality is limited, we also allowed adjacent higher-level evidence sources when they addressed suicidality-relevant predictor domains in youth, mood disorders, bipolar disorder, or clinical practice guidance directly relevant to the candidate predictors. Sources with broader age ranges or broader diagnostic populations were retained only when they mapped to prespecified predictor domains and are flagged in Table 1. The intervention and comparator criteria were not applicable, given that the objective was to construct a domain knowledge corpus for feature weighting rather than to evaluate treatment efficacy or comparative intervention effects. The outcomes of interest were suicidality, including suicidal ideation and suicidal behavior. The study design criterion was restricted to higher-level evidence documents, including systematic reviews, meta-analyses, network meta-analyses, umbrella reviews, and clinical practice guidelines.

**Table 1.**
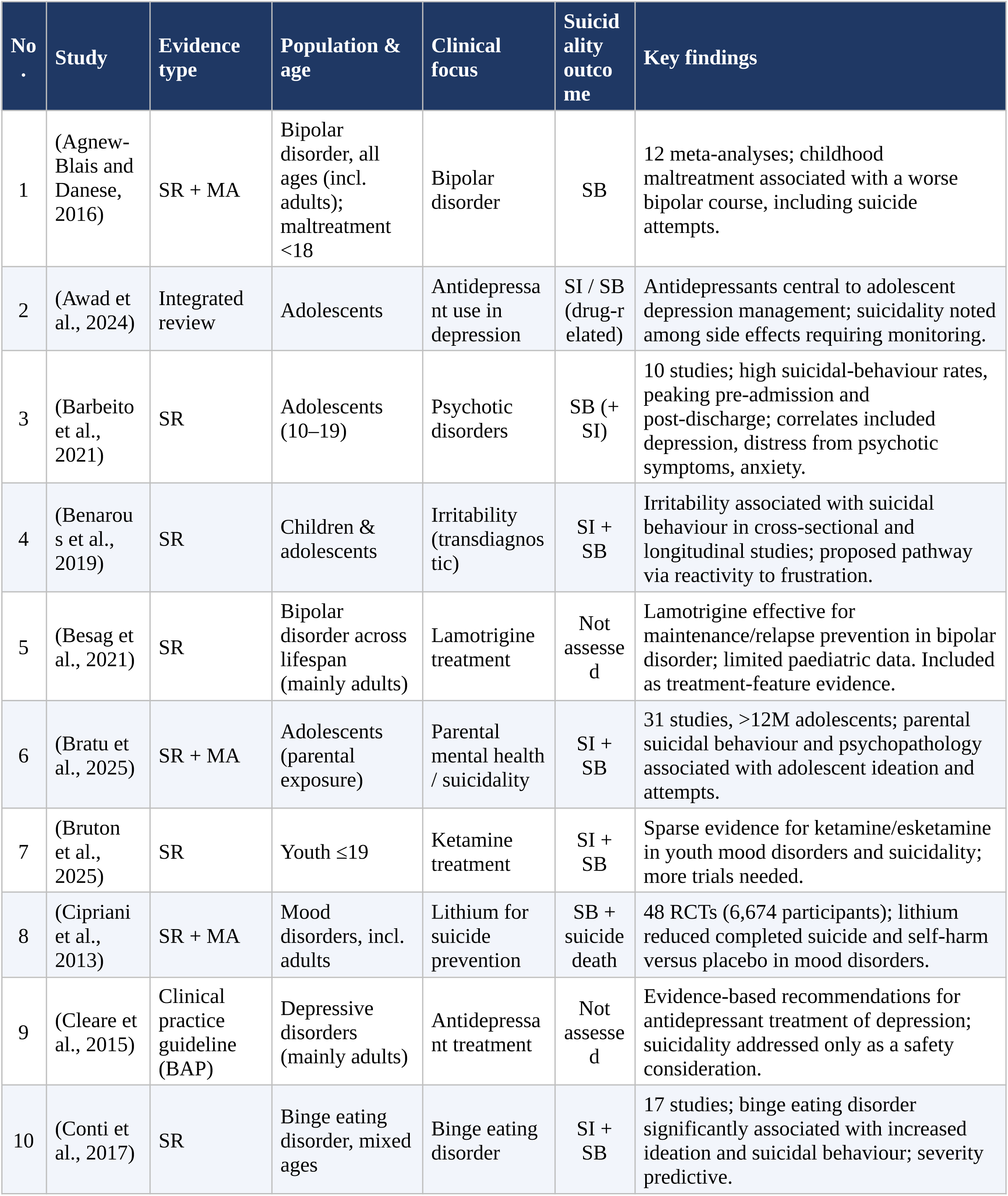
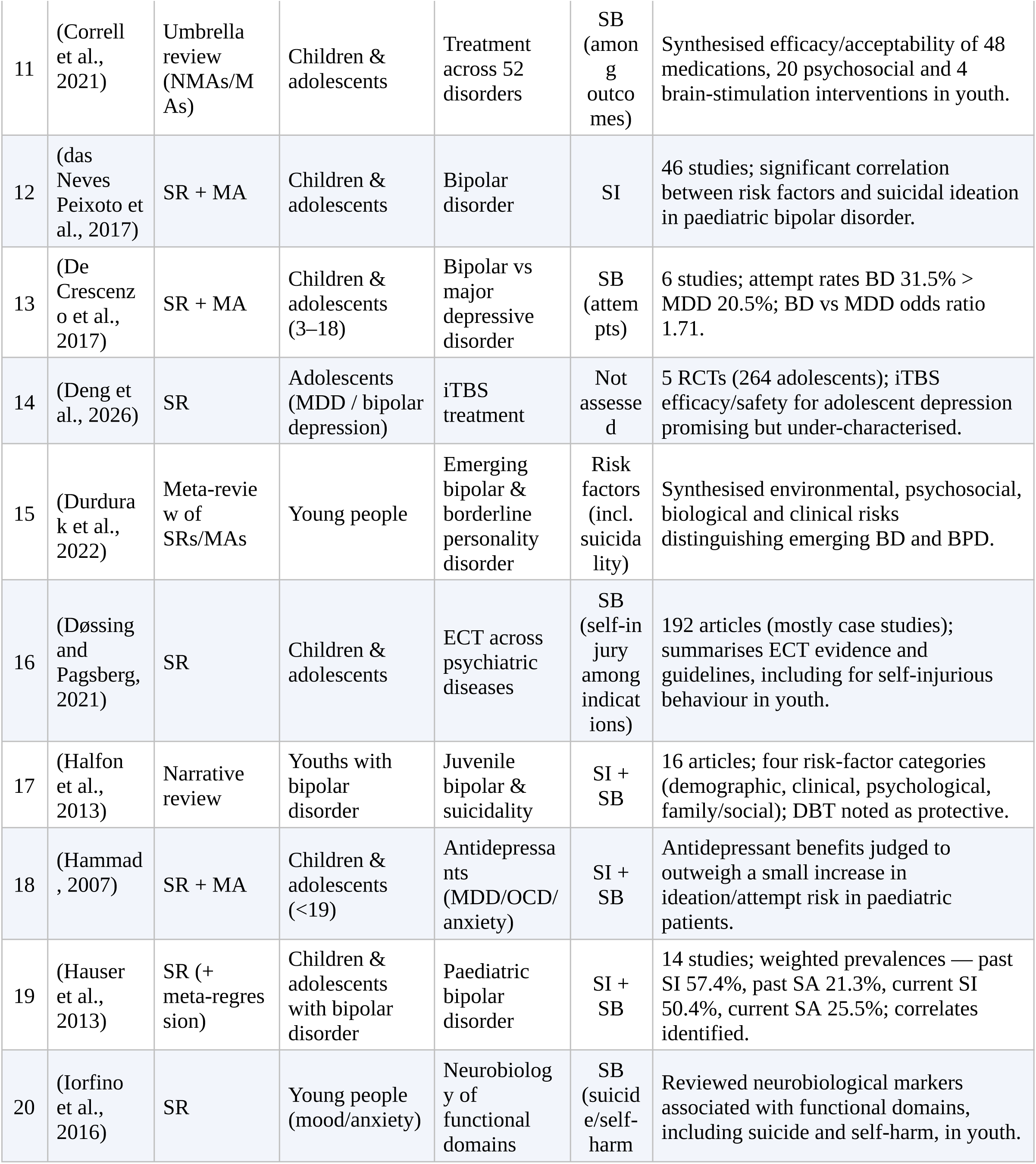
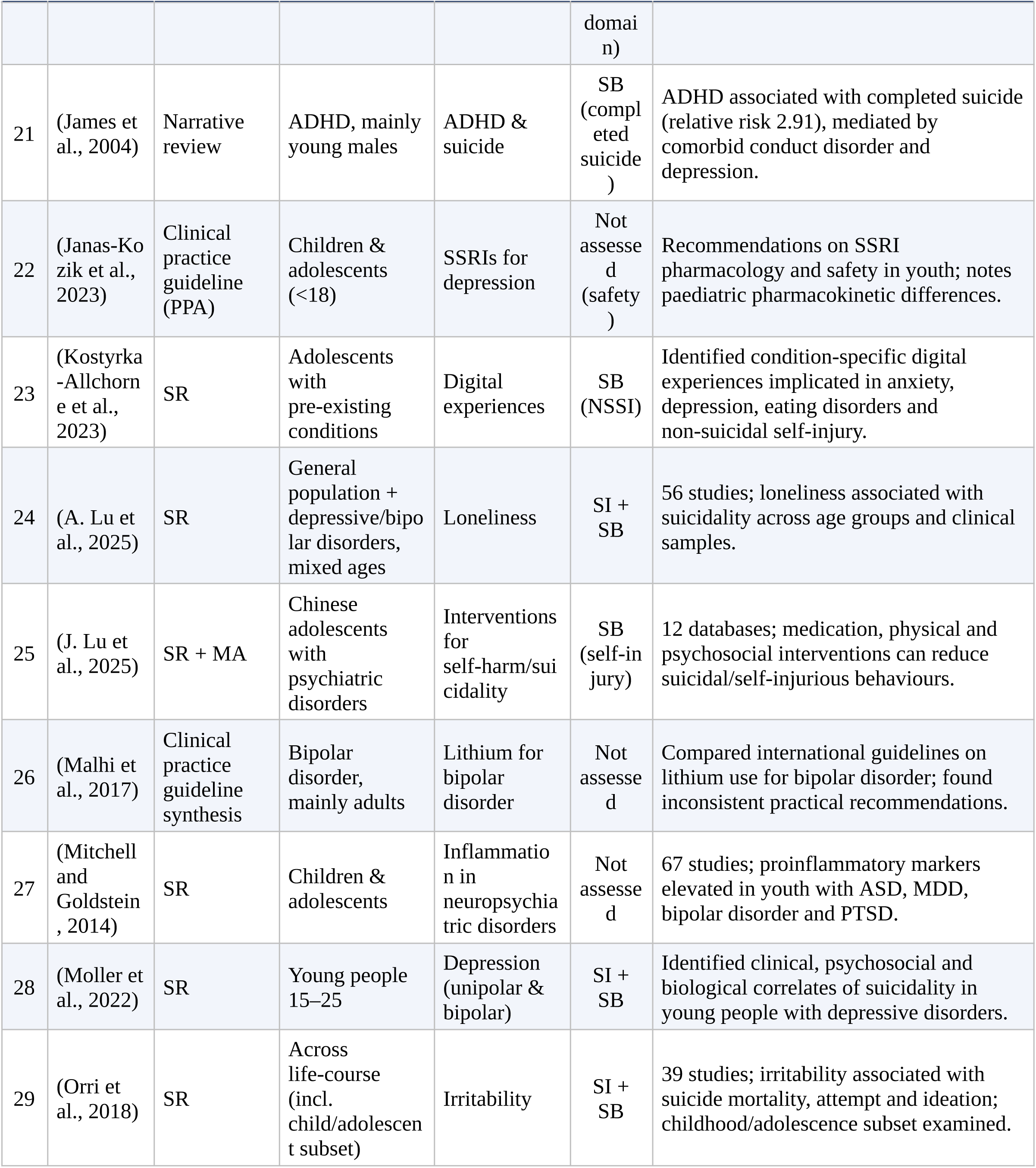
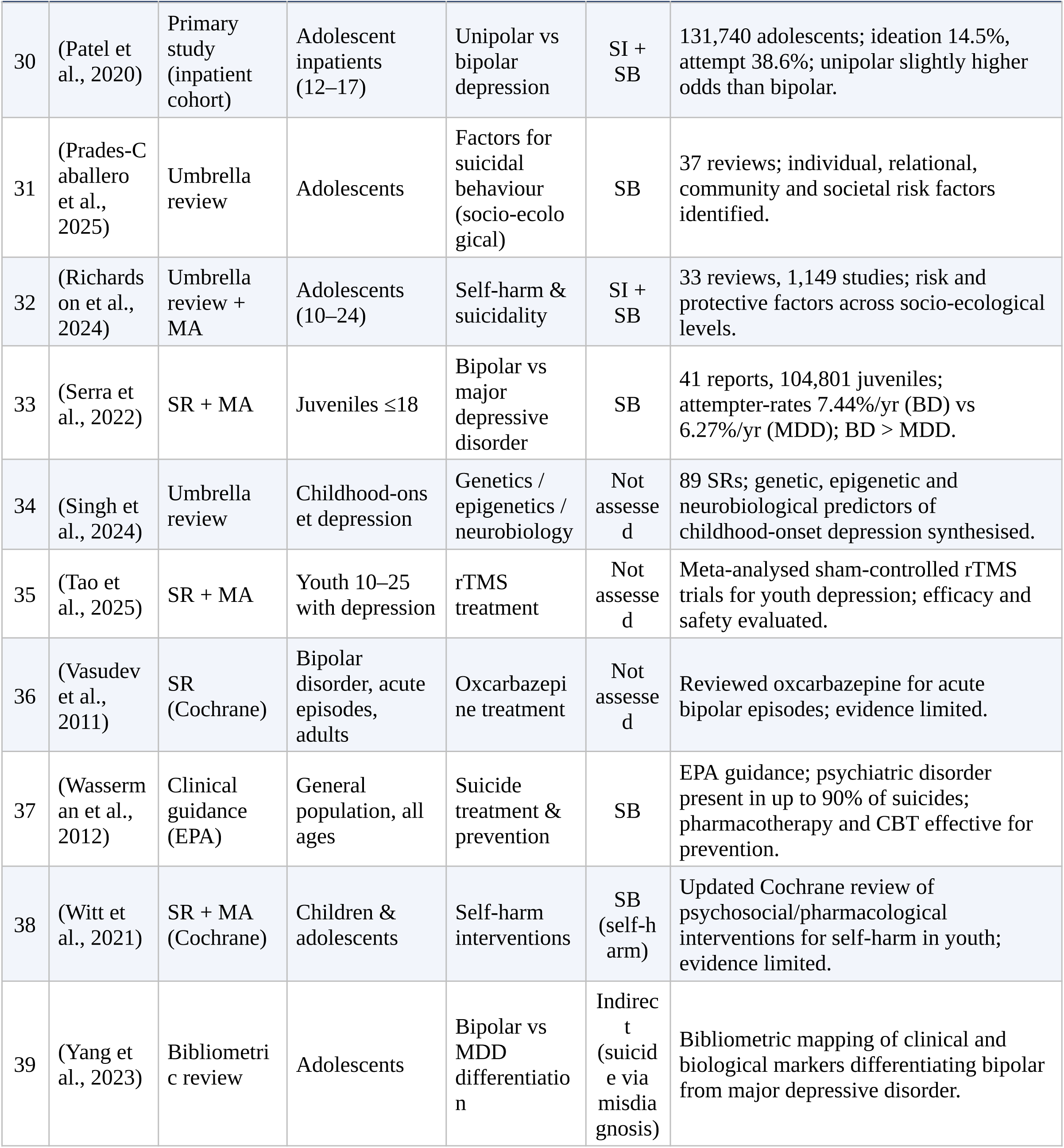
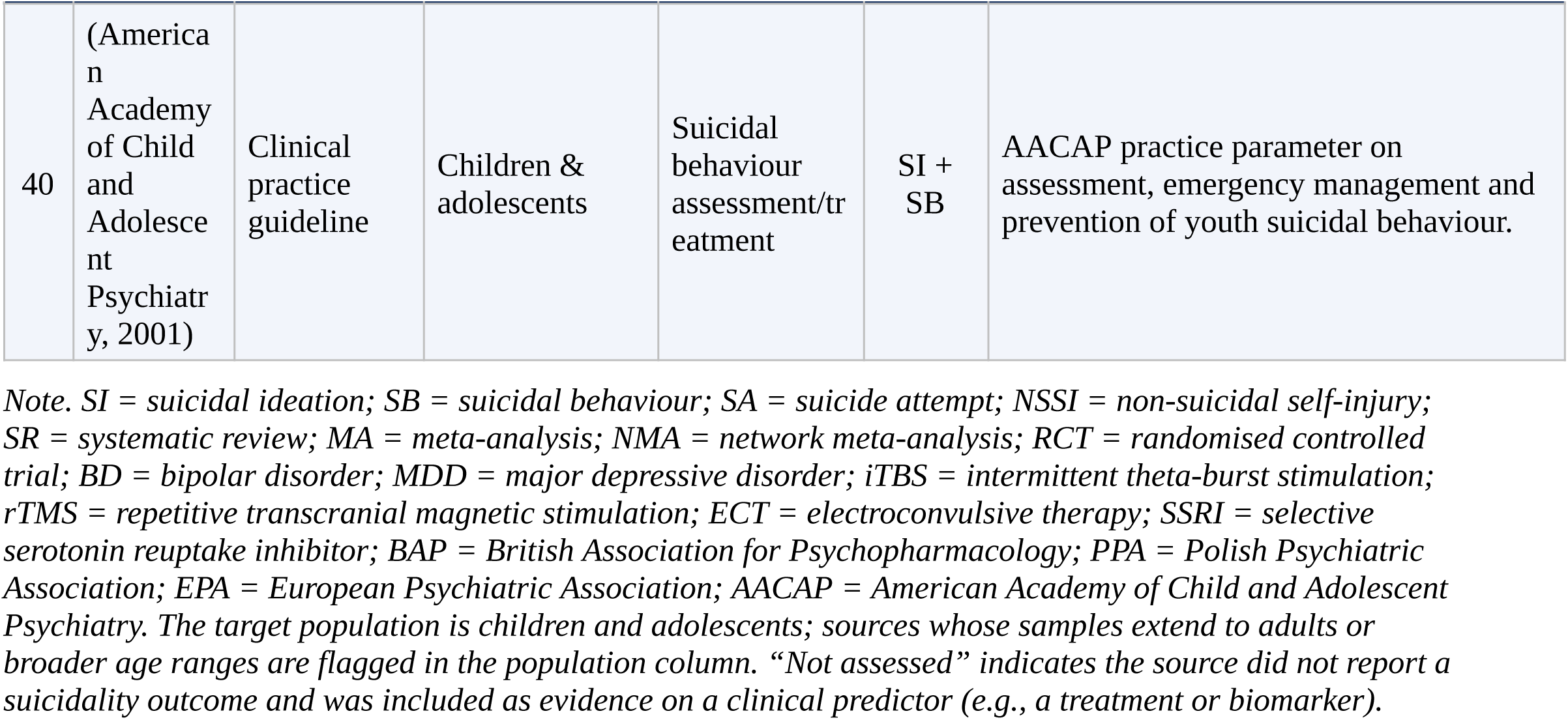
Characteristics of included studies (N = 40).

A systematic search was conducted across PubMed, Embase, PsycINFO, CINAHL, the Cochrane Library, and Web of Science. Records were imported into EndNote for reference management and duplicate removal, followed by additional manual deduplication to ensure accurate record consolidation. The full search strategies for each database, including search dates and database-specific syntax, are provided in Supplementary Methods S1. This evidence synthesis was conducted as a methodological component of the present LLM AI study and was not prospectively registered.

Study selection proceeded in two stages conducted by a single expert reviewer (HJASH). The first stage involved title and abstract screening to identify potentially eligible records. The second stage involved full-text screening to determine final inclusion. Records were included if they met the following prespecified PICOS criteria. First, involved pediatric patients with bipolar spectrum disorders, including bipolar I, bipolar II, or not otherwise specified. Second, evaluated suicidality, suicidal ideation, suicidal behavior, or related risk factors. Third, utilized higher-level or structured evidence designs, including systematic reviews, meta-analyses, network meta-analyses, umbrella reviews, meta-reviews, clinical practice guidelines, or structured evidence-mapping reviews directly relevant to the candidate predictor domains. Records were excluded if they did not address pediatric bipolar disorder, were outside the pediatric age range of interest, did not address suicidality, suicidal ideation, suicidal behavior, or related risk-factor domains, or did not meet the prespecified higher-level evidence study design criterion. Accordingly, primary studies, narrative reviews, scoping reviews, non-systematic literature reviews, commentaries, editorials, letters, case reports, case series, conference abstracts, study protocols, and other study designs not meeting the prespecified high-level evidence criteria were excluded.

Following final inclusion, a total of 40 higher-level evidence documents constituted the domain knowledge corpus used for feature weighting. These documents were then provided to the LLM, with the model-based evidence integration and feature-weighting procedures described in the subsequent sections of the paper (Figure 2). A PRISMA flow diagram summarizing the study selection process is provided in Figure 1. A complete list of included studies are shown in Table 1. In addition a table of excluded full-text studies with reasons for exclusion are provided in Supplementary Tables S2 and S3, respectively.

**Figure 1.**
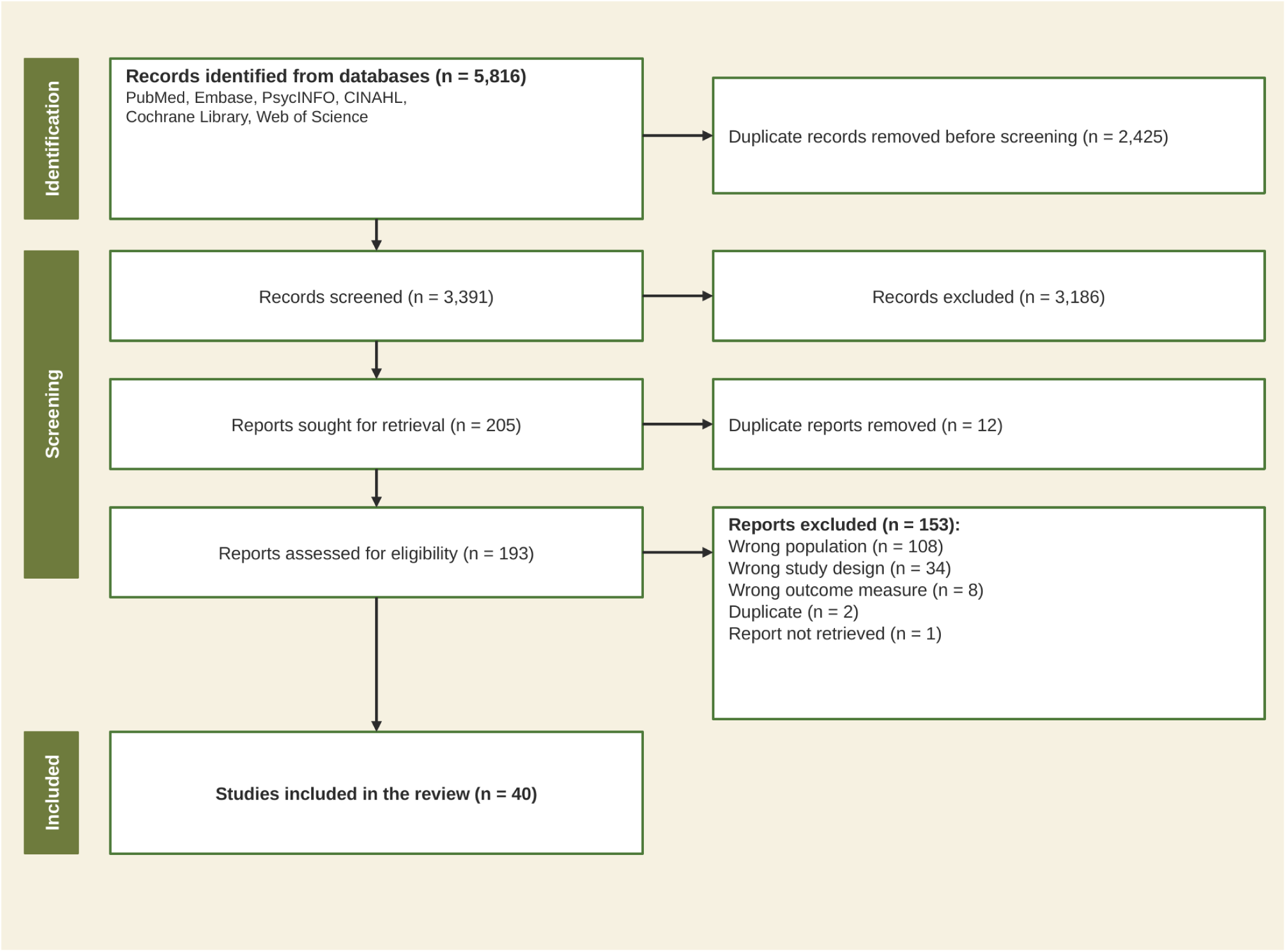
PRISMA flow diagram of the systematic evidence synthesis used to construct the domain knowledge corpus for evidence-guided regularization. Records were identified from six databases (PubMed, Embase, PsycINFO, CINAHL, Cochrane Library, and Web of Science) and screened in two stages by a single expert reviewer (HJASH). Eligible sources were restricted to systematic reviews, meta-analyses, umbrella reviews, and clinical practice guidelines addressing suicidal ideation or suicidal behavior in youth. After duplicate removal and full-text screening, 40 studies were included in the final evidence corpus. These studies served as the knowledge base from which retrieval-augmented generation extracted relevant passages for feature-specific penalty weight assignment. Exclusion reasons at the full-text stage and a complete list of included studies are provided in Supplementary Tables S3 and S2, respectively.

**Figure 2.**
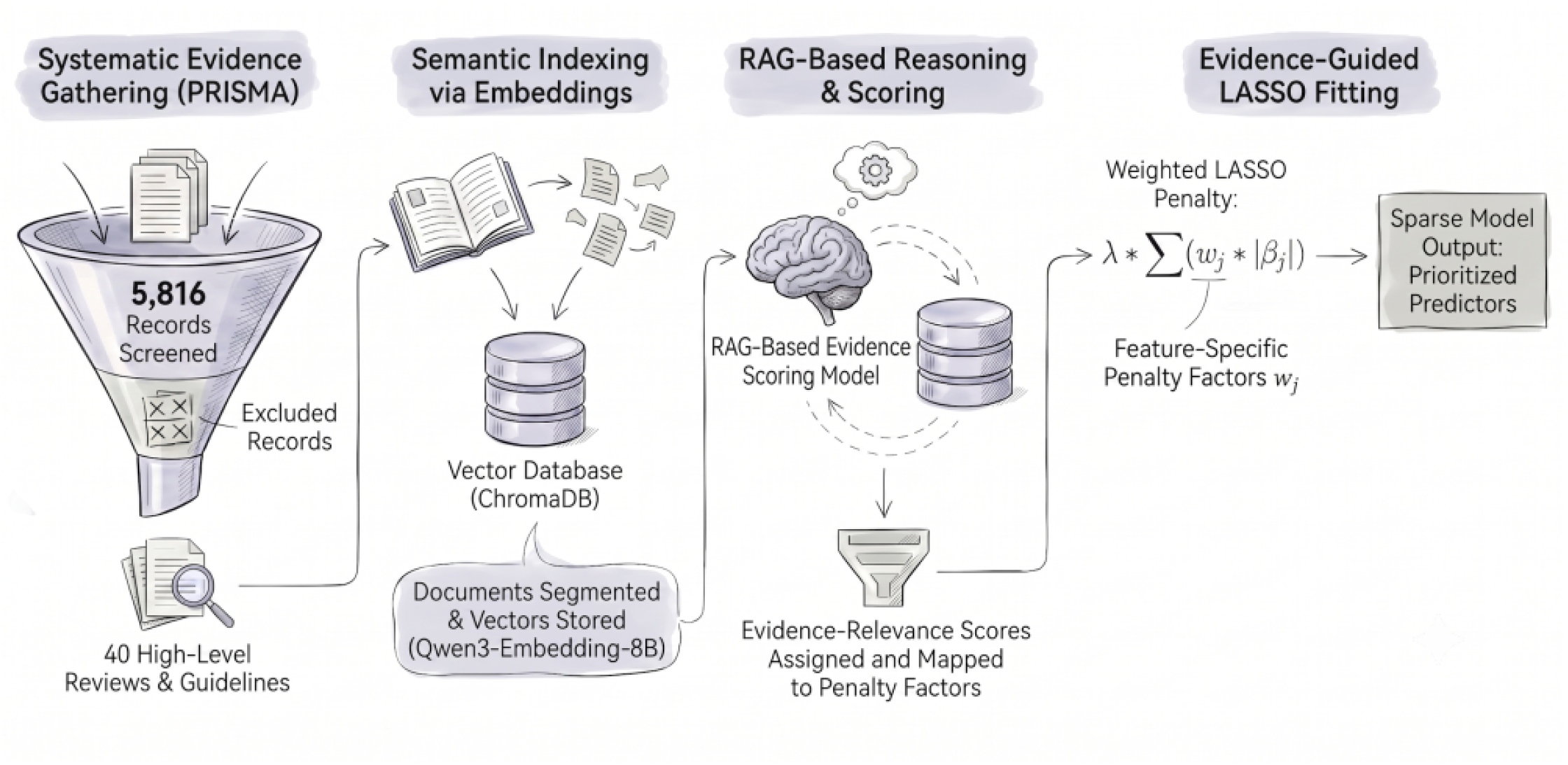
Overview of the Evidence-Based 1 LASSO (EBAL) analytical pipeline. The pipeline translates curated clinical evidence into feature specific penalty weights for regularized regression through four sequential stages. Stage 1 - A systematic search following PRISMA reporting principles screened 5,816 records and identified 40 eligible high-level reviews, meta-analyses, and clinical. practice guidelines. Stage 2 - The included documents were segmented into text chunks, embedded using an open-weight language model (Owen3-Embedding-8B), and stored in a local ChromaDB vector database for semantic retrieval. Stage 3 - A retrieval-augmented generation (RAG) system retrieved the most relevant evidence passages for each candidate predictor and passed them to a reasoning language model (Qwen3-30B-A3B-Thinking), which assigned evidence-relevance cores that were mapped to feature-specific penalty factors Stage 4 - Penalty factors paramertized a weighted LASSO penalty so that predictors supported by stronger clinical evidence received less penalization while those with weaker or absent evidence support were penalized more heavily. The resulting output is a sparse model retaining a prioritized subset of clinically interpretable predisctors.

### Translating evidence into feature-specific penalty weights

A central challenge in implementing evidence-guided regularization is connecting the corpus of curated clinical literature to the numerical penalty weights that the LASSO model requires. To bridge this gap, we used a technique known as retrieval-augmented generation (RAG), in which a LLM is given relevant passages retrieved from the synthesized knowledge base before it makes a judgment, rather than relying solely on its own pretrained knowledge (Lewis et al., 2020). In our pipeline, this meant that when the language model was asked to evaluate the relevance of a candidate predictor to suicidal ideation, it first received the most pertinent segments from the 40 screened evidence documents as context before assigning a penalty score.

Two open-weight language models from the Qwen3 family were used, both served through the vLLM inference framework (v0.12.0) (Kwon, 2025). The first was Qwen3-Embedding-8B (Y. Zhang et al., 2025), an 8-billion-parameter text-embedding model that converts text into high-dimensional vector representations suitable for similarity-based retrieval. We selected this model because it provides up to 4,096-dimensional embeddings, supports flexible output dimensionality through Matryoshka representation learning, and achieved the strongest reported performance among the Qwen3 embedding family on multilingual embedding benchmarks (Li et al., 2026). This larger, less-compressed representation was preferred for distinguishing closely related clinical concepts, where semantic similarity tasks are known to be challenging because clinical language often contains specialized terminology and abbreviations. This is important when the retrieval system must differentiate between closely related psychiatric constructs such as depressive symptom burden and general mood state. The second model was Qwen3-30B-A3B-Thinking (Xu et al., 2025), a mixture-of-experts reasoning model with approximately 30.5 billion total parameters and 3.3 billion activated parameters per forward pass (Mwangi et al., 2026). We selected this model for penalty scoring because it’s thinking-mode generation is designed for complex, multi-step reasoning rather than rapid single-pass response generation. In this study, penalty scoring required the model to evaluate the relationship among the candidate predictors, the retrieved clinical evidence, and the target outcome, then assign an evidence-relevance weight. This task is better framed as structured clinical relevance judgment as opposed to a simple lexical or embedding similarity matching. Notably, both models were also selected for their open-weight availability, which ensures that the exact model weights and configurations used in this study can be reproduced by other groups without dependence on proprietary cloud services. Both models were deployed on local institutional GPU hardware using containerized vLLM (v0.12.0) inference servers. The reasoning LLM (Qwen3-30B-A3B-Thinking) was served on an NVIDIA Blackwell RTX GPU with 96 GB of video memory, and the embedding model (Qwen3-Embedding-8B) was served on a separate NVIDIA ADA GPU with 48 GB of video memory. No clinical or evidence data left our institutional computing environment at any stage of the analysis.

To build the evidence knowledge base, the 40 included documents were first preprocessed by splitting the text into segments of approximately 1,000 characters with a 200-character overlap. These segments were subsequently converted into high-dimensional vector representations using the Qwen3 embedding model before being indexed into a local ChromaDB vector database (Boppana et al., 2024). Reference and bibliography pages were automatically identified and filtered to prevent citation metadata from contaminating the retrieval results. This process produced 4,246 indexed text segments drawn from 878 pages of source material, with 134 pages (15.3%) excluded as reference content. The full indexing process of the 40 documents completed in approximately 24 minutes. At the penalty scoring stage, the pipeline issued a structured query for each of the 20 candidate predictors, asking the retrieval system to return three text segments most semantically relevant to that predictor in the context of suicidal ideation. Across all 20 predictors, 20 retrieval queries were issued, yielding 60 retrieved evidence segments, and 20 of the 40 source documents contributed at least one retrieved segment. The retrieved evidence corpus was used to define and audit the feature-weighting context, but the final penalty assignment was constrained to a name-grounded rubric to prevent uncontrolled clinical inference by the language model, which was instructed through a structured prompt to assign an integer penalty factor on a prespecified scale from 2 to 5. A score of 2 indicated that the predictor name explicitly referenced suicidal ideation or self-harm and was supported by the retrieved evidence. A score of 3 indicated that the predictor referenced a severe mood or affective construct within the diagnostic scope. A score of 4 indicated a broader clinical or psychosocial construct such as anxiety or attention-deficit/hyperactivity disorder. A score of 5 indicated that the predictor was demographic, administrative, or not interpretable as directly relevant to suicidal ideation based on the available evidence. The model operated at a temperature of 0.1 to produce deterministic outputs, and all 20 predictors were scored in a single pass. Total inference time for penalty scoring across all 20 predictors was approximately 70 seconds. The complete reasoning trace produced by the language model during penalty scoring, including the chain-of-thought evaluation for each of the 20 predictors, is provided in Supplementary Methods S3. Crucially, the LLM received only the predictor name and the retrieved evidence segments, with no access to clinical outcome data, ensuring complete separation between the evidence-derived weights and the analytical dataset (Figure 2).

### Model fitting and evaluation

Given the binary outcome, both the EBAL and the standard LASSO comparator were fit as penalized logistic regression models with pure L1 regularization (Tibshirani, 1996; E. Zhang et al., 2025). All candidate predictors were normalized by z-scoring prior to model fitting, with the scaling parameters (mean and standard deviation) computed from the training data and applied identically to the test observation in each evaluation fold. To address the class imbalance problem where the primary outcome (suicidal ideation) exhibited a class imbalance of approximately 2.7 to 1, we applied the Synthetic Minority Oversampling Technique (SMOTE) method (Chawla et al., 2002). SMOTE was applied exclusively to training data and never to the held-out test observations, ensuring that performance estimates were not contaminated by synthetic samples.

Model performance was evaluated using a nested leave-one-out cross-validation (LOOCV) design. In the outer loop, each of the 136 participants was held out once as a single-observation test set, producing one predicted probability per participant across 136 iterations. Within each outer fold, an inner 10-fold cross-validation was used to select the optimal regularization strength (lambda) from a path of 100 candidate values. The inner loop was optimizing for balanced classification accuracy. After the best lambda was identified, the model was refitted on the full SMOTE-augmented training set at that lambda value, and the held-out participant’s probability was predicted. Coefficients from each fold were also collected for stability analysis. The LLM-derived penalty weights were computed once before the cross-validation loop and held constant across all 136 folds, so that the evidence-to-penalty translation was independent of the evaluation procedure. Predicted probabilities were converted to binary classifications using a 0.5 probability threshold.

After all 136 LOO predictions were collected, we computed discrimination and classification metrics including the area under the receiver operating characteristic curve (AUROC), accuracy, balanced accuracy, sensitivity, specificity, precision, F1 score, and Matthews correlation coefficient (MCC). Ninety-five percent confidence intervals for all performance metrics were generated using a standard percentile bootstrap with 1,000 resamples (Tibshirani and Efron, 1993). A summary of bootstrap confidence interval widths for all metrics is provided in Supplementary Figure S2. To obtain stable coefficient estimates for reporting, a final model was fit on all 136 participants using the same SMOTE augmentation and inner cross-validation procedure. Feature selection stability was assessed by computing the proportion of LOO folds in which each predictor received a non-zero coefficient, with features selected in more than 50% of folds considered stable predictors (Figure 5B).

### Standard LASSO as a comparator model

To evaluate the incremental value of evidence-informed weighting, we fit a standard LASSO model using the same 20 candidate predictors but with uniform penalty factors in which all variables were penalized equally (Tibshirani, 1996). This comparator used an identical evaluation pipeline, including the same leave-one-out cross-validation structure, SMOTE oversampling, inner cross-validation for regularization parameter selection, and lambda path, so that the only difference between the two models was the penalty vector. This design ensures that any observed difference in performance or sparsity can be attributed to the evidence-guided penalty weighting rather than to differences in model fitting or evaluation procedures.

### Software and reproducibility

All analyses were implemented using the Python programming language. The complete EBAL pipeline code, including the evidence-retrieval system, penalty scoring, model fitting, and evaluation scripts, is publicly available at https://github.com/bmwas/LLM-Lasso. This repository is a modified and extended version of the original LLM-LASSO codebase (E. Zhang et al., 2025), adapted to support evidence-based corpus curation, local open-weight model deployment, and the clinical prediction workflow described in this study. Penalized logistic regression with feature-specific penalty factors was fit using the Adelie library (E. Zhang et al., 2025). Cross-validation utilities relied on scikit-learn (Pedregosa et al., 2011), and class imbalance correction used the SMOTE implementation in imbalanced-learn (Chawla et al., 2002). The retrieval-augmented generation pipeline was orchestrated through LangChain, with ChromaDB serving as the local vector database for evidence storage and retrieval. Both language models used in this study (Qwen3-30B-A3B-Thinking and Qwen3-Embedding-8B) are open-weight models with publicly available model files hosted on HuggingFace and were served locally through containerized vLLM (v0.12.0) Docker inference servers. The complete prompting templates, scoring instructions, and penalty normalization procedures are provided in Supplementary Methods S2.

## Results

### Participant Demographics

A total of 136 participants with pediatric bipolar spectrum disorder were included in the analysis (Table 2). Mean age was 13.2 years (SD 3.1, range 6–17), and 70 participants (51.5%) were female. The primary outcome, MINI-KID suicidality item B1a, was reported by 99 of the 136 participants (72.8%). Outcome data were complete with no missing values for B1a.

**Table 2.**
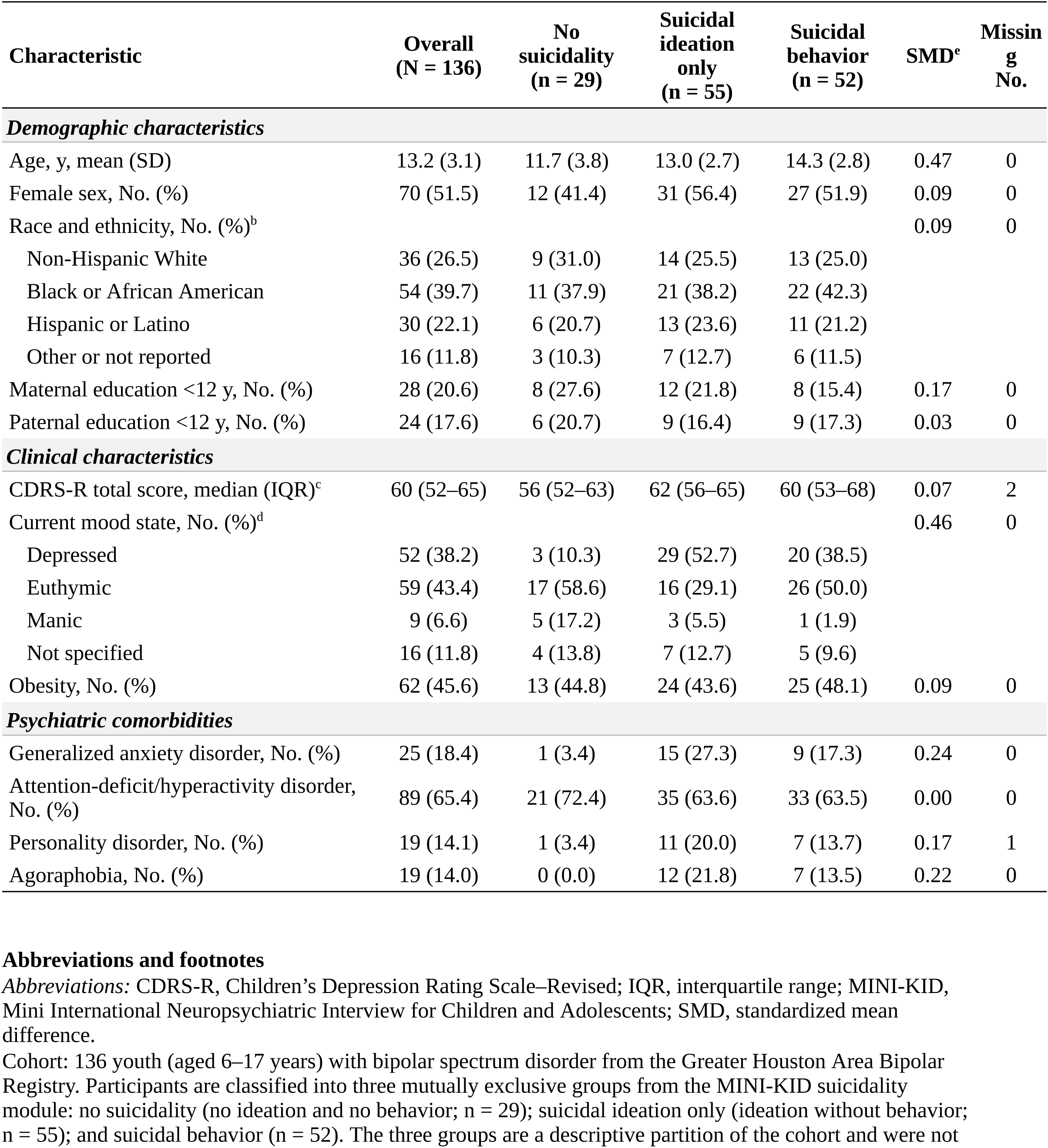
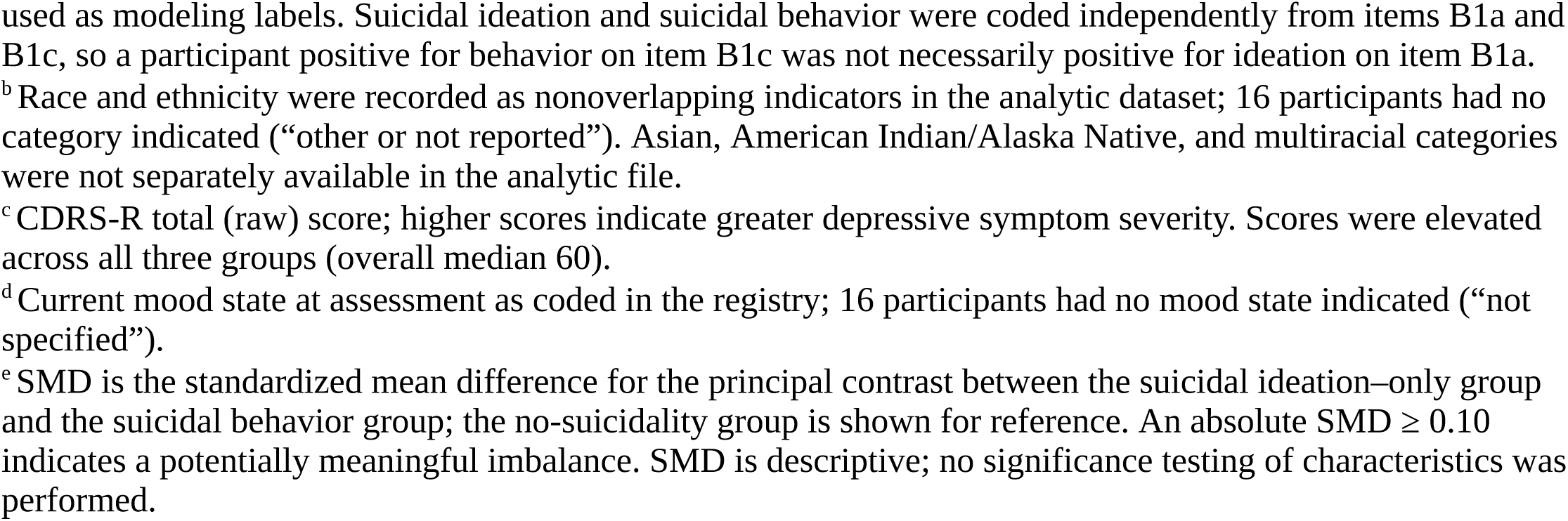
Baseline Demographic and Clinical Characteristics of the Pediatric Bipolar Cohort, Overall and by Suicidality Group (No Suicidality, Suicidal Ideation Only, and Suicidal Behavior)

| Characteristic | Overall<br>(N = 136) | No<br>suicidality<br>(n = 29) | Suicidal<br>ideation<br>only<br>(n = 55) | Suicidal<br>behavior<br>(n = 52) | SMD<sup>e</sup> | Missing<br>No. |
| --- | --- | --- | --- | --- | --- | --- |
| **Demographic characteristics** |  |  |  |  |  |  |
| Age, y, mean (SD) | 13.2 (3.1) | 11.7 (3.8) | 13.0 (2.7) | 14.3 (2.8) | 0.47 | 0 |
| Female sex, No. (%) | 70 (51.5) | 12 (41.4) | 31 (56.4) | 27 (51.9) | 0.09 | 0 |
| Race and ethnicity, No. (%)<sup>b</sup> |  |  |  |  | 0.09 | 0 |
| Non-Hispanic White | 36 (26.5) | 9 (31.0) | 14 (25.5) | 13 (25.0) |  |  |
| Black or African American | 54 (39.7) | 11 (37.9) | 21 (38.2) | 22 (42.3) |  |  |
| Hispanic or Latino | 30 (22.1) | 6 (20.7) | 13 (23.6) | 11 (21.2) |  |  |
| Other or not reported | 16 (11.8) | 3 (10.3) | 7 (12.7) | 6 (11.5) |  |  |
| Maternal education <12 y, No. (%) | 28 (20.6) | 8 (27.6) | 12 (21.8) | 8 (15.4) | 0.17 | 0 |
| Paternal education <12 y, No. (%) | 24 (17.6) | 6 (20.7) | 9 (16.4) | 9 (17.3) | 0.03 | 0 |
| **Clinical characteristics** |  |  |  |  |  |  |
| CDRS-R total score, median (IQR)<sup>c</sup> | 60 (52–65) | 56 (52–63) | 62 (56–65) | 60 (53–68) | 0.07 | 2 |
| Current mood state, No. (%)<sup>d</sup> |  |  |  |  | 0.46 | 0 |
| Depressed | 52 (38.2) | 3 (10.3) | 29 (52.7) | 20 (38.5) |  |  |
| Euthymic | 59 (43.4) | 17 (58.6) | 16 (29.1) | 26 (50.0) |  |  |
| Manic | 9 (6.6) | 5 (17.2) | 3 (5.5) | 1 (1.9) |  |  |
| Not specified | 16 (11.8) | 4 (13.8) | 7 (12.7) | 5 (9.6) |  |  |
| Obesity, No. (%) | 62 (45.6) | 13 (44.8) | 24 (43.6) | 25 (48.1) | 0.09 | 0 |
| **Psychiatric comorbidities** |  |  |  |  |  |  |
| Generalized anxiety disorder, No. (%) | 25 (18.4) | 1 (3.4) | 15 (27.3) | 9 (17.3) | 0.24 | 0 |
| Attention-deficit/hyperactivity disorder, No. (%) | 89 (65.4) | 21 (72.4) | 35 (63.6) | 33 (63.5) | 0.00 | 0 |
| Personality disorder, No. (%) | 19 (14.1) | 1 (3.4) | 11 (20.0) | 7 (13.7) | 0.17 | 1 |
| Agoraphobia, No. (%) | 19 (14.0) | 0 (0.0) | 12 (21.8) | 7 (13.5) | 0.22 | 0 |

### Evidence Synthesis Corpus

The systematic search identified 5,816 records, of which 3,391 unique records remained after duplicate removal. Following title and abstract screening, 205 reports were sought for retrieval. After removal of 12 further duplicate reports, 193 reports were assessed for full-text eligibility, of which 153 were excluded, leaving

40 studies that met the predefined inclusion criteria and were included in the final evidence corpus. The resulting EBAL analytical workflow, from evidence synthesis through retrieval, penalty scoring, and weighted LASSO fitting, is summarized in Figure 2. The study selection process is summarized in the PRISMA flow diagram (Figure 1). A complete list of included studies is provided in Table 1, and excluded full-text studies with reasons for exclusion are provided in Supplementary Table S3. The distribution of retrieved evidence across the highest-contributing studies and candidate predictors is shown in Figure 3.

**Figure 3.**
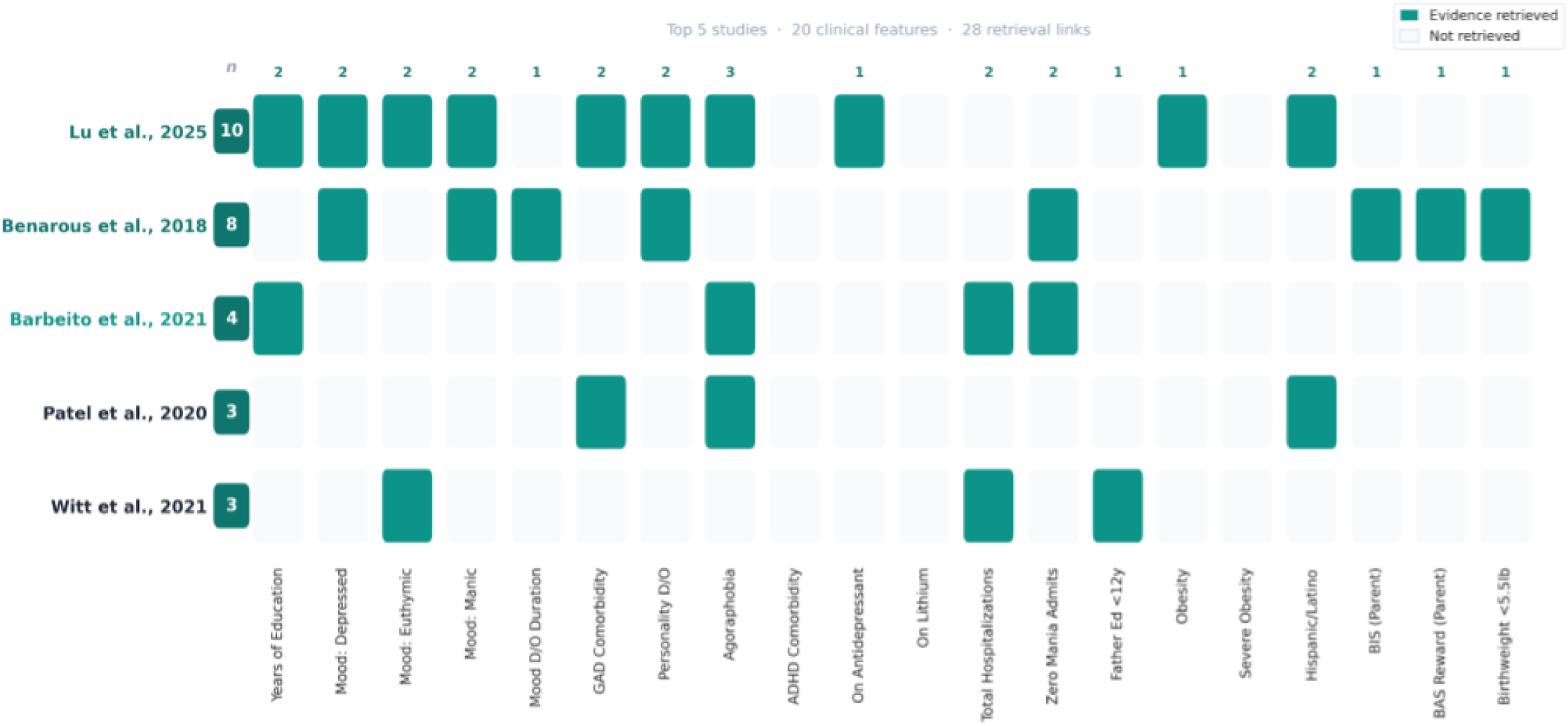
Evidence retrieval distribution across the five highest-contributing studies in tile curated evidence corpus. Rows represent the five studies that contributed evidence to the greatest number of candidate predictors, ranked by total retrieval count (n, shown at left). Columns correspond to the 20 candidate clinical predictors evaluated in the penalty scoring stage. Filled cells indicate that the retrieval-augmentcd generation system retrieved at least one text segment from that study when querying for the corresponding predictor, and empty cells indicate no segment was retrieved. The number above each column indicates how many of these five studies contributed evidence for that predictor. Of the 40 studies in the full evidence corpus, 20 contributed at least one retrieved segment across all predictor queries, and the five studies shown here accounted for 28 of the total retrieval links. Lu et al. 2025 provided the broadest coverage, contributing evidence to 10 of the 20 predictors, followed by Benarous et al.2019 with evidence spanning 8 predictors.

#### Evidence-Based AI LASSO

Using LOOCV with a prespecified probability threshold of 0.5, the EBAL achieved an area under the receiver operating characteristic curve (AUROC) of 0.768 (95% CI 0.672 to 0.857). Balanced accuracy was 0.757 (95% CI 0.676 to 0.837), with sensitivity of 0.758 (95% CI 0.670 to 0.842) and specificity of 0.757 (95% CI 0.610 to 0.900). Positive predictive value was 0.893 (95% CI 0.819 to 0.959) and negative predictive value was 0.538 (95% CI 0.400 to 0.673). The final model retained 11 of 20 candidate predictors (see Figure 5B and Supplementary Data S3). The three strongest positive predictors were; current depressed mood, duration of mood disorder illness, and comorbid generalized anxiety disorder. Feature selection was stable, with the top seven predictors selected in 100% of the 136 cross-validation folds. The retained coefficient profile is shown in Figure 4B, and selection-frequency stability across leave-one-out folds is shown in Figure 5B. Additional performance metrics are provided in the supplementary materials.

**Figure 4.**
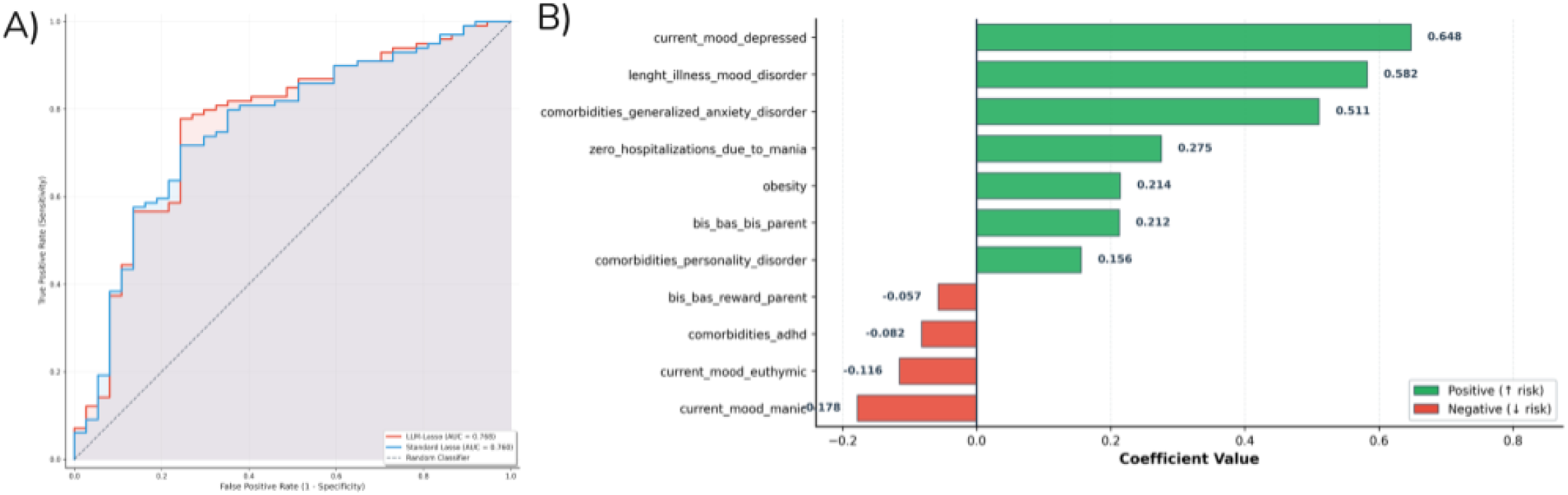
Classification performance and retained predictors of Evidence-Based AI LASSO for suicidal ideation (MINI-KID item B1a, N=136). (A) Receiver operating characteristic curves comparing Evidence-Based AI LASSO (AUROC = 0.768) and standard LASSO with uniform penalties (AUROC = 0.760), evaluated using leave-one-out cross-validation. The dashed line represents chance-level classification. (B) Standardized coefficients from the final Evidence-Based AI LASSO model, which retained 11 of 20 candidate predictors. Green bars indicate positive associations with suicidal ideation (increased risk) and red bars indicate inverse associations (decreased risk). The seven-positive predictors were current depressed mood, duration of mood disorder illness, comorbid generalized anxiety disorder, no prior mania-related hospitalizations, obesity, parent-reported Behavioral Inhibition System (BIS) score, and comorbid personality disorder. The four inverse predictors were current manic mood, current euthymic mood, comorbid attention-deficit/hypernctivity disorder, and parent-reported Behavioral Activation System (BAS) Reward score. Nine candidate predictors received zero coefficients and were eliminated from the model.

#### Standard LASSO comparator

Under the same evaluation framework, the standard LASSO achieved an AUROC of 0.760 (95% CI 0.664 to 0.848), balanced accuracy of 0.715 (95% CI 0.627 to 0.803), sensitivity of 0.727 (95% CI 0.644 to 0.814), and specificity of 0.703 (95% CI 0.545 to 0.861). Positive predictive value was 0.868 (95% CI 0.792 to 0.935) and negative predictive value was 0.491 (95% CI 0.357 to 0.628). The standard LASSO retained 18 of 20 candidate predictors, indicating substantially less sparsity than our proposed method - EBAL.

#### Head-to-head comparison

EBAL outperformed the standard LASSO across all primary evaluation metrics for suicidal ideation (Figure 4), while retaining only 11 predictors compared with 18 for the standard LASSO, a 39% reduction in model complexity. Post-hoc paired statistical tests were conducted on the 136 leave-one-out predictions to formally evaluate these differences. McNemar’s exact test identified five discordant participant-level predictions between the two models, all five of which favored EBAL with none favoring the standard LASSO (exact two-sided p = 0.063, mid-p = 0.047). A DeLong test comparing the AUROCs of the two models was not significant (z = 1.01, p = 0.313), consistent with the small absolute difference in discrimination (+0.008). However, paired bootstrap comparisons with 10,000 iterations confirmed that EBAL’s advantages over standard LASSO in balanced accuracy (+0.042, p = 0.010) and Matthews correlation coefficient (+0.079, p = 0.010) were each statistically significant. The overall pattern indicates a consistent and statistically supported advantage for EBAL in classification performance for suicidal ideation, despite the AUROC difference alone not reaching significance.

### Suicidal behavior (MINI-KID item B1c, secondary outcome)

#### Evidence-Based AI LASSO

For the secondary outcome of suicidal behavior (52 positive, 84 negative), the EBAL achieved an AUROC of 0.594 (95% CI 0.490 to 0.689). Balanced accuracy was 0.577 (95% CI 0.479 to 0.665), with sensitivity of 0.558 (95% CI 0.422 to 0.691) and specificity of 0.595 (95% CI 0.494 to 0.698). Positive predictive value was 0.460 (95% CI 0.338 to 0.576) and negative predictive value was 0.685 (95% CI 0.578 to 0.789). Unlike the ideation model, the EBAL for suicidal behavior retained all 20 candidate predictors, indicating that the evidence-guided penalty differentials were insufficient to achieve meaningful variable selection for this outcome.

#### Standard LASSO comparator

The standard LASSO performed comparably for suicidal behavior, with an AUROC of 0.592, balanced accuracy of 0.584, sensitivity of 0.596, and specificity of 0.571. Positive predictive value was 0.463 (95% CI 0.344 to 0.586) and negative predictive value was 0.696 (95% CI 0.583 to 0.803). The standard LASSO also retained all 20 features for this outcome.

#### Head-to-head comparison

Neither model achieved clinically meaningful discrimination for suicidal behavior (Figure 5A). Post-hoc tests confirmed the absence of any difference between the two approaches for this outcome. McNemar’s exact test found only four discordant predictions, evenly split between EBAL and the standard LASSO (exact two-sided p = 1.000). The DeLong test comparing the two AUROCs was not significant (z = 0.19, p = 0.853), and none of the paired bootstrap tests for accuracy (p = 1.000), balanced accuracy (p = 0.680), F1 score (p = 0.409), or Matthews correlation coefficient (p = 0.689) approached significance. Both models retained all 20 features and performed near chance for suicidal behavior, confirming that the evidence-guided penalty information provided no measurable advantage for this outcome.

**Figure 5.**
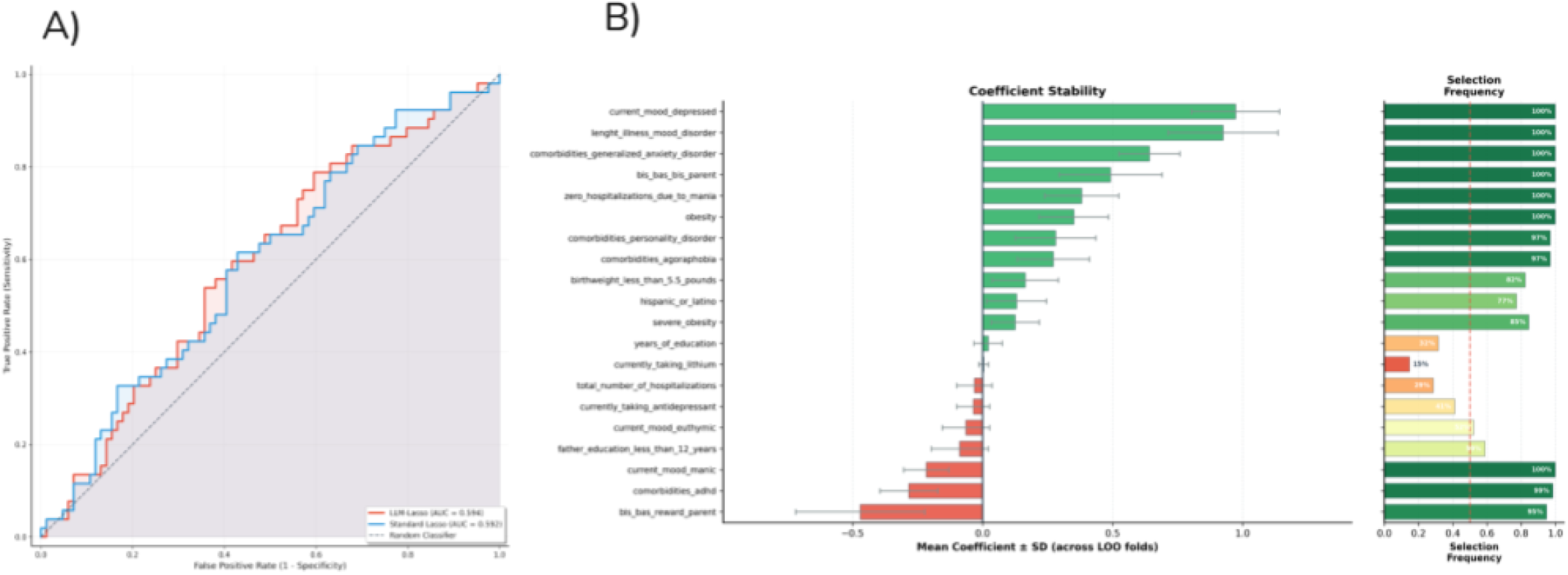
Classification performance for suicidal behavior and predictor stability for suicidal ideation. **A)** Receiver operating characteristic curves comparing Evidence-Based Al LASSO (AUROC = 0.594) and standard LASSO (AUROC = 0.592) for the secondary outcome of suicidal behavior (MINI-KID item B1c, N= 136, 52 positive and 84 negative). Neither model achieved clinically meaningful discrimination for suicidal behavior, with both curves near the diagonal reference line. **B)** Coefficient stability of the Evidence-Based Al LASSO model for suicidal ideation (MINI-KID item BIa) across 136 leave-one-out cross-validation folds. Each bar represents the mean coefficient value for a predictor, with error bars indicating the standard deviation across folds, Selection frequency (percentage of folds in which each predictor received a non-zero coefficient) is shown alongside each bar. The seven most stable predictors, including current depressed mood, duration of mood disorder illness and comorbid generalized anxiety disorder, were selected in t00% of iterations.

## Discussion

In this study, we developed Evidence-Based AI LASSO (EBAL), a hybrid pipeline that translates curated clinical evidence into feature-specific penalty factors for regularized regression, and applied it to predict suicidal ideation and suicidal behavior in youth with bipolar disorder. For suicidal ideation, EBAL produced a sparser model than standard LASSO, retaining 11 of 20 predictors compared with 18, and outperformed the standard LASSO comparator across every primary metric. These findings support the central premise of the framework, which is that embedding curated clinical evidence into the penalty structure before estimation yields a more clinically coherent predictor set than evidence-agnostic regularization. The classification gains were statistically supported by paired bootstrap tests, although the AUROC difference did not reach significance on the DeLong test, a pattern consistent with the modest sample size and the shared LASSO architecture of the two models. For suicidal behavior, however, EBAL did not improve prediction. Both models performed near chance and neither achieved meaningful variable selection. This outcome-specific divergence shapes both the clinical interpretation and the methodological boundaries of the approach.

The signed coefficients from the final EBAL model revealed a clinically interpretable structure for suicidal ideation in pediatric bipolar disorder. Predictors with positive coefficients clustered around internalizing symptoms, illness chronicity, and physical health burden, while negative coefficients clustered around non-depressive mood states and externalizing or reward-approach traits. Importantly, this separation was not specified a priori by the investigators but rather, emerged from the interaction between evidence-derived penalty weights and the observed data. A post-hoc review of the RAG retrieval records confirmed that the penalty scores were traceable to specific publications in the curated corpus (Figure 3). Two studies anchored most of this evidence base. (A. Lu et al., 2025), a systematic review of loneliness and suicidality across general populations and mood disorder samples, contributed evidence to 10 of 20 candidate predictors, while (Benarous et al., 2019), a systematic review of irritability and suicidal behaviors in children and adolescents, contributed to 8. The mechanistic frameworks described in these two reviews provide a lens for interpreting the retained predictor set.

The current depressed mood carried the largest positive coefficient. Indeed, depressive polarity has been consistently identified as the primary risk period for suicidal thoughts and behaviors in bipolar disorder across age groups, and the ISBD Task Force meta-analysis ranked it among the strongest correlates of suicide attempts across 13 pooled variables (Goldstein et al., 2009; Hauser et al., 2013; Schaffer et al., 2015). It is known that, depression produces hopelessness, a state that narrows the individual’s capacity to envision a tolerable future and functions as a proximal mediator of suicidal ideation. Equally important, are the psychosocial consequences of depressive episodes. For example, withdrawal from peers, academic disengagement, and disrupted family relationships amplify the sense of isolation that (A. Lu et al., 2025) identified as a consistent correlate of suicidality across populations. Their review further showed that depressive symptoms mediate the relationship between loneliness and suicidality, suggesting that depression does not simply co-occur with social isolation but actively produces the conditions through which isolation elevates suicide risk.

Duration of mood disorder illness carried the second largest positive coefficient. The mechanisms linking illness duration to suicidal ideation operate at three converging levels. Clinically, longer illness trajectories are associated with more frequent depressive recurrence, greater cumulative exposure to suicide-relevant cognitive states, and progressive psychosocial impairment across academic, family, and peer domains (Birmaher et al., 2006; Goldstein et al., 2009; Hauser et al., 2013). Repeated mood episodes may progressively sensitize stress-responsive circuitry, lowering the threshold for subsequent episodes and amplifying their affective intensity. On the other hand, prolonged illness compounds loneliness through direct social withdrawal during depressive episodes and through the indirect effects of stigma, functional impairment, and peer estrangement that accumulate over the illness course. Notably, (A. Lu et al., 2025) emphasized that the experience and causes of loneliness vary across developmental stages, and the cumulative psychosocial burden of pediatric bipolar disorder may be particularly potent during adolescence, when peer connection and identity formation are central to neurodevelopmental.

Comorbid generalized anxiety disorder carried the third largest positive coefficient. The ISBD meta-analysis identified comorbid anxiety as a significant independent correlate of suicide attempts in bipolar disorder (Schaffer et al., 2015), and subsequent work has shown that co-occurring anxiety worsens disease severity, disrupts sleep architecture, and reduces remission rates in adolescents with bipolar depression. The combination of depressive hopelessness and anxious hyperarousal may produce a particularly volatile cognitive state in which the individual simultaneously feels unable to escape current suffering and unable to tolerate it. Anxiety also sustains rumination, a repetitive cognitive process that maintains attention on negative self-referential content and mediates the link between negative affect and suicidal ideation. (Benarous et al., 2019) extend this picture by proposing that irritability constitutes a risk factor for suicidal ideation specifically through the onset of internalized disorder, and GAD may represent one of the primary clinical expressions of that pathway in youth with bipolar disorder.

The other positive predictors extend this internalizing framework. For example, the absence of prior mania-related hospitalization may index a predominantly depressive illness course, which carries higher suicidal risk because these patients accumulate more time in depressive states and may receive less intensive clinical monitoring than those whose manic episodes trigger hospitalization (Schaffer et al., 2015). Obesity, in turn, has been linked to suicidality through both psychosocial pathways involving body image distress and weight-related stigma. Elevated parent-reported behavioral inhibition system (BIS) scores reflect a temperamental sensitivity to punishment and threat that predisposes to anxiety and behavioral withdrawal, a profile that during a depressive episode may amplify the cognitive and social constriction that (A. Lu et al., 2025) describe as the “loneliness-to-suicidality” pathway. Finally, comorbid personality disorder, the weakest positive predictor, likely captures trait-level emotional dysregulation and interpersonal instability that compound mood episode severity.

Most notably, the four negative coefficients tell a complementary story. Current manic mood carried the largest inverse coefficient, followed by current euthymic mood. These observations indicate that being assessed in a manic or stable mood state lowered the predicted probability of suicidal ideation relative to participants assessed during a depressive episode. Manic states are characterized by grandiosity, elevated self-esteem, and increased social engagement, all of which are incompatible with the hopelessness and interpersonal isolation that drive the pathways described by (A. Lu et al., 2025) and (Benarous et al., 2019). On the other hand, euthymia, reflects a period of relative mood stability in which cognitive function and social connectedness are preserved, buffering against the ruminative processes that sustain suicidal ideation during depressive episodes. Comorbid ADHD also carried a negative coefficient, which aligns with the dimensional framework proposed by (Benarous et al., 2019) in which irritability increases suicide risk primarily through internalizing rather than externalizing pathways. Youth whose presentation is weighted toward externalizing features may therefore be relatively protected against the ruminative internalizing pathway to suicidal ideation (Axelson et al., 2006). The small negative coefficient for parent-reported BAS reward responsiveness fits the same pattern. A reward-approach temperament orients behavior toward engagement and goal pursuit, which may protect against the withdrawal and avoidance that sustain the internalizing pathway to suicidal ideation discussed earlier.

Predictors eliminated by EBAL are equally informative. The standard LASSO kept 18 of 20 predictors, including paternal education, ethnicity, low birthweight, severe obesity, years of education, current lithium use, current antidepressant use, total hospitalizations, and comorbid agoraphobia. However, none of these predictors are established proximal risk factors in the pediatric bipolar suicidality literature, and their retention likely reflects sample-specific correlations as opposed to generalizable clinical signals. EBAL eliminated all nine of these variables, producing a model in which every retained predictor can be evaluated against the clinical and mechanistic frameworks as explored previously (Figure 4B). Importantly, this elimination was not driven by manual investigator judgment but by penalty weights derived from a traceable evidence corpus, offering a level of methodological transparency that conventional data-driven feature selection methods do not provide.

Neither EBAL nor standard LASSO achieved meaningful classification for suicidal behavior. This finding aligns with a longstanding challenge in the field. Suicidal ideation and suicidal behavior, while clinically related, differ in their proximal drivers. Ideation tends to build through sustained cognitive and affective processes such as hopelessness, rumination, and social isolation. These processes - which unfold over days to weeks map well onto the clinical, illness chronicity and psychosocial variables in our predictor set. Suicidal behavior, by contrast, often emerges acutely in response to situational triggers such as interpersonal conflict, perceived humiliation, or sudden access to means (Franklin et al., 2017). In their review, (Benarous et al., 2019) noted that suicide attempts in children and adolescents are frequently sudden and unplanned, with little preceding ideation. They further proposed that irritability may predispose to attempts through a pathway involving disproportionate reactivity to acute frustration rather than through sustained internalized distress. These acute, state-dependent constructs are rarely captured in structured clinical assessments, and the intake battery used in the present study was no exception. This predictor-outcome mismatch, likely explains the failure of the model in predicting suicidal behavior rather than a failure of either EBAL or standard lasso modeling strategy.

The methodological contribution of this work extends beyond predicting suicidality in pediatric bipolar disorder. The core principle of EBAL is that curated clinical evidence can be translated into feature-specific penalty weights through a language model and used to constrain regularized regression. This principle is not specific to suicidality or to pediatric bipolar disorder and is applicable to any clinical prediction problem where a structured evidence base relevant to the outcome exists and where feature selection instability is a concern. Most notably, EBAL builds directly on the LLM-LASSO framework introduced by (E. Zhang et al., 2025), which demonstrated that language model derived penalty factors could improve feature selection in genomic classification tasks. Therefore, our contribution is a systematic solution to the input-quality problem that limits the original framework’s clinical applicability. EBAL addresses this problem by restricting the language model’s input to a PRISMA-aligned, expert-screened evidence corpus composed exclusively of systematic reviews, meta-analyses, and clinical practice guidelines. This constraint ensures that the penalty weights encode the highest available level of evidence rather than an uncontrolled mixture of sources of varying quality and relevance.

Our approach is also complementary to other emerging strategies for integrating structured clinical knowledge into psychiatric prediction. For example, (Gao et al., 2025) recently demonstrated that LLM-constructed knowledge graphs containing over 10 million biomedical relations could enhance prediction of major depression, anxiety, and bipolar disorder in the UK Biobank, with AUROC improvements ranging from 0.02 to 0.21. However, with a noticeable difference, their mental disorders knowledge graph operates at the representation level, enriching patient feature vectors with graph-derived embeddings that encoding relationships between clinical entities. Our solution on the other hand - EBAL, operates at the regularization level, using LLM-mediated evidence synthesis to constrain which features survive penalization. Notably, both approaches share the premise that structured domain knowledge improves psychiatric prediction over purely data-driven methods, but they intervene at different points in the modeling pipeline.

A deliberate design choice in this pipeline was the use of open-weight language models with publicly available parameters. In practice, this means that the exact models that generated the penalty weights reported in this study can be downloaded, inspected, and redeployed by any research group without requiring access to a commercial API or a subscription to a cloud service. This distinction matters for clinical research because proprietary cloud-based models can change their underlying parameters, training data, or response behavior between software versions without notification, which means that a penalty score generated today may not be reproducible in future even when the same prompt and evidence are used. Open-weight models eliminate this dependency entirely. The model weights are fixed, version controlled, and permanently archived, so another team running the same pipeline on the same evidence corpus will obtain identical penalty scores. To further support this transparency, the full structured prompt template and the complete language model reasoning trace are provided in Supplementary Methods S2 and S3, respectively. Even more importantly for psychiatric research, open-weight models can be deployed entirely on institutional hardware, which means that no clinical data, diagnostic information, or patient-level features ever leave the clinical site. In studies involving sensitive outcomes such as suicidal ideation, where even de-identified predictor sets may carry re-identification risk, the ability to run the full pipeline within the institutional computing environment without transmitting data to an external server is not merely a convenience but a governance requirement.

Several limitations of the current study should be considered. First, the study used a cross-sectional design, which means that the retained predictors represent correlates of current suicidal ideation status rather than prospective risk factors for future ideation onset. Second, the sample was drawn from a single academic site with 136 participants, and no external validation was performed on an independent cohort. The modest sample size also limits statistical power to detect small but clinically meaningful differences between models, which may partly explain why the AUROC difference did not reach significance on the DeLong test despite consistent advantages across all other metrics. Third, the evidence synthesis was conducted by a single expert reviewer, introducing potential screening bias that dual independent screening with formal inter-rater reliability assessment would mitigate. Fourth, only 20 of the available clinical variables were included as candidate predictors, and a richer feature set might improve performance for both outcomes. Lastly, the LLM penalty scores were generated in a single deterministic pass, and multi-trial scoring with temperature variation could produce more stable estimates for predictors where the evidence is ambiguous.

A limitation specific to evidence-guided approaches deserves separate attention. If the cohort being analyzed has generated published studies that were included in the evidence corpus, the penalty weights would be partially informed by findings derived from the same patient population, creating a form of circular inference analogous to the double-dipping problem described in neuroimaging (dos Santos Machado et al., n.d.; Kriegeskorte et al., 2010, 2009; Mwangi et al., 2014; Passos et al., 2016). The severity of this concern depends on the overlap between the analytic sample and the populations described in the evidence corpus, and on whether the specific outcome being predicted was the focus of those prior publications. In the present study, the Greater Houston Area Bipolar Registry has been described in a published protocol paper (Diaz et al., 2021) but has not, generated published findings on suicidal ideation outcomes that were included in the 40 evidence documents. Nevertheless, this concern is generalizable, and investigators adopting the framework should verify that the evidence corpus does not contain studies from the same analytic sample with the same outcome. This risk could be mitigated by excluding cohort-specific publications or by conducting sensitivity analyses with and without potentially overlapping sources.

External validation on an independent pediatric bipolar cohort is the most immediate next step. A multisite validation study would test whether the evidence-guided penalty structure generalizes to a different patient population and whether the same 11 predictors are retained, providing the strongest available test of feature selection stability. Notably, beyond pediatric bipolar disorder, the framework is directly transferable to other clinical prediction contexts where structured evidence exists and where interpretable, sparse models are preferred. EBAL demonstrates that the principles of evidence-based medicine can be operationalized within the penalty structure of a regularized regression model, offering a reproducible and transparent framework for aligning clinical prediction with prior knowledge rather than treating feature selection as a purely data-driven exercise.

## Data Availability

Software code and models used in this study are openly shared and available on github.

## Acknowledgements

This work was supported by a NECMHR01 grant from the Texas Child Mental Health Care Consortium (TCMHCC), Dunn Foundation and The National Institute of Health (NIH) AIM-AHEAD consortium.

## References

Agnew-Blais, J., Danese, A., 2016. Childhood maltreatment and unfavourable clinical outcomes in bipolar disorder: a systematic review and meta-analysis. Lancet Psychiatry 3, 342–349.

American Academy of Child and Adolescent Psychiatry, 2001. Practice parameter for the assessment and treatment of children and adolescents with suicidal behavior. American Academy of Child and Adolescent Psychiatry. J. Am. Acad. Child Adolesc. Psychiatry 40, 24S–51S.

Awad, S., Sheikh, S., Ahmad, M., Abid, F., Mazhar, Z., Raza, S.H., 2024. Antidepressant Use Side Effects Among Adolescents: Integrated Review. Journal Population Therapeutics Clinical Pharmacology 31, 1852–1861.

Axelson, D., Birmaher, B., Strober, M., Gill, M.K., Valeri, S., Chiappetta, L., Ryan, N., Leonard, H., Hunt, J., Iyengar, S., Bridge, J., Keller, M., 2006. Phenomenology of children and adolescents with bipolar spectrum disorders. Arch. Gen. Psychiatry 63, 1139–1148.

Barbeito, S., Vega, P., Sánchez-Gutiérrez, T., Becerra, J.A., González-Pinto, A., Calvo, A., 2021. A systematic review of suicide and suicide attempts in adolescents with psychotic disorders. Schizophr. Res. 235, 80–90.

Belsher, B.E., Smolenski, D.J., Pruitt, L.D., Bush, N.E., Beech, E.H., Workman, D.E., Morgan, R.L., Evatt, D.P., Tucker, J., Skopp, N.A., 2019. Prediction models for suicide attempts and deaths: A systematic review and simulation: A systematic review and simulation. JAMA Psychiatry 76, 642–651.

Benarous, X., Consoli, A., Cohen, D., Renaud, J., Lahaye, H., Guilé, J.-M., 2019. Suicidal behaviors and irritability in children and adolescents: a systematic review of the nature and mechanisms of the association. Eur. Child Adolesc. Psychiatry 28, 667–683.

Besag, F.M.C., Vasey, M.J., Sharma, A.N., Lam, I.C.H., 2021. Efficacy and safety of lamotrigine in the treatment of bipolar disorder across the lifespan: a systematic review. Ther. Adv. Psychopharmacol. 11, 20451253211045870.

Birmaher, B., Axelson, D., Strober, M., Gill, M.K., Valeri, S., Chiappetta, L., Ryan, N., Leonard, H., Hunt, J., Iyengar, S., Keller, M., 2006. Clinical course of children and adolescents with bipolar spectrum disorders. Arch. Gen. Psychiatry 63, 175–183.

Boppana, L., Bhadoria, M., Kodali, R.K., 2024. An Open-Source RAG Architecture for LLMs. In: TENCON 2024 - 2024 IEEE Region 10 Conference (TENCON). Presented at the TENCON 2024 - 2024 IEEE Region 10 Conference (TENCON), IEEE, pp. 43–46.

Bratu, E.A., Moroianu, L.-A., Isailă, O.-M., Pleșea-Condratovici, C., Avram, O.-E., Drima, E., 2025. Parental mental health and suicidal behavior as predictors of adolescent suicidal ideation and attempts: A systematic review and meta-analysis. J. Clin. Med. 14, 6860.

Bruton, A.M., Wesemann, D.G., Machingo, T.A., Majak, G., Johnstone, J.M., Marshall, R.D., 2025. Ketamine for mood disorders, anxiety, and suicidality in children and adolescents: a systematic review. Eur. Child Adolesc. Psychiatry 34, 141–157.

Chawla, N.V., Bowyer, K.W., Hall, L.O., Kegelmeyer, W.P., 2002. SMOTE: Synthetic minority over-sampling technique. J. Artif. Intell. Res. 16, 321–357.

Cipriani, A., Hawton, K., Stockton, S., Geddes, J.R., 2013. Lithium in the prevention of suicide in mood disorders: updated systematic review and meta-analysis. BMJ 346, f3646.

Cleare, A., Pariante, C.M., Young, A.H., Anderson, I.M., Christmas, D., Cowen, P.J., Dickens, C., Ferrier, I.N., Geddes, J., Gilbody, S., Haddad, P.M., Katona, C., Lewis, G., Malizia, A., McAllister-Williams, R.H., Ramchandani, P., Scott, J., Taylor, D., Uher, R., Members of the Consensus Meeting, 2015. Evidence-based guidelines for treating depressive disorders with antidepressants: A revision of the 2008 British Association for Psychopharmacology guidelines. J. Psychopharmacol. 29, 459–525.

Collins, G.S., Moons, K., Dhiman, P., Riley, R., Beam, A.L., Van Calster, B., Ghassemi, M., Liu, X., Reitsma, J.B., van Smeden, M., Boulesteix, A., Camaradou, J., Celi, L., Denaxas, S., Denniston, A., Glocker, B., Golub, R.M., Harvey, H., Heinze, G., Hoffman, M.M., Kengne, A., Lam, E., Lee, N., Loder, E.W., Maier-Hein, L., Mateen, B., Mccradden, M., Oakden-Rayner, L., Ordish, J., Parnell, R., Rose, S., Singh, K., Wynants, L., Logullo, P., 2024. TRIPOD+AI statement: updated guidance for reporting clinical prediction models that use regression or machine learning methods. BMJ 385.

Conti, C., Lanzara, R., Scipioni, M., Iasenza, M., Guagnano, M.T., Fulcheri, M., 2017. The relationship between binge eating disorder and suicidality: A systematic review. Front. Psychol. 8, 2125.

Correll, C.U., Cortese, S., Croatto, G., Monaco, F., Krinitski, D., Arrondo, G., Ostinelli, E.G., Zangani, C., Fornaro, M., Estradé, A., Fusar-Poli, P., Carvalho, A.F., Solmi, M., 2021. Efficacy and acceptability of pharmacological, psychosocial, and brain stimulation interventions in children and adolescents with mental disorders: an umbrella review. World Psychiatry 20, 244–275.

da Costa Santos, C.M., de Mattos Pimenta, C.A., Nobre, M.R.C., 2007. The PICO strategy for the research question construction and evidence search. Rev. Lat. Am. Enfermagem 15, 508–511.

das Neves Peixoto, F.S., de Sousa, D.F., Luz, D.C.R.P., Vieira, N.B., Gonçalves Júnior, J., Dos Santos, G.C.A., da Silva, F.C.T., Rolim Neto, M.L., 2017. Bipolarity and suicidal ideation in children and adolescents: a systematic review with meta-analysis. Ann. Gen. Psychiatry 16, 22.

De Crescenzo, F., Serra, G., Maisto, F., Uchida, M., Woodworth, H., Casini, M.P., Baldessarini, R.J., Vicari, S., 2017. Suicide attempts in juvenile bipolar versus major depressive disorders: Systematic review and meta-analysis. J. Am. Acad. Child Adolesc. Psychiatry 56, 825–831.e3.

Deng, Y.-Y., Shi, Z.-M., Tang, Y.-Y., Li, Z., Cai, D.-B., Si, Z.-X., Yang, X.-H., Huang, X.-B., Zheng, W., 2026. Intermittent theta burst stimulation for adolescent patients with major depressive disorder or bipolar depression: A systematic review. J. Affect. Disord. 394, 120435.

Diaz, A.P., Cuellar, V.A., Vinson, E.L., Suchting, R., Durkin, K., Fernandes, B.S., Scaini, G., Kazimi, I., Zunta-Soares, G.B., Quevedo, J., Sanches, M., Soares, J.C., 2021. The Greater Houston Area Bipolar Registry-clinical and neurobiological trajectories of children and adolescents with bipolar disorders and high-risk unaffected offspring. Front. Psychiatry 12, 671840.

dos Santos Machado, C., Ballester, P.L., Cao, B., Mwangi, B., Caldieraro, M.A., Kapczinski, F., Passos, I.C., n.d. Prediction of suicide attempts in a prospective cohort study with a nationally representative sample of the US population. Psychol. Med. 1–12.

Døssing, E., Pagsberg, A.K., 2021. Electroconvulsive therapy in children and adolescents: A systematic review of current literature and guidelines. J. ECT 37, 158–170.

Durdurak, B.B., Altaweel, N., Upthegrove, R., Marwaha, S., 2022. Understanding the development of bipolar disorder and borderline personality disorder in young people: a meta-review of systematic reviews. Psychol. Med. 52, 3769–3782.

Franklin, J.C., Ribeiro, J.D., Fox, K.R., Bentley, K.H., Kleiman, E.M., Huang, X., Musacchio, K.M., Jaroszewski, A.C., Chang, B.P., Nock, M.K., 2017. Risk factors for suicidal thoughts and behaviors: A meta-analysis of 50 years of research. Psychol. Bull. 143, 187–232.

Gao, S., Yu, K., Yang, Y., Yu, S., Shi, C., Wang, X., Tang, N., Zhu, H., 2025. Large language model powered knowledge graph construction for mental health exploration. Nat Commun 16.

Goldstein, T.R., Birmaher, B., Axelson, D., Goldstein, B.I., Gill, M.K., Esposito-Smythers, C., Ryan, N.D., Strober, M.A., Hunt, J., Keller, M., 2009. Family environment and suicidal ideation among bipolar youth. Arch. Suicide Res. 13, 378–388.

Halfon, N., Labelle, R., Cohen, D., Guilé, J.-M., Breton, J.-J., 2013. Juvenile bipolar disorder and suicidality: a review of the last 10 years of literature. Eur. Child Adolesc. Psychiatry 22, 139–151.

Hammad, T.A., 2007. Review: benefits of antidepressants outweigh risks of suicidal ideation and attempts in children and adolescents. Evid. Based. Ment. Health 10, 108.

Hauser, M., Galling, B., Correll, C.U., 2013. Suicidal ideation and suicide attempts in children and adolescents with bipolar disorder: a systematic review of prevalence and incidence rates, correlates, and targeted interventions. Bipolar Disord. 15, 507–523.

Higgins, J.P.T., Davey Smith, G., Altman, D.G., Egger, M., 2022. Principles of systematic reviewing. Systematic Reviews in Health Research.

Iorfino, F., Hickie, I.B., Lee, R.S.C., Lagopoulos, J., Hermens, D.F., 2016. The underlying neurobiology of key functional domains in young people with mood and anxiety disorders: a systematic review. BMC Psychiatry 16, 156.

James, A., Lai, F.H., Dahl, C., 2004. Attention deficit hyperactivity disorder and suicide: a review of possible associations. Acta Psychiatr. Scand. 110, 408–415.

Janas-Kozik, M.H., Słopień, A., Remberk, B., Siwek, M., 2023. The place of selective serotonin reuptake inhibitors (SSRIs) in the treatment of depressive disorders in children and adolescents. Recommendations of the Main Board of the Polish Psychiatric Association. Part 2 - pharmacological properties and safety of use. Psychiatr. Pol. 57, 917–940.

Kostyrka-Allchorne, K., Stoilova, M., Bourgaize, J., Rahali, M., Livingstone, S., Sonuga-Barke, E., 2023. Review: Digital experiences and their impact on the lives of adolescents with pre-existing anxiety, depression, eating and nonsuicidal self-injury conditions - a systematic review. Child Adolesc. Ment. Health 28, 22–32.

Kriegeskorte, N., Lindquist, M.A., Nichols, T.E., Poldrack, R.A., Vul, E., 2010. Everything you never wanted to know about circular analysis, but were afraid to ask. J. Cereb. Blood Flow Metab. 30, 1551–1557.

Kriegeskorte, N., Simmons, W.K., Bellgowan, P.S.F., Baker, C.I., 2009. Circular analysis in systems neuroscience: the dangers of double dipping. Nat. Neurosci. 12, 535–540.

Kwon, W., 2025. vLLM: An Efficient Inference Engine for Large Language Models.

Lewis, P., Perez, E., Piktus, A., Petroni, F., Karpukhin, V., Goyal, N., Küttler, H., Lewis, M., Yih, W.-T., Rocktäschel, T., Riedel, S., Kiela, D., 2020. Retrieval-augmented generation for knowledge-intensive NLP tasks. arXiv [cs.CL].

Li, M., Zhang, Y., Long, D., Chen, K., Song, S., Bai, S., Yang, Z., Xie, P., Yang, A., Liu, D., Zhou, J., Lin, J., 2026. Qwen3-VL-embedding and Qwen3-VL-Reranker: A unified framework for state-of-the-art multimodal retrieval and ranking. arXiv [cs.CL].

Lu, A., Chan, A., Menon, T., Le, G.H., Wong, S., Ho, R., Mcintyre R, 2025. Association between loneliness and suicidality among general populations and persons with depressive and bipolar disorders: A systematic review. Journal of Affective Disorders 380, 777–801.

Lu, J., Huang, J., Gao, W., Wang, Z., Yang, N., Luo, Y., Guo, J., Pang, W.I.P., Lok, G.K.I., Rao, W., 2025. Interventions for suicidal and self-injurious related behaviors in adolescents with psychiatric disorders: a systematic review and meta-analysis. Transl. Psychiatry 15, 73.

Malhi, G.S., Gessler, D., Outhred, T., 2017. The use of lithium for the treatment of bipolar disorder: Recommendations from clinical practice guidelines. J. Affect. Disord. 217, 266–280.

Mitchell, R.H.B., Goldstein, B.I., 2014. Inflammation in children and adolescents with neuropsychiatric disorders: a systematic review. J. Am. Acad. Child Adolesc. Psychiatry 53, 274–296.

Moller, C.I., Davey, C.G., Badcock, P.B., Wrobel, A.L., Cao, A., Murrihy, S., Sharmin, S., Cotton, S.M., 2022. Correlates of suicidality in young people with depressive disorders: A systematic review. Aust. N. Z. J. Psychiatry 56, 910–948.

Moons, K.G.M., Damen, J.A.A., Kaul, T., Hooft, L., Andaur Navarro, C., Dhiman, P., Beam, A.L., Van Calster, B., Celi, L.A., Denaxas, S., Denniston, A.K., Ghassemi, M., Heinze, G., Kengne, A.P., Maier-Hein, L., Liu, X., Logullo, P., McCradden, M.D., Liu, N., Oakden-Rayner, L., Singh, K., Ting, D.S., Wynants, L., Yang, B., Reitsma, J.B., Riley, R.D., Collins, G.S., van Smeden, M., 2025. PROBAST+AI: an updated quality, risk of bias, and applicability assessment tool for prediction models using regression or artificial intelligence methods. BMJ 388, e082505.

Murad, M.H., Asi, N., Alsawas, M., Alahdab, F., 2016. New evidence pyramid. BMJ Evidence-Based Medicine 21, 125–127.

Mwangi, B., Jabbar Abd Sattar Hamoudi, H., Sanches, M., Dogan, N., Chaudhary, P., Wu, M.-J., Zunta-Soares, G.B., Soares, J.C., Martin, A., Soutullo, C.A., 2026. Human vs AI clinical assessment: Benchmarking a multimodal foundation model against multi-center expert judgment on the Mental Status Examination. medRxiv.

Mwangi, B., Tian, T.S., Soares, J.C., 2014. A review of feature reduction techniques in neuroimaging. Neuroinformatics 12, 229–244.

Nock, M.K., Green, J.G., Hwang, I., McLaughlin, K.A., Sampson, N.A., Zaslavsky, A.M., Kessler, R.C., 2013. Prevalence, correlates, and treatment of lifetime suicidal behavior among adolescents: results from the National Comorbidity Survey Replication Adolescent Supplement. JAMA Psychiatry 70, 300–310.

Nogueira, S., Brown, G., 2016. Measuring the stability of feature selection. In: Machine Learning and Knowledge Discovery in Databases, Lecture Notes in Computer Science. Springer International Publishing, Cham, pp. 442–457.

Orri, M., Perret, L.C., Turecki, G., Geoffroy, M.-C., 2018. Association between irritability and suicide-related outcomes across the life-course. Systematic review of both community and clinical studies. J. Affect. Disord. 239, 220–233.

Page, M.J., McKenzie, J.E., Bossuyt, P.M., Boutron, I., Hoffmann, T.C., Mulrow, C.D., Shamseer, L., Tetzlaff, J.M., Akl, E.A., Brennan, S.E., Chou, R., Glanville, J., Grimshaw, J.M., Hróbjartsson, A., Lalu, M.M., Li, T., Loder, E.W., Mayo-Wilson, E., McDonald, S., McGuinness, L.A., Stewart, L.A., Thomas, J., Tricco, A.C., Welch, V.A., Whiting, P., Moher, D., 2021. The PRISMA 2020 statement: an updated guideline for reporting systematic reviews. BMJ 372, n71.

Passos, I.C., Mwangi, B., Cao, B., Hamilton, J.E., Wu, M.-J., Zhang, X.Y., Zunta-Soares, G.B., Quevedo, J., Kauer-Sant’Anna, M., Kapczinski, F., Soares, J.C., 2016. Identifying a clinical signature of suicidality among patients with mood disorders: A pilot study using a machine learning approach. J. Affect. Disord. 193, 109–116.

Patel, R.S., Onyeaka, H., Youssef, N.A., 2020. Suicidal ideation and attempts in unipolar versus bipolar depression: analysis of 131,740 adolescent inpatients nationwide. Psychiatry Res. 291, 113231.

Pedregosa, F., Varoquaux, G., Gramfort, A., Michel, V., Thirion, B., Grisel, O., Duchesnay, 2011. Scikit-learn: Machine learning in Python. the Journal of machine Learning research 12, 2825–2830.

Pigoni, A., Delvecchio, G., Turtulici, N., Madonna, D., Pietrini, P., Cecchetti, L., Brambilla, P., 2024. Machine learning and the prediction of suicide in psychiatric populations: a systematic review. Transl. Psychiatry 14, 140.

Prades-Caballero, V., Navarro-Pérez, J.-J., Carbonell, Á., 2025. Factors associated with suicidal behavior in adolescents: An umbrella review using the Socio-ecological model. Community Ment. Health J. 61, 612–628.

Richardson, R., Connell, T., Foster, M., Blamires, J., Keshoor, S., Moir, C., Zeng, I.S., 2024. Risk and protective factors of self-harm and suicidality in adolescents: An umbrella review with meta-analysis. J. Youth Adolesc. 53, 1301–1322.

Sackett, D.L., Rosenberg, W.M., Gray, J.A., Haynes, R.B., Richardson, W.S., 1996. Evidence based medicine: what it is and what it isn’t. BMJ 312, 71–72.

Schaffer, A., Isometsä, E.T., Tondo, L., H Moreno, D., Turecki, G., Reis, C., Cassidy, F., Sinyor, M., Azorin, J., Kessing, L.V., Ha, K., Goldstein, T., Weizman, A., Beautrais, A., Chou, Y., Diazgranados, N., Levitt, A.J., Zarate, C.A., Jr, Rihmer, Z., Yatham, L.N., 2015. International Society for Bipolar Disorders Task Force on Suicide: meta-analyses and meta-regression of correlates of suicide attempts and suicide deaths in bipolar disorder. Bipolar Disorders 17, 1–16.

Serra, G., De Crescenzo, F., Maisto, F., Galante, J.R., Iannoni, M.E., Trasolini, M., Maglio, G., Tondo, L., Baldessarini, R.J., Vicari, S., 2022. Suicidal behavior in juvenile bipolar disorder and major depressive disorder patients: Systematic review and meta-analysis. J. Affect. Disord. 311, 572–581.

Seyedsalehi, A., Bailey, J., Ogonah, M.G.T., Fanshawe, T.R., Fazel, S., 2025. Prediction models for self-harm and suicide: a systematic review and critical appraisal. BMC Med. 23, 549.

Sheehan, D.V., Sheehan, K.H., Shytle, R.D., Janavs, J., Bannon, Y., Rogers, J.E., Milo, K.M., Stock, S.L., Wilkinson, B., 2010. Reliability and validity of the Mini International Neuropsychiatric Interview for children and adolescents (MINI-KID). J. Clin. Psychiatry 71, 313–326.

Singh, M.K., Gorelik, A.J., Stave, C., Gotlib, I.H., 2024. Genetics, epigenetics, and neurobiology of childhood-onset depression: an umbrella review. Mol. Psychiatry 29, 553–565.

Spittal, M.J., Guo, X.A., Kang, L., Kirtley, O.J., Clapperton, A., Hawton, K., Kapur, N., Pirkis, J., Carter, G., 2025. Machine learning algorithms and their predictive accuracy for suicide and self-harm: Systematic review and meta-analysis. PLoS Med. 22, e1004581.

Steyerberg, E.W., Vickers, A.J., Cook, N.R., Gerds, T., Gonen, M., Obuchowski, N., Pencina, M.J., Kattan, M.W., 2010. Assessing the performance of prediction models: a framework for traditional and novel measures: A framework for traditional and novel measures. Epidemiology 21, 128–138.

Tai, F., Pan, W., 2007. Incorporating prior knowledge of predictors into penalized classifiers with multiple penalty terms. Bioinformatics 23, 1775–1782.

Tao, Y.-J., Duan, X.-X., Liu, P., Wang, M.-W., Li, S.-X., Luo, T.-T., Xing, H.-Y., Huang, Y., 2025. Efficacy and safety of repetitive transcranial magnetic stimulation in youth with depression: a systematic review and meta-analysis of randomized sham-controlled trials. World J. Pediatr. 21, 1258–1274.

Thirunavukarasu, A.J., Ting, D.S.J., Elangovan, K., Gutierrez, L., Tan, T.F., Ting, D.S.W., 2023. Large language models in medicine. Nat. Med. 29, 1930–1940.

Tibshirani, R., 1996. Regression shrinkage and selection via the lasso. J. R. Stat. Soc. Series B Stat. Methodol. 58, 267–288.

Tibshirani, R.J., Efron, B., 1993. An introduction to the bootstrap. Monographs on statistics and applied probability 57, 1–436.

Vasudev, A., Macritchie, K., Vasudev, K., Watson, S., Geddes, J., Young, A.H., 2011. Oxcarbazepine for acute affective episodes in bipolar disorder. Cochrane Database Syst. Rev. CD004857.

Wasserman, D., Rihmer, Z., Rujescu, D., Sarchiapone, M., Sokolowski, M., Titelman, D., Carli, 2012. The European Psychiatric Association (EPA) guidance on suicide treatment and prevention. European Psychiatry 27, 129–141.

Witt, K.G., Hetrick, S.E., Rajaram, G., Hazell, P., Taylor Salisbury, T.L., Townsend, E., Hawton, K., 2021. Interventions for self-harm in children and adolescents. Cochrane Database Syst. Rev. 3, CD013667.

Xu, J., Guo, Z., Hu, H., Chu, Y., Wang, X., He, J., Wang, Y., Shi, X., He, T., Zhu, X., Lv, Y., Wang, Y., Guo, D., Wang, H., Ma, L., Zhang, P., Zhang, X., Hao, H., Guo, Z., Yang, B., Zhang, B., Ma, Z., Wei, X., Bai, S., Chen, K., Liu, X., Wang, P., Yang, M., Liu, D., Ren, X., Zheng, B., Men, R., Zhou, F., Yu, B., Yang, J., Yu, L., Zhou, J., Lin, J., 2025. Qwen3-Omni Technical Report. arXiv [cs.CL].

Yang, R., Zhao, Y., Tan, Z., Lai, J., Chen, J., Zhang, X., Sun, J., Chen, L., Lu, K., Cao, L., Liu, X., 2023. Differentiation between bipolar disorder and major depressive disorder in adolescents: from clinical to biological biomarkers. Front. Hum. Neurosci. 17, 1192544.

Zhang, E., Goto, R., Sagan, N., Mutter, J., Phillips, N., Alizadeh, A., Lee, K., Blanchet, J., Pilanci, M., Tibshirani, R., 2025. LLM-Lasso: A robust framework for domain-informed feature selection and regularization. arXiv [cs.LG].

Zhang, Y., Li, M., Long, D., Zhang, X., Lin, H., Yang, B., Xie, P., Yang, A., Liu, D., Lin, J., Huang, F., Zhou, J., 2025. Qwen3 embedding: Advancing text embedding and reranking through foundation models. arXiv [cs.CL].

Zou, H., 2006. The adaptive lasso and its oracle properties. J. Am. Stat. Assoc. 101, 1418–1429.

