## Supplemental materials for "Evidence-guided AI regularization for suicidal ideation prediction in pediatric bipolar disorder"

**Supplementary materials**

**Supplementary Methods S1. Database search strategies**

Search strategy (January 20th 2026) **PUBMED**

Comprehensive search strategy

| # | Search Terms | # of results |
| --- | --- | --- |
| 1 | ( "Mood Disorders"[Mesh] OR "Bipolar Disorder"[Mesh] OR "Depressive Disorder"[Mesh] OR "Depressive Disorder, Major"[Mesh] OR "Dysthymic Disorder"[Mesh] OR "Cyclothymic Disorder"[Mesh] OR mood disorder*[tiab] OR affective disorder*[tiab] OR bipolar disorder*[tiab] OR bipolar spectrum[tiab] OR BD[tiab] OR BD-I[tiab] OR BD-II[tiab] OR bipolar I[tiab] OR bipolar II[tiab] OR manic episode*[tiab] OR hypomanic episode*[tiab] OR mania[tiab] OR hypomania[tiab] OR major depressive disorder[tiab] OR MDD[tiab] OR depressive disorder*[tiab] OR depress*[tiab] OR unipolar depress*[tiab] OR dysthymi*[tiab] OR cyclothymi*[tiab] ) | 760570 |
| 2 | ( "Child"[Mesh] OR "Adolescent"[Mesh] OR "Pediatrics"[Mesh] OR "Adolescent Psychiatry"[Mesh] OR child*[tiab] OR adolescent*[tiab] OR pediatric*[tiab] OR paediatric*[tiab] OR youth*[tiab] OR teen*[tiab] OR juvenile*[tiab] OR young people[tiab] OR young person*[tiab] ) | 4470231 |
| 3 | ( "Suicidal Ideation"[Mesh] OR "Suicide, Attempted"[Mesh] OR "Suicide"[Mesh] OR "Self-Injurious Behavior"[Mesh] OR suicid*[tiab] OR suicidal ideation[tiab] OR suicidal thought*[tiab] OR suicide attempt*[tiab] OR self-harm[tiab] OR self harm[tiab] OR self-injur*[tiab] OR non-suicidal self-injury[tiab] OR NSSI[tiab] OR "Columbia-Suicide Severity Rating Scale"[tiab] OR C-SSRS[tiab] OR CSSRS[tiab] OR "Suicide Behavior Questionnaire"[tiab] OR SBQ[tiab] OR SBQ-R[tiab] OR "Beck Scale for Suicide Ideation"[tiab] OR BSSI[tiab] OR "Suicidal Ideation Questionnaire"[tiab] OR SIQ[tiab] OR "Self-Injurious Thoughts and Behaviors Interview"[tiab] OR SITBI[tiab] ) | 144591 |
| 4 | #1 AND #2 AND 3# | 16502 |

Search strategy (January 20th 2026) **PUBMED**

Narrow search strategy

| # | Search Terms | # of results |
| --- | --- | --- |
| 1 | ( ( "Bipolar Disorder"[Mesh] OR bipolar disorder[tiab] OR bipolar I[tiab] OR bipolar II[tiab] OR BD[tiab] OR BD-I[tiab] OR BD-II[tiab] ) AND ( "Child"[Mesh] OR "Adolescent"[Mesh] ) AND ( "Suicidal Ideation"[Mesh] OR "Suicide, Attempted"[Mesh] OR "Columbia-Suicide Severity Rating Scale"[tiab] OR C-SSRS[tiab] OR "Suicidal Ideation Questionnaire"[tiab] OR SIQ[tiab] OR "Beck Scale for Suicide Ideation"[tiab] OR BSSI[tiab] OR "Self-Injurious Thoughts and Behaviors Interview"[tiab] OR SITBI[tiab] ) ) | 451 |

Search Strategy 23/January/2026 **PUBMED**

| # | Search Terms | # of results |
| --- | --- | --- |
| 1 | ( "Mood Disorders"[Mesh] OR "Bipolar Disorder"[Mesh] OR "Depressive Disorder"[Mesh] OR "Depressive Disorder, Major"[Mesh] OR "Dysthymic Disorder"[Mesh] OR "Cyclothymic Disorder"[Mesh] OR mood disorder*[tiab] OR affective disorder*[tiab] OR bipolar disorder*[tiab] OR bipolar spectrum[tiab] OR BD[tiab] OR BD-I[tiab] OR BD-II[tiab] OR bipolar I[tiab] OR bipolar II[tiab] OR manic episode*[tiab] OR hypomanic episode*[tiab] OR mania[tiab] OR hypomania[tiab] OR major depressive disorder[tiab] OR MDD[tiab] OR depressive disorder*[tiab] OR depress*[tiab] OR unipolar depress*[tiab] OR dysthymi*[tiab] OR cyclothymi*[tiab] ) | 760570 |
| 2 | ( "Child"[Mesh] OR "Adolescent"[Mesh] OR "Pediatrics"[Mesh] "Paediatrics"[Mesh] OR "Adolescent Psychiatry"[Mesh] OR child*[tiab] OR adolescent*[tiab] OR pediatric*[tiab] OR paediatric*[tiab] OR youth*[tiab] OR teen*[tiab] OR juvenile*[tiab] OR "young people"[tiab] OR young person*[tiab] ) | 4470231 |
| 3 | ( "Suicidal Ideation"[Mesh] OR "Suicide, Attempted"[Mesh] OR "Suicide"[Mesh] OR "Self-Injurious Behavior"[Mesh] OR suicid*[tiab] OR suicidal ideation[tiab] OR suicidal thought*[tiab] OR suicide attempt*[tiab] OR self-harm[tiab] OR self harm[tiab] OR self-injur*[tiab] OR non-suicidal self-injury[tiab] OR NSSI[tiab] OR "Columbia-Suicide Severity Rating Scale"[tiab] OR C-SSRS[tiab] OR CSSRS[tiab] OR "Suicide Behavior Questionnaire"[tiab] OR SBQ[tiab] OR SBQ-R[tiab] OR "Beck Scale for Suicide Ideation"[tiab] OR BSSI[tiab] OR "Suicidal Ideation Questionnaire"[tiab] OR SIQ[tiab] OR "Self-Injurious Thoughts and Behaviors Interview"[tiab] OR SITBI[tiab] ) | 144684 |
| 4 | ( systematic review[pt] OR meta-analysis[pt] OR guideline[pt] OR practice guideline[pt] OR systematic[sb] OR "network meta-analysis"[tiab] OR "network meta analysis"[tiab] OR "umbrella review"[tiab] OR "overview of reviews"[tiab] OR "review of reviews"[tiab] OR "overview of systematic reviews"[tiab] ) NOT (protocol[ti] OR protocols[ti]) | 520557 |
| 5 | #1 AND #2 AND 3# AND #4 | 565 |

Search Strategy 23/January/2026 **EMBASE**

| # | Search Terms | # of results |
| --- | --- | --- |
| 1 | (  'mood disorder'/exp  OR 'bipolar disorder'/exp  OR 'depressive disorder'/exp  OR 'major depressive disorder'/exp  OR mood disorder*:ab,ti,kw  OR affective disorder*:ab,ti,kw  OR bipolar disorder*:ab,ti,kw  OR bipolar spectrum:ab,ti,kw  OR BD:ab,ti,kw  OR (BD NEAR/1 (I OR II OR 1 OR 2)):ab,ti,kw  OR (bipolar NEAR/1 (I OR II OR 1 OR 2)):ab,ti,kw  OR mania:ab,ti,kw  OR hypomania:ab,ti,kw  OR (manic NEAR/1 episode*):ab,ti,kw  OR (hypomanic NEAR/1 episode*):ab,ti,kw  OR major depressive disorder:ab,ti,kw  OR MDD:ab,ti,kw  OR depress*:ab,ti,kw  OR unipolar depress*:ab,ti,kw  OR dysthymi*:ab,ti,kw  OR cyclothymi*:ab,ti,kw  ) | 958826 |
| 2 | (  'child'/exp  OR 'adolescent'/exp  OR 'pediatrics'/exp  OR child*:ab,ti,kw  OR adolescen*:ab,ti,kw  OR pediatric*:ab,ti,kw  OR paediatric*:ab,ti,kw  OR youth*:ab,ti,kw  OR teen*:ab,ti,kw  OR juvenile*:ab,ti,kw  OR (young NEAR/1 people):ab,ti,kw  OR (young NEAR/1 person*):ab,ti,kw  ) | 5949546 |
| 3 | (  'suicidal ideation'/exp  OR 'suicide attempt'/exp  OR 'suicide'/exp  OR 'self injurious behavior'/exp  OR suicid*:ab,ti,kw  OR (suicidal NEAR/1 ideation):ab,ti,kw  OR (suicidal NEAR/1 thought*):ab,ti,kw  OR (suicide NEAR/1 attempt*):ab,ti,kw  OR self-harm:ab,ti,kw  OR 'self harm':ab,ti,kw  OR self-injur*:ab,ti,kw  OR (non-suicidal NEAR/1 self-injur*):ab,ti,kw  OR NSSI:ab,ti,kw  OR 'Columbia-Suicide Severity Rating Scale':ab,ti,kw  OR C-SSRS:ab,ti,kw  OR CSSRS:ab,ti,kw  OR 'Suicide Behavior Questionnaire':ab,ti,kw  OR SBQ:ab,ti,kw  OR SBQ-R:ab,ti,kw  OR 'Beck Scale for Suicide Ideation':ab,ti,kw  OR BSSI:ab,ti,kw  OR 'Suicidal Ideation Questionnaire':ab,ti,kw  OR SIQ:ab,ti,kw  OR SITBI:ab,ti,kw  ) | 209654 |
| 4 | (  'meta analysis'/de  OR 'systematic review'/de  OR 'guideline'/de  OR 'practice guideline'/de  OR meta-analys*:ab,ti,kw  OR 'systematic review':ab,ti,kw  OR 'network meta-analysis':ab,ti,kw  OR 'umbrella review':ab,ti,kw  OR 'overview of reviews':ab,ti,kw  OR guideline*:ab,ti,kw  OR 'practice guideline':ab,ti,kw  ) | 1988195 |
| 5 | #1 AND #2 AND 3# AND #4 | 1734 |

Search Strategy 23/January/2026 **OVID PsychInfo**

| # | Search Terms | # of results |
| --- | --- | --- |
| 1 | (  exp mood disorders/  OR exp bipolar disorder/  OR exp depressive disorders/  OR exp major depression/  OR (mood disorder* OR affective disorder* OR bipolar disorder* OR bipolar spectrum  OR (BD ADJ1 (I OR II OR 1 OR 2)) OR (bipolar ADJ1 (I OR II OR 1 OR 2))  OR mania OR hypomania OR (manic ADJ1 episode*) OR (hypomanic ADJ1 episode*)  OR "major depressive disorder" OR MDD OR depress* OR unipolar depress*  OR dysthymi* OR cyclothymi*  ).ti,ab.  ) | 445047 |
| 2 | (  'child'/exp  OR 'adolescent'/exp  OR 'pediatrics'/exp  OR child*:ab,ti,kw  OR adolescen*:ab,ti,kw  OR pediatric*:ab,ti,kw  OR paediatric*:ab,ti,kw  OR youth*:ab,ti,kw  OR teen*:ab,ti,kw  OR juvenile*:ab,ti,kw  OR (young NEAR/1 people):ab,ti,kw  OR (young NEAR/1 person*):ab,ti,kw  ) | 1121388 |
| 3 | (  exp suicidal ideation/  OR exp suicide attempt/  OR exp suicide/  OR exp self injurious behavior/  OR (suicid* OR (suicidal ADJ1 ideation) OR (suicidal ADJ1 thought*)  OR (suicide ADJ1 attempt*) OR self-harm OR "self harm" OR self-injur*  OR (non-suicidal ADJ1 self-injur*) OR NSSI  OR "Columbia-Suicide Severity Rating Scale" OR C-SSRS OR CSSRS  OR "Suicide Behavior Questionnaire" OR SBQ OR SBQ-R  OR "Beck Scale for Suicide Ideation" OR BSSI  OR "Suicidal Ideation Questionnaire" OR SIQ OR SITBI  ).ti,ab.  ) | 95822 |
| 4 | (  meta analysis/  OR systematic review/  OR practice guideline/  OR guidelines/  OR (meta-analys* OR "systematic review" OR "network meta-analysis"  OR "umbrella review" OR "overview of reviews" OR guideline* OR  "practice guideline"  ).ti,ab.  ) | 177686 |
| 5 | #1 AND #2 AND 3# AND #4 | 602 |

Search Strategy 23/January/2026 **Cochrane**

| # | Search Terms | # of results |
| --- | --- | --- |
| 1 | ([mh "Mood Disorders"] OR [mh "Bipolar Disorder"] OR [mh "Depressive Disorder"] OR mood disorder* OR affective disorder* OR bipolar disorder* OR bipolar spectrum OR BD OR BD1 OR BD2 OR mania OR hypomania OR manic episode* OR hypomanic episode* OR "major depressive disorder" OR MDD OR depress* OR unipolar depress* OR dysthymi* OR cyclothymi*) | 164202 |
| 2 | ([mh "Child"] OR [mh "Adolescent"] OR [mh "Pediatrics"] OR child* OR adolescen* OR pediatric* OR paediatric* OR youth* OR teen* OR juvenile* OR "young people" OR "young person") | 386725 |
| 3 | (  [mh "Suicidal Ideation"]  OR [mh "Suicide"]  OR [mh "Self-Injurious Behavior"]  OR suicid*  OR (suicidal NEXT ideation)  OR (suicidal NEXT thought*)  OR (suicide NEXT attempt*)  OR self-harm  OR "self harm"  OR self-injur*  OR (non-suicidal NEXT self-injur*)  OR NSSI  OR (Columbia NEXT Suicide NEXT Severity NEXT Rating NEXT Scale)  OR C-SSRS  OR CSSRS  OR (Suicide NEXT Behavior NEXT Questionnaire)  OR SBQ  OR SBQ-R  OR (Beck NEXT Scale NEXT for NEXT Suicide NEXT Ideation)  OR BSSI  OR (Suicidal NEXT Ideation NEXT Questionnaire)  OR SIQ  OR SITBI  ) | 11816 |
| 4 | (systematic review OR meta-analys* OR "network meta-analysis" OR "umbrella review" OR "overview of reviews" OR guideline OR "practice guideline") | 75318 |
| 5 | #1 AND #2 AND 3# AND #4 | 903 |

Search Strategy 23/January/2026 **WEB OF SCIENCE**

| # | Search Terms | # of results |
| --- | --- | --- |
| 1 | TS=(mood disorder* OR affective disorder* OR bipolar disorder* OR bipolar spectrum OR BD OR bipolar I OR bipolar II OR mania OR hypomania OR manic episode* OR hypomanic episode* OR major depressive disorder OR MDD OR depress* OR unipolar depress* OR dysthymi* OR cyclothymi*) | 1170427 |
| 2 | TS=(  child* OR adolescen* OR pediatric* OR paediatric* OR youth* OR teen* OR juvenile*  OR "young people" OR "young person*"  ) | 3752784 |
| 3 | TS=(  suicid* OR "suicidal ideation" OR "suicidal thought*" OR "suicide attempt*"  OR "self-harm" OR "self harm" OR self-injur* OR "non-suicidal self-injury" OR NSSI  OR C-SSRS OR CSSRS OR SBQ OR "SBQ-R" OR BSSI OR SIQ OR SITBI  ) | 169141 |
| 4 | TS=(  "systematic review" OR meta-analys* OR "network meta-analysis"  OR "umbrella review" OR "overview of reviews"  OR guideline* OR "practice guideline"  ) | 1591034 |
| 5 | #1 AND #2 AND 3# AND #4 | 1494 |

**Supplementary Table S3. Full-text studies excluded, with reasons (N = 152).**

| **Author (year)** | **Reason for exclusion** |
| --- | --- |
| Shaffer, D (2001) | Duplicate |
| Slawson, D (2006) | Wrong study design |
| Goldney, RD (2007) | Wrong study design |
| Memon, AM (2018) | Wrong population |
| Abascal-Peiró, S. (2023) | Wrong study design |
| Abdul Rahim, K. (2024) | Wrong population |
| Abid, M. (2025) | Wrong population |
| Alhassan, M. A. (2025) | Wrong population |
| Ang, S. H. (2025) | Wrong population |
| Arsenault-Lapierre, G. (2004) | Wrong population |
| Bahji, A. (2021) | Wrong population |
| Baldini, V. (2025) | Wrong population |
| Barbui, C. (2009) | Wrong population |
| Belvederi Murri, M. (2016) | Wrong population |
| Bennett, K. J. (2016) | Wrong study design |
| Beril Durdurak, B. (2022) | Wrong study design |
| Beynon, S. (2009) | Wrong outcome measure |
| Bochicchio, Lauren (2022) | Wrong population |
| Bordalo, F. (2022) | Wrong population |
| Boughdady, M. (2022) | Wrong study design |
| Bower, M. (2023) | Wrong population |
| Boylan, K. (2007) | Wrong study design |
| Brausch, A. M. (2012) | Wrong population |
| Bridge, Jeffrey A. (2007) | Wrong population |
| Caldwell, Deborah M. (2020) | Wrong study design |
| Calvo, Natalia (2022) | Wrong population |
| Cantor, N. (2023) | Wrong population |
| Cardoso, T. A. (2018) | Wrong population |
| Carrasco-Barrios, M. T. (2020) | Wrong population |
| Carter, G. (2003) | Wrong study design |
| Cheung, A. (2007) | Wrong study design |
| Cheung, A. (2009) | Wrong study design |
| Chiu, H. Y. (2018) | Wrong population |
| Cipriani, A. (2013) | Wrong population |
| Colic, L. (2021) | Wrong study design |
| Courtney, D. B. (2016) | Wrong study design |
| Courtney, Darren B. (2022) | Wrong population |
| Crowley, G. (2026) | Wrong population |
| De Codt, A. (2016) | Wrong study design |
| Detullio, David (2022) | Wrong study design |
| Du Toit, Stefani (2025) | Wrong study design |
| Du, Wei (2023) | Wrong population |
| Duarte, D. (2020) | Wrong population |
| Ernoul, A. (2016) | Wrong population |
| Fetter, Anna Kawennison (2023) | Wrong population |
| Fischer-Grote, L. (2024) | Wrong population |
| Fleischmann, A. (2005) | Wrong population |
| Fliege, H. (2009) | Wrong population |
| Forte, A. (2021) | Wrong population |
| French, A. (2023) | Wrong study design |
| French, B. (2024) | Wrong population |
| Geoffroy, Marie-Claude (2024) | Wrong study design |
| Gibbons, R. D. (2012) | Wrong population |
| Girela-Serrano, B. M. (2022) | Wrong outcome measure |
| Gol, Joanna (1995) | Wrong study design |
| Gong, W. (2025) | Wrong population |
| Grande, A. J. (2022) | Wrong population |
| Gøtzsche, P. C. (2017) | Wrong population |
| Hawton, K. (2015) | Wrong population |
| Hetrick, S. (2007) | Wrong study design |
| Hobbs, Elizabeth (2024) | Wrong population |
| Holtmann, M. (2006) | Wrong population |
| Hui-Ming, Niu (2023) | Wrong population |
| Iyengar, U. (2018) | Wrong study design |
| Jantarapakdee, Russunan (2025) | Wrong study design |
| Jiang, Y. (2025) | Wrong study design |
| Jin, S. S. (2021) | Wrong study design |
| Jurek, Lucie (2025) | Wrong population |
| Kaizar, E. E. (2006) | Wrong population |
| Karanikola, Maria N. K. (2018) | Wrong population |
| Karkın, A. N. (2023) | Wrong population |
| Kham-ai, Prasert (2026) | Wrong population |
| Kim, Jae-Won (2014) | Wrong study design |
| Kothgassner, Oswald D. (2020) | Wrong population |
| Labelle, Real (2015) | Wrong population |
| Lee, Aryb (2023) | Wrong population |
| Lee, G. (2023) | Wrong study design |
| Leigh, E. (2023) | Wrong population |
| Li, R. (2025) | Wrong population |
| Li, Wei (2014) | Wrong population |
| Li, X. Y. (2020) | Wrong population |
| Lim, H. C. (2025) | Wrong population |
| Ling, Y. (2025) | Wrong population |
| Liu, H. (2022) | Wrong population |
| Liu, Jen-Wei (2019) | Wrong population |
| Liu, R. T. (2022) | Wrong population |
| Livanou, M. (2016) | Wrong population |
| Livanou, Maria (2019) | Wrong population |
| Loyola, Gladys (2010) | Wrong population |
| Luo, J. (2024) | Wrong population |
| Maffre Maviel, G. (2025) | Wrong population |
| Makhija, Nita J. (2007) | Wrong outcome measure |
| March, J. S. (2006) | Wrong study design |
| Marchionatti, L. E. (2024) | Wrong outcome measure |
| Marconi, E. (2023) | Wrong population |
| Matra, Julian (2025) | Wrong population |
| McHugh, C. M. (2019) | Wrong population |
| McKay, Jamie Gamble (2024) | Wrong study design |
| McKetin, R. (2019) | Wrong population |
| Medina, Diana Chalán (2024) | Wrong population |
| Mento, Carmela (2022) | Wrong population |
| Miller, Matthew (2014) | Wrong population |
| Miranda-Mendizábal, A. (2017) | Wrong population |
| Mirkovic, B. (2014) | Wrong population |
| Mirsu-Paun, A. (2017) | Wrong study design |
| Morriss, R. (2007) | Wrong outcome measure |
| Mosholder, Andrew D. (2006) | Wrong outcome measure |
| Mosquera, L. (2016) | Wrong study design |
| Neary, Donna (2000) | Wrong population |
| Nielassoff, E. (2023) | Wrong study design |
| Oakley Browne, M. (2016) | Wrong study design |
| Ozinci, Z. (2018) | Wrong population |
| Patterson, S. (2024) | Wrong population |
| Peixoto, F. S. D. (2017) | Duplicate |
| Plener, P. L. (2017) | Wrong population |
| Pu, J. (2017) | Wrong population |
| Pérez Arqueros, V. (2023) | Wrong population |
| Qin, B. (2014) | Wrong population |
| Rahman, F. (2021) | Wrong population |
| Renaud-Charest, O. (2023) | Wrong population |
| Reyes, Madeleine (2020) | Wrong study design |
| Richard-Devantoy, S. (2016) | Wrong population |
| Rotenstein, L. S. (2016) | Wrong population |
| Sahle, B. W. (2022) | Wrong population |
| Sanchez, Maria Gonzalez (2024) | Wrong population |
| Schulte-Frankenfeld, P. M. (2024) | Wrong population |
| Sheldon, E. (2021) | Wrong population |
| Shooshtari, Mitra Hakim (2014) | Wrong population |
| Shropshire, Ali M. (2011) | Wrong population |
| Sideli, L. (2020) | Wrong population |
| Siu, Andrew M. H. (2019) | Wrong population |
| Soares-Weiser, K. (2007) | Wrong outcome measure |
| Soto-Sanz, V. (2019) | Wrong population |
| Souza, L. C. (2023) | Wrong population |
| Speckens, A. E. (2005) | Wrong population |
| Stanley, I. H. (2018) | Wrong population |
| Stocchero, B. A. (2024) | Wrong population |
| Suárez-Brito, P. (2024) | Wrong population |
| Tom, K. (2024) | Wrong population |
| Tsapakis, E. M. (2008) | Wrong study design |
| Vaudreuil, C. (2019) | Wrong population |
| Velasco, A. (2023) | Wrong population |
| Victor, S. E. (2014) | Wrong outcome measure |
| von Sydow, Kirsten (2006) | Wrong population |
| Wang, M. (2025) | Wrong population |
| Waraan, L. (2023) | Wrong population |
| Wohlfarth, Tamar D. (2006) | Wrong population |
| Wyatt, L. C. (2015) | Wrong population |
| Yiu, H. W. (2021) | Wrong population |
| Yu, Minghua (2026) | Wrong population |
| Zhou, X. (2020) | Wrong population |
| Zuriaga, A. (2021) | Wrong population |

*One report (Van Meter et al., 2011) could not be retrieved and is reported separately as “reports not retrieved” (n = 1), not as an exclusion. “Duplicate” denotes a duplicate report of a study included elsewhere.*

**References (excluded studies)**

Shaffer, D. 2001. “Summary of the Practice Parameters for the Assessment and Treatment of Children and Adolescents with Suicidal Behavior.” *Journal of the American Academy of Child & Adolescent Psychiatry* 40 (4): 495–499.

Slawson, D. 2006. “Antidepressant Drugs Increase Suicide Risk in Children.” *Journal of Family Practice* 55 (6): 488.

Goldney, RD. 2007. “All Ssris Can Cause Increased Risk of Suicide or Self-Harm in Youths with Depression.” *Australian Journal of Pharmacy* 88 (1049): 86.

Memon, AM. 2018. “The Role of Online Social Networking on Deliberate Self-Harm and Suicidality in Adolescents: A Systematized Review of Literature.” *Indian Journal of Psychiatry* 60 (4): 384–392.

Abascal-Peiró, S., A. Alacreu-Crespo, I. Peñuelas-Calvo, B. Ezquerra-de la Cruz, L. Jiménez-Muñoz, E. Baca-García, and A. Porras-Segovia. 2023. “Factors Associated with First Suicide Attempt Vs. Re-Attempt in Children and Adolescents: A Systematic Review and Meta-Analysis.” *European Psychiatry* 66: S584.

Abdul Rahim, K., N. J. Egglestone, I. G. Tsagareli, W. Usmani, S. Meherali, and Z. S. Lassi. 2024. “Mental Health Outcomes Beyond the Post-Partum Period Among Adolescent Mothers: A Systematic Review and Meta-Analysis.” *Health Psychol Behav Med* 12 (1): 2305741.

Abid, M., A. Iqbal, W. Mansell, A. Khaliq, W. Shehzad, and S. Shahzad. 2025. “Suicide Prevention Psychosocial Interventions for Youth in Low- and Middle-Income Countries: Systematic Review.” *BJPsych Open* 11 (6): e280.

Alhassan, M. A., M. A. Alarabi, W. M. Albalawi, A. Alkhodairi, K. A. Alghalayini, A. K. Alsomari, and H. A. Al Fiaar. 2025. “Prevalence and Risk Factors of Suicidal Ideation and Suicide Attempts Among Young People in the MENAT Region: A Systematic Review and Meta-Analysis.” *BMC Psychiatry* 25 (1): 1149.

Ang, S. H., S. Venkateswaran, M. B. Goda, K. N. C. Naidu, G. K. Kundadak, and M. Subramaniam. 2025. “Prevalence of Suicidal Behaviour in Adolescents and Youth at Ultra-High Risk for Psychosis: A Systematic Review and Meta-Analysis.” *European Psychiatry* 68 (1): e56.

Arsenault-Lapierre, G., C. Kim, and G. Turecki. 2004. “Psychiatric Diagnoses in 3275 Suicides: A Meta-Analysis.” *BMC Psychiatry* 4: 37.

Bahji, A., M. Pierce, J. Wong, J. N. Roberge, I. Ortega, and S. Patten. 2021. “Comparative Efficacy and Acceptability of Psychotherapies for Self-Harm and Suicidal Behavior Among Children and Adolescents: A Systematic Review and Network Meta-Analysis.” *JAMA Netw Open* 4 (4): e216614.

Baldini, V., C. Gottardi, R. Di Stefano, L. V. Rindi, G. Pazzocco, G. Varallo, M. Purgato, D. De Ronchi, C. Barbui, and G. Ostuzzi. 2025. “Association Between Adverse Childhood Experiences and Suicidal Behavior in Affective Disorders: A Systematic Review and Meta-Analysis.” *European Psychiatry* 68 (1): e58.

Barbui, C., E. Esposito, and A. Cipriani. 2009. “Selective Serotonin Reuptake Inhibitors and Risk of Suicide: A Systematic Review of Observational Studies.” *Cmaj* 180 (3): 291–297.

Belvederi Murri, M., D. Prestia, V. Mondelli, C. Pariante, S. Patti, B. Olivieri, C. Arzani, M. Masotti, M. Respino, M. Antonioli, L. Vassallo, G. Serafini, G. Perna, M. Pompili, and M. Amore. 2016. “The HPA Axis in Bipolar Disorder: Systematic Review and Meta-Analysis.” *Psychoneuroendocrinology* 63: 327–342.

Bennett, K. J., and S. J. Duda. 2016. “Towards Improved Systematic Review and Meta-Analysis Quality in Child and Adolescent Mental Health: A Systematic Review.” *Journal of the American Academy of Child and Adolescent Psychiatry* 55 (10): S173.

Beril Durdurak, B., R. Upthegrove, N. Altaweel, and S. Marwaha. 2022. “Understanding the Developmental Pathways and Onset of Bipolar Disorder and Borderline Personality Disorder in Young People: A Systematic Review of Reviews.” *BJPsych Open* 8: S60.

Beynon, S., K. Soares-Weiser, N. Woolacott, S. Duffy, and J. R. Geddes. 2009. “Pharmacological Interventions for the Prevention of Relapse in Bipolar Disorder: A Systematic Review of Controlled Trials.” *Journal of Psychopharmacology* 23 (5): 574–591.

Bochicchio, Lauren, Kelsey Reeder, Andre Ivanoff, Hunter Pope, and Ana Stefancic. 2022. “Psychotherapeutic Interventions for LGBTQ + Youth: A Systematic Review.” *Journal of LGBT Youth* 19 (2): 152–179.

Bordalo, F., and I. P. Carvalho. 2022. “The Role of Alexithymia as a Risk Factor for Self-Harm Among Adolescents in Depression - a Systematic Review.” *Journal of Affective Disorders* 297: 130–144.

Boughdady, M., R. M. Cosme-Cruz, and B. Goubran. 2022. “3.124 Is There a Need to Expand Religion-Based CBT as a Treatment Option for Self-Harming Behaviors?.” *Journal of the American Academy of Child and Adolescent Psychiatry* 61 (10): S268.

Bower, M., S. Smout, A. Donohoe-Bales, S. O’Dean, L. Teesson, J. Boyle, D. Lim, A. Nguyen, A. L. Calear, P. J. Batterham, K. Gournay, and M. Teesson. 2023. “A Hidden Pandemic? an Umbrella Review of Global Evidence on Mental Health in the Time of COVID-19.” *Frontiers in Psychiatry* 14.

Boylan, K., and P. Szatmari. 2007. “Review: Antidepressants May Increase Risk of Self-Harm or Suicidal Behaviour in Children and Adolescents.” *Evidence-based Mental Health* 10 (3): 89–89.

Brausch, A. M., and S. K. Girresch. 2012. “A Review of Empirical Treatment Studies for Adolescent Nonsuicidal Self-Injury.” *Journal of Cognitive Psychotherapy* 26 (1): 3–18.

Bridge, Jeffrey A., Satish Iyengar, Cheryl B. Salary, Remy Barbe, Boris Birmaher, Harold Alan Pincus, Lulu Ren, and David A. Brent. 2007. “Clinical Response and Risk for Reported Suicidal Ideation and Suicide Attempts in Pediatric Antidepressant Treatment: A Meta-Analysis of Randomized Controlled Trials.” *JAMA: Journal of the American Medical Association* 297 (15): 1683–1696.

Caldwell, Deborah M., Sarah R. Davies, Sarah E. Hetrick, Jennifer C. Palmer, Paola Caro, Jose A. Lopez-Lopez, David Gunnell, Judi Kidger, James Thomas, Clare French, Emily Stockings, Rona Campbell, and Nicky J. Welton. 2020. “"school-Based Interventions to Prevent Anxiety and Depression in Children and Young People: A Systematic Review and Network Meta-Analysis": Correction.” *The Lancet Psychiatry* 7 (9): e59.

Calvo, Natalia, Sara Garcia-Gonzalez, Citlalli Perez-Galbarro, Christina Regales-Peco, Jorge Lugo-Marin, Josep-Antoni Ramos-Quiroga, and Marc Ferrer. 2022. “Psychotherapeutic Interventions Specifically Developed for NSSI in Adolescence: A Systematic Review.” *European Neuropsychopharmacology* 58: 86–98.

Cantor, N., M. Kingsbury, E. Warner, H. Landry, Z. Clayborne, R. Islam, and I. Colman. 2023. “Young Adult Outcomes Associated with Adolescent Suicidality: A Meta-Analysis.” *Pediatrics* 151 (3).

Cardoso, T. A., T. C. Mondin, L. B. Azevedo, L. M. D. Toralles, and L. D. de Mattos Souza. 2018. “Is Suicide Risk a Predictor of Diagnosis Conversion to Bipolar Disorder?.” *Psychiatry Research* 268: 473–477.

Carrasco-Barrios, M. T., P. Huertas, P. Martín, C. Martín, M. C. Castillejos, E. Petkari, and B. Moreno-Küstner. 2020. “Determinants of Suicidality in the European General Population: A Systematic Review and Meta-Analysis.” *Int J Environ Res Public Health* 17 (11).

Carter, G.. 2003. “Review: Evidence Is Lacking About Suicide Prevention in Young People.” *Evidence-based Mental Health* 6 (4): 121–121.

Cheung, A.. 2007. “Review: Antidepressant Use Increases the Risk of Suicidal Behaviour and Ideation in Children.” *Evidence-based Mental Health* 10 (1): 20–20.

Cheung, A.. 2009. “Review: Ssris Are Associated with Increased Risk for Attempted or Completed Suicide in Adolescents but Not in Adults or the Elderly.” *ACP Journal Club* 150 (6): 3p–3p.

Chiu, H. Y., H. C. Lee, P. Y. Chen, Y. F. Lai, and Y. K. Tu. 2018. “Associations Between Sleep Duration and Suicidality in Adolescents: A Systematic Review and Dose–response Meta-Analysis.” *Sleep Medicine Reviews* 42: 119–126.

Cipriani, A., K. Reid, A. H. Young, K. Macritchie, and J. Geddes. 2013. “Valproic Acid, Valproate and Divalproex in the Maintenance Treatment of Bipolar Disorder.” *Cochrane Database of Systematic Reviews* (10).

Colic, L., L. Villa, M. Dauvermann, L. Van Velzen, A. Sankar, D. Goldman, P. Panchal, J. Kim, L. Schmaal, A. L. Van Harmelen, and H. Blumberg. 2021. “Brain Structure Associated with Future Suicide Thoughts and Behaviors in Female Adolescents with Bipolar Disorder.” *Neuropsychopharmacology* 46: 270.

Courtney, D. B., S. J. Duda, J. Henderson, P. Szatmari, and K. J. Bennett. 2016. “Quality Appraisal of Clinical Practice Guidelines for Depression in Children and Adolescents.” *Journal of the American Academy of Child and Adolescent Psychiatry* 55 (10): S172.

Courtney, Darren B., Priya Watson, Karolin R. Krause, Benjamin W. C. Chan, Kathryn Bennett, Meredith Gunlicks-Stoessel, Terri Rodak, Kirsten Neprily, Tabitha Zentner, and Peter Szatmari. 2022. “Predictors, Moderators, and Mediators Associated with Treatment Outcome in Randomized Clinical Trials Among Adolescents with Depression: A Scoping Review.” *JAMA Network Open* 5 (2): e2146331–e2146331.

Crowley, G., S. Gnanapragasam, J. B. Fanshawe, H. Allberry, L. Bojanić, S. G. Tham, S. R. Hannam-Swain, F. Mughal, R. Pendrous, K. Russell, N. Kapur, M. I. Troya, and D. Knipe. 2026. “Characteristics of Individuals from Ethnic Minority Backgrounds Who Die by Suicide: A Systematic Review and Meta-Analysis.” *Journal of Affective Disorders* 398.

De Codt, A., P. Monhonval, X. Bongaerts, I. Belkacemi, and J. M. Tecco. 2016. “Bipolar Disorder and Early Affective Trauma.” *Psychiatria Danubina* 28: 4–8.

Detullio, David, Danielle H. Millen, and Tom D. Kennedy. 2022. “A Meta-Analysis of the Association Between the Children's Depression Inventory (CDI) and Suicidality.” *Aggression and Violent Behavior* 64: 1–11.

Du Toit, Stefani, Mark Tomlinson, Christina A. Laurenzi, Sarah Gordon, Laura Hartmann, Nina Abrahams, Melissa Bradshaw, Amanda Brand, G. J. Melendez-Torres, Chiara Servili, Tarun Dua, David A. Ross, Joanna Lai, and Sarah Skeen. 2025. “Psychosocial Interventions for Preventing Mental Health Conditions in Adolescents with Emotional Problems: A Meta-Analysis.” *Journal of Adolescent Health* 76 (2): 187–209.

Du, Wei, Yi Jie Jia, Fei Hong Hu, Meng Wei Ge, Yu Jie Cheng, Xin Qu, and Hong Lin Chen. 2023. “Prevalence of Suicidal Ideation and Correlated Risk Factors During the COVID-19 Pandemic: A Meta-Analysis of 113 Studies from 31 Countries.” *Journal of Psychiatric Research* 166: 147–168.

Duarte, D., R. Belzeaux, B. Etain, K. T. Greenway, E. Rancourt, H. Correa, G. Turecki, and S. Richard-Devantoy. 2020. “Childhood-Maltreatment Subtypes in Bipolar Patients with Suicidal Behavior: Systematic Review and Meta-Analysis.” *Braz J Psychiatry* 42 (5): 558–567.

Ernoul, A., M. Orsat, and G. Dubois de Prisque. 2016. “Sexual Assaults and Self-Cuttings in Adolescence.” *Annales Medico-Psychologiques* 174 (6): 442–447.

Fetter, Anna Kawennison, Andrea Wiglesworth, LittleDove F. Rey, Michael Azarani, Micah L. Prairie Chicken, Amanda R. Young, Amy Riegelman, and Joseph P. Gone. 2023. “Risk Factors for Suicidal Behaviors in American Indian and Alaska Native Peoples: A Systematic Review.” *Clinical Psychological Science* 11 (3): 528–551.

Fischer-Grote, L., V. Fössing, M. Aigner, E. Fehrmann, and M. Boeckle. 2024. “Effectiveness of Online and Remote Interventions for Mental Health in Children, Adolescents, and Young Adults After the Onset of the COVID-19 Pandemic: Systematic Review and Meta-Analysis.” *JMIR Mental Health* 11 (1).

Fleischmann, A., J. M. Bertolote, M. Belfer, and A. Beautrais. 2005. “Completed Suicide and Psychiatric Diagnoses in Young People: A Critical Examination of the Evidence.” *Am J Orthopsychiatry* 75 (4): 676–683.

Fliege, H., J. R. Lee, A. Grimm, and B. F. Klapp. 2009. “Risk Factors and Correlates of Deliberate Self-Harm Behavior: A Systematic Review.” *J Psychosom Res* 66 (6): 477–493.

Forte, A., G. Sarli, L. Polidori, D. Lester, and M. Pompili. 2021. “The Role of New Technologies to Prevent Suicide in Adolescence: A Systematic Review of the Literature.” *Medicina (Kaunas)* 57 (2).

French, A., K. Duffy, A. Cameron, J. Bower, and F. Nearchou. 2023. “Exploring Third Factors in the Relationship Between Sleep Difficulties and Suicidal Behaviour in Adolescents and Young Adults: A Systematic Review.” *Journal of Psychosomatic Research* 169.

French, B., G. Nalbant, H. Wright, K. Sayal, D. Daley, M. J. Groom, S. Cassidy, and C. L. Hall. 2024. “The Impacts Associated with Having ADHD: An Umbrella Review.” *Frontiers in Psychiatry* 15.

Geoffroy, M. C., N. Chadi, S. Bouchard, J. Fuoco, E. Chartrand, T. Loose, A. Sciola, J. T. Boruff, S. N. Iyer, Y. Sun, J. P. Gouin, S. M. Côté, and B. D. Thombs. 2024. “Mental Health of Canadian Youth: A Systematic Review and Meta-Analysis of Studies Examining Changes in Depression, Anxiety, and Suicide-Related Outcomes During the COVID-19 Pandemic.” *Can J Public Health* 115 (3): 408–424.

Gibbons, R. D., C. H. Brown, K. Hur, J. M. Davis, and J. J. Mann. 2012. “Suicidal Thoughts and Behavior with Antidepressant Treatment: Reanalysis of the Randomized Placebo-Controlled Studies of Fluoxetine and Venlafaxine.” *Archives of General Psychiatry* 69 (6): 580–587.

Girela-Serrano, B. M., M. Guerrero-Jiménez, A. D. V. Spiers, and L. Gutiérrez-Rojas. 2022. “Obesity and Overweight Among Children and Adolescents with Bipolar Disorder from the General Population: A Review of the Scientific Literature and a Meta-Analysis.” *Early Interv Psychiatry* 16 (2): 113–125.

Gol, Joanna. 1995. “Adolescent Suicide Attempts: A Meta-Analysis of Antecedent Variables.” *Dissertation Abstracts International: Section B: The Sciences and Engineering* 56 (6-B): 3445.

Gong, W., H. Zou, Z. Chen, R. Yan, H. Liu, and Z. Yao. 2025. “Meta-Analysis of the Association Between Childhood Trauma and Non-Suicidal Self-Injury Behavior in Patients with Depression.” *Chinese Journal of Psychiatry* 58 (1): 37–46.

Grande, A. J., C. Elia, C. Peixoto, P. T. C. Jardim, P. Dazzan, A. B. Veras, J. K. Cruickshank, M. I. D. Rosa, and S. Harding. 2022. “Mental Health Interventions for Suicide Prevention Among Indigenous Adolescents: A Systematic Review.” *Sao Paulo Med J* 140 (3): 486–498.

Gøtzsche, P. C., and P. K. Gøtzsche. 2017. “Cognitive Behavioural Therapy Halves the Risk of Repeated Suicide Attempts: Systematic Review.” *J R Soc Med* 110 (10): 404–410.

Hawton, K., K. G. Witt, T. L. Taylor Salisbury, E. Arensman, D. Gunnell, E. Townsend, K. van Heeringen, and P. Hazell. 2015. “Interventions for Self-Harm in Children and Adolescents.” *Cochrane Database Syst Rev* 2015 (12): Cd012013.

Hetrick, S. E., S. Merry, J. McKenzie, P. Sindahl, and M. Proctor. 2007. “Selective Serotonin Reuptake Inhibitors (ssris) for Depressive Disorders in Children and Adolescents.” *Cochrane Database of Systematic Reviews* (3).

Hobbs, Elizabeth, Rachel Reed, Julia Dorfman, and Adelaide S. Robb. 2024. “Psychopharmacology for Pediatric Bipolar Disorder.” *Pediatric psychopharmacology evidence: A clinician's guide.* : 453–524.

Holtmann, M., S. Bölte, and F. Poustka. 2006. “[suicidality in Depressive Children and Adolescents During Treatment with Selective Serotonin Reuptake Inhibitors. Review and Meta-Analysis of the Available Randomised, Placebo Controlled Trials].” *Nervenarzt* 77 (11): 1332–1337.

Hui-Ming, Niu, Zhang Zi-Ming, Mu Xiao-Mei, and Zhao Hai-Jing. 2023. “The Characteristics and Influencing Factors of Nonsuicidal Self-Injury of Adolescents with Depressive Disorder in China: A Meta-Analysis.” *Journal of Nervous & Mental Disease* 211 (6): 448–452.

Iyengar, U., N. Snowden, J. R. Asarnow, P. Moran, T. Tranah, and D. Ougrin. 2018. “A Further Look at Therapeutic Interventions for Suicide Attempts and Self-Harm in Adolescents: An Updated Systematic Review of Randomized Controlled Trials.” *Front Psychiatry* 9: 583.

Jantarapakdee, Russunan, Penpaktr Uthis, and Ratchaneekorn Upasen. 2025. “Risk Factors of Depression Among Adolescents with Non-Suicidal Self-Injury: A Systematic Review.” *Journal of Client Centered Nursing Care* 11 (4): 283–296.

Jiang, Y., Q. Liu, Y. Ding, and Y. Sun. 2025. “Systematic Review and Meta-Analysis of the Correlation Between Tinnitus and Mental Health.” *American Journal of Otolaryngology - Head and Neck Medicine and Surgery* 46 (3).

Jin, S. S., T. M. Dolan, A. A. Cloutier, E. Bojdani, and L. DeLisi. 2021. “Systematic Review of Depression and Suicidality in Child and Adolescent (CAP) Refugees.” *Psychiatry Research* 302: 114025.

Jurek, Lucie, Samuele Cortese, and Mikail Nourredine. 2025. “Long-Term Safety of Methyphenidate.” *Risques du methylphenidate au long cours.* 183 (3): 332–337.

Kaizar, E. E., J. B. Greenhouse, H. Seltman, and K. Kelleher. 2006. “Do Antidepressants Cause Suicidality in Children? a Bayesian Meta-Analysis.” *Clin Trials* 3 (2): 73–90; discussion 91-8.

Karanikola, Maria N. K., Anne Lyberg, Anne-Lise Holm, and Elisabeth Severinsson. 2018. “The Association Between Deliberate Self-Harm and School Bullying Victimization and the Mediating Effect of Depressive Symptoms and Self-Stigma: A Systematic Review.” *BioMed Research International* : 1–36.

Karkın, A. N., and M. Eskin. 2023. “Prevalence, Correlates, and Risk Factors of Suicidal Ideation and Attempts in Turkey.” *Neuropsychiatric Investigation* 61 (1): 19–36.

Kham-ai, Prasert, Mattika Chaichan, Narisara Sripo, Kanyaphat Pongchangyou, Dao Weiangkham, and Karen Heaton. 2026. “Pesticide Exposure and the Risk of Depression, Anxiety, and Suicide: A Meta-Analysis.” *Western Journal of Nursing Research* 48 (1): 102–113.

Kim, Jae-Won, Eva M. Szigethy, Nadine M. Melhem, Ester M. Saghafi, and David A. Brent. 2014. “Inflammatory Markers and the Pathogenesis of Pediatric Depression and Suicide: A Systematic Review of the Literature.” *The Journal of Clinical Psychiatry* 75 (11): 1242–1253.

Kothgassner, Oswald D., Kealagh Robinson, Andreas Goreis, Dennis Ougrin, and Paul L. Plener. 2020. “Does Treatment Method Matter? a Meta-Analysis of the Past 20 Years of Research on Therapeutic Interventions for Self-Harm and Suicidal Ideation in Adolescents.” *Borderline Personality Disorder and Emotion Dysregulation* 7.

Labelle, Real, Louise Pouliot, and Alain Janelle. 2015. “A Systematic Review and Meta-Analysis of Cognitive Behavioural Treatments for Suicidal and Self-Harm Behaviours in Adolescents.” *Canadian Psychology / Psychologie canadienne* 56 (4): 368–378.

Lee, Aryb, C. E. Low, C. E. Yau, J. Li, R. Ho, and C. S. H. Ho. 2023. “Lifetime Burden of Psychological Symptoms, Disorders, and Suicide Due to Cancer in Childhood, Adolescent, and Young Adult Years: A Systematic Review and Meta-Analysis.” *JAMA Pediatr* 177 (8): 790–799.

Lee, G., N. Zou, D. Durand, J. Hom, and R. Cáceda. 2023. “6.85 a Systematic Review on Changes in Suicide Attempts in Pediatric Population After the Start of the COVID-19 Pandemic.” *Journal of the American Academy of Child and Adolescent Psychiatry* 62 (10): S313.

Leigh, E., K. Chiu, and E. D. Ballard. 2023. “Social Anxiety and Suicidality in Youth: A Systematic Review and Meta-Analysis.” *Res Child Adolesc Psychopathol* 51 (4): 441–454.

Li, R., Y. Yue, X. Gu, L. Xiong, M. Luo, and L. Li. 2025. “Risk Prediction Models for Adolescent Suicide: A Systematic Review and Meta-Analysis.” *Psychiatry Research* 347: 116405.

Li, Wei, Yumei Wan, Juanjuan Ren, Ting Li, and Chunbo Li. 2014. “Appraisal of the Methodological Quality and Summary of the Findings of Systematic Reviews on the Relationship Between Ssris and Suicidality.” *Shanghai Archives of Psychiatry* 26 (5): 248–258.

Li, X. Y., X. H. Zhang, and Q. B. Jiang. 2020. “Research Progress in the Risk of Ssris for Causing Suicide in Children and Adolescents.” *Chinese Journal of New Drugs* 29 (18): 2098–2102.

Lim, H. C., H. K. H. Tsui, J. T. Wong, K. Y. Chen, F. Hau, D. C. F. Ma, J. Y. M. Tang, and S. K. W. Chan. 2025. “Childhood Trauma and Longitudinal Clinical Outcomes in Bipolar Affective Disorder: A Systematic Review.” *East Asian Arch Psychiatry* 35 (3): 185–193.

Ling, Y., Y. Gu, O. M. Solomon, L. Li, X. Chen, Y. Wang, and Y. Wei. 2025. “A Review of the Scope of Non-Suicidal Self-Injury Behavior in Adolescents with Depressive Disorders: An Analysis of Related Influencing Factors.” *BMC Psychiatry* 25 (1).

Liu, H., J. Kerzner, I. Demchenko, D. N. Wijeysundera, S. H. Kennedy, K. S. Ladha, and V. Bhat. 2022. “Nitrous Oxide for the Treatment of Psychiatric Disorders: A Systematic Review of the Clinical Trial Landscape.” *Acta Psychiatrica Scandinavica* 146 (2): 126–138.

Liu, J. W., Y. K. Tu, Y. F. Lai, H. C. Lee, P. S. Tsai, T. J. Chen, H. C. Huang, Y. T. Chen, and H. Y. Chiu. 2019. “Associations Between Sleep Disturbances and Suicidal Ideation, Plans, and Attempts in Adolescents: A Systematic Review and Meta-Analysis.” *Sleep* 42 (6).

Liu, R. T., R. F. L. Walsh, A. E. Sheehan, S. M. Cheek, and C. M. Sanzari. 2022. “Prevalence and Correlates of Suicide and Nonsuicidal Self-Injury in Children: A Systematic Review and Meta-Analysis.” *JAMA Psychiatry* 79 (7): 718–726.

Livanou, M., V. Furtado, and S. Singh. 2016. “Prevalence and Nature of Mental Disorders Among Young Offenders in Custody and Community: A Meta-Analysis.” *European Psychiatry* 33: S460.

Livanou, Maria, Vivek Furtado, Catherine Winsper, Annabelle Silvester, and Swaran P. Singh. 2019. “Prevalence of Mental Disorders and Symptoms Among Incarcerated Youth: A Meta-Analysis of 30 Studies.” *The International Journal of Forensic Mental Health* 18 (4): 400–414.

Loyola, Gladys. 2010. “Evidence-Based Treatment of Unipolar Depression in Adolescents.” *Evidence-based Treatment of Unipolar Depression in Adolescents* : 214 p–214 p.

Luo, J., J. Zhang, S. Ye, and Q. Liu. 2024. “Influence of Childhood Psychosocial Stress on Pubertal Emotional and Behavioral Problems: A Systematic Review.” *Chinese Journal of Evidence-Based Medicine* 24 (12): 1428–1435.

Maffre Maviel, G., C. Somma, C. Davisse-Paturet, G. Airagnes, and M. Melchior. 2025. “The Role of Depression in the Relationship Between Cannabis Use and Suicidal Behaviours: A Systematic Review and Meta-Analysis.” *Drug Alcohol Depend* 273: 112714.

Makhija, Nita J., and Leo Sher. 2007. “Preventing Suicide in Adolescents with Alcohol Use Disorders.” *Special Issue: Alcohol and adolescent medicine* 19 (1): 53–59.

March, J. S., B. J. Klee, and C. M. E. Kremer. 2006. “Treatment Benefit and the Risk of Suicidality in Multicenter, Randomized, Controlled Trials of Sertraline in Children and Adolescents.” *Journal of Child and Adolescent Psychopharmacology* 16 (1-2): 91–102.

Marchionatti, L. E., A. C. Campello, J. A. Veronesi, C. Ziebold, A. C. Tonon, C. B. Casella, J. L. Schafer, A. N. Madyun, A. Caye, C. Kieling, L. A. Rohde, G. V. Polanczyk, J. Mari, R. Rocha, L. Rosa, D. Rosa, Z. M. Sanchez, R. A. Bressan, S. Saxena, S. Evans-Lacko, P. Cuijpers, K. R. Merikangas, B. A. Kohrt, J. Bantjes, S. Reynolds, Z. Mneimneh, and G. A. Salum. 2024. “The Science of Child and Adolescent Mental Health in Brazil: A Nationwide Systematic Review and Compendium of Evidence-Based Resources.” .

Marconi, E., L. Monti, A. Marfoli, G. D. Kotzalidis, D. Janiri, C. Cianfriglia, F. Moriconi, S. Costa, C. Veredice, G. Sani, and D. P. R. Chieffo. 2023. “A Systematic Review on Gender Dysphoria in Adolescents and Young Adults: Focus on Suicidal and Self-Harming Ideation and Behaviours.” *Child Adolesc Psychiatry Ment Health* 17 (1): 110.

Matra, Julian. 2025. “A Systematic Review of Psychotherapeutic Interventions Aimed at Reducing Suicidal Ideation in Black American Adolescents.” *Dissertation Abstracts International Section A: Humanities and Social Sciences* 86 (4-A): No–Specified.

McHugh, C. M., A. Corderoy, C. J. Ryan, I. B. Hickie, and M. M. Large. 2019. “Association Between Suicidal Ideation and Suicide: Meta-Analyses of Odds Ratios, Sensitivity, Specificity and Positive Predictive Value.” *BJPsych Open* 5 (2).

McKay, Jamie Gamble. 2024. “Dialectical Behavior Therapy Skills Group Component as a Stand-Alone Treatment for Children, Adolescents and University Students: A Systematic Review and Meta-Analysis.” *Dissertation Abstracts International: Section B: The Sciences and Engineering* 85 (3-B): No–Specified.

McKetin, R., J. Leung, E. Stockings, Y. Huo, J. Foulds, J. M. Lappin, C. Cumming, S. Arunogiri, J. T. Young, G. Sara, M. Farrell, and L. Degenhardt. 2019. “Mental Health Outcomes Associated with of the Use of Amphetamines: A Systematic Review and Meta-Analysis.” *EClinicalMedicine* 16: 81–97.

Medina, Diana Chalán, Milton Eduardo Chalán Medina, María Soledad Carrión Cabrera, and Katherine Michelle González Guambaña. 2024. “Factores De Riesgo Asociados a Las Conductas Suicidas En Adolescentes: Un Artículo De Revisión.” *Nure Investigación* (129): 1–9.

Mento, Carmela, Maria Catena Silvestri, Maria Rosaria Anna Muscatello, Amelia Rizzo, Laura Celebre, Antonio Bruno, and Antonio Rocco Zoccali. 2022. “Psychological Pain and Risk of Suicide in Adolescence.” *International Journal of Adolescent Medicine and Health* 34 (3): 1–8.

Miller, Matthew, V. Pate, S. A. Swanson, D. Azrael, A. White, and T. Sturmer. 2014. “Antidepressant Class, Age, and the Risk of Deliberate Self-Harm: A Propensity Score Matched Cohort Study of SSRI and SNRI Users in the USA.” *CNS Drugs* 28 (1): 79–88.

Miranda-Mendizábal, A., P. Castellví, O. Parés-Badell, J. Almenara, I. Alonso, M. J. Blasco, A. Cebrià, A. Gabilondo, M. Gili, C. Lagares, J. A. Piqueras, M. Roca, J. Rodríguez-Marín, T. Rodríguez, V. Soto-Sanz, G. Vilagut, J. Alonso, and T. Rodríguez-Jiménez. 2017. “Sexual Orientation and Suicidal Behaviour in Adolescents and Young Adults: Systematic Review and Meta-Analysis.” *British Journal of Psychiatry* 211 (2): 77–87.

Mirkovic, B., V. Belloncle, C. Rousseau, A. Knafo, J. M. Guile, and P. Gerardin. 2014. “Prevention Strategies of Suicide and Suicidal Behavior in Adolescents: A Systematic Review.” *Strategies de prevention du suicide et des conduites suicidaires a l'adolescence : Revue systematique de la litterature.* 62 (1): 33–46.

Mirsu-Paun, A., and J. A. Oliver. 2017. “How Much Does Love Really Hurt? Psychopathology and Romantic Relationships: A Meta-Analysis.” *European Psychiatry* 41: S696.

Morriss, R., M. A. Faizal, A. P. Jones, P. R. Williamson, C. A. Bolton, and J. P. McCarthy. 2007. “Interventions for Helping People Recognise Early Signs of Recurrence in Bipolar Disorder.” *Cochrane Database of Systematic Reviews* (1).

Mosholder, Andrew D., and Mary Willy. 2006. “Suicidal Adverse Events in Pediatric Randomized, Controlled Clinical Trials of Antidepressant Drugs Are Associated with Active Drug Treatment: A Meta-Analysis.” *Journal of Child and Adolescent Psychopharmacology* 16 (1-2): 25–32.

Mosquera, L.. 2016. “Suicide Behavior in Childhood.” *REVISTA DE PSICOLOGIA CLINICA CON NINOS Y ADOLESCENTES* 3 (1): 9–18.

Neary, Donna. 2000. “The Effects of Adolescent and Young Adult Suicide on Family and Friends: A Meta-Analysis.” *Dissertation Abstracts International: Section B: The Sciences and Engineering* 61 (3-B): 1646.

Nielassoff, E., M. Le Floch, C. Avril, B. Gohier, P. Duverger, and E. Riquin. 2023. “Protective Factors of Suicidal Behaviors in Children and Adolescents/ Young Adults: A Literature Review.” *ARCHIVES DE PEDIATRIE* 30 (8): 607–616.

Oakley Browne, M., C. Galletly, G. Andrews, G. Malhi, G. Carter, and P. Hay. 2016. “The Royal Australian and New Zealand College of Psychiatrists Clinical Practice Guideline Project and Clinical Practice Guidelines for Anxiety Disorders, Mood Disorders, Schizophrenia and Related Disorders.” *Australian and New Zealand Journal of Psychiatry* 50: 31.

Ozinci, Z., J. Anjum, and R. Aggarwal. 2018. “Internet Addiction and Self-Harm/Suicide: How Are They Related?.” *Journal of the American Academy of Child and Adolescent Psychiatry* 57 (10): S147.

Patterson, S., and G. Leavey. 2024. “The Psychological Impact of Parental Suicide on Surviving Children: A Systematic Review.” *Archives of Disease in Childhood* 109: A347.

Peixoto, F. S. D., D. F. de Sousa, Dcrp Luz, N. B. Vieira, J. Goncalves, G. C. A. dos Santos, F. C. T. da Silva, and M. L. R. Neto. 2017. “Bipolarity and Suicidal Ideation in Children and Adolescents: A Systematic Review with Meta-Analysis.” *ANNALS OF GENERAL PSYCHIATRY* 16.

Plener, P. L., J. M. Fegert, M. Kaess, N. D. Kapusta, R. Brunner, R. C. Groschwitz, T. In-Albon, F. Resch, and K. Becker. 2017. “Nonsuicidal Self-Injury in Adolescence: A Clinical Guideline for Diagnostics and Therapy.” *ZEITSCHRIFT FUR KINDER-UND JUGENDPSYCHIATRIE UND PSYCHOTHERAPIE* 45 (6): 463–474.

Pu, J., X. Zhou, L. Liu, Y. Zhang, L. Yang, S. Yuan, H. Zhang, Y. Han, D. Zou, and P. Xie. 2017. “Efficacy and Acceptability of Interpersonal Psychotherapy for Depression in Adolescents: A Meta-Analysis of Randomized Controlled Trials.” *Psychiatry Research* 253: 226–232.

Pérez Arqueros, V., B. Ibáñez-Beroiz, A. Goñi-Sarriés, and A. Galbete Jiménez. 2023. “Efficacy of Psychotherapeutic Interventions for Non-Suicidal Self-Injury in Adolescent Population: Systematic Review and Meta-Analysis.” *Span J Psychiatry Ment Health* 16 (2): 119–126.

Qin, B., Y. Zhang, X. Zhou, P. Cheng, Y. Liu, J. Chen, Y. Fu, Q. Luo, and P. Xie. 2014. “Selective Serotonin Reuptake Inhibitors Versus Tricyclic Antidepressants in Young Patients: A Meta-Analysis of Efficacy and Acceptability.” *Clin Ther* 36 (7): 1087–1095.e4.

Rahman, F., R. T. Webb, and A. Wittkowski. 2021. “Risk Factors for Self-Harm Repetition in Adolescents: A Systematic Review.” *Clin Psychol Rev* 88: 102048.

Renaud-Charest, O., A. Stoljar Gold, E. Mok, J. Kichler, P. Li, and M. Nakhla. 2023. “Suicidal Ideation, Suicide Attempts, and Suicide Deaths in Adolescents and Young Adults with Type 1 Diabetes: A Systematic Review and Meta-Analysis.” *Canadian Journal of Diabetes* 47 (7): S113.

Reyes, Madeleine, and Charles P. Buscemi. 2020. “157 Systematic Review: An Educational Strategy to Improve Medication Compliance and Decrease Hospital Readmission Among Adolescents with Bipolar Disorder.” *CNS Spectrums: The International Journal of Neuropsychiatric Medicine* 25 (2): 300–301.

Richard-Devantoy, S., Y. Ding, G. Turecki, and F. Jollant. 2016. “Attentional Bias Toward Suicide-Relevant Information in Suicide Attempters: A Cross-Sectional Study and a Meta-Analysis.” *Journal of Affective Disorders* 196: 101–108.

Rotenstein, L. S., M. A. Ramos, M. Torre, J. Bradley Segal, M. J. Peluso, C. Guille, S. Sen, and D. A. Mata. 2016. “Prevalence of Depression, Depressive Symptoms, and Suicidal Ideation Among Medical Students a Systematic Review and Meta-Analysis.” *JAMA - Journal of the American Medical Association* 316 (21): 2214–2236.

Sahle, B. W., N. J. Reavley, W. Li, A. J. Morgan, M. B. H. Yap, A. Reupert, and A. F. Jorm. 2022. “The Association Between Adverse Childhood Experiences and Common Mental Disorders and Suicidality: An Umbrella Review of Systematic Reviews and Meta-Analyses.” *European Child & Adolescent Psychiatry* 31 (10): 1489–1499.

Sanchez, Maria Gonzalez, Pedro Gil Madrona, Luisa Losada Puente, and Ramon Garcia Perales. 2024. “Adolescent Suicide Prevention Programs: A Systematic Review.” *Programas de prevencion del suicidio en adolescentes: Una revision sistematica.* 17 (1): 1–28.

Schulte-Frankenfeld, P. M., J. J. F. Breedvelt, M. E. Brouwer, N. van der Spek, G. Bosmans, and C. L. Bockting. 2024. “Effectiveness of Attachment-Based Family Therapy for Suicidal Adolescents and Young Adults: A Systematic Review and Meta-Analysis.” *Clin Psychol Eur* 6 (4): e13717.

Sheldon, E., M. Simmonds-Buckley, C. Bone, T. Mascarenhas, N. Chan, M. Wincott, H. Gleeson, K. Sow, D. Hind, and M. Barkham. 2021. “Prevalence and Risk Factors for Mental Health Problems in University Undergraduate Students: A Systematic Review with Meta-Analysis.” *Journal of Affective Disorders* 287: 282–292.

Shooshtari, Mitra Hakim, and Hamid Khanipour. 2014. “Comparison of Self-Harm and Suicide Attempt in Adolescents: A Systematic Review.” *Iranian Journal of Psychiatry and Clinical Psychology* 20 (1): 3–13.

Shropshire, Ali M., and Kathy Thornton. 2011. “Prevention Measures for Adolescent Suicide: An Evidence-Based Review.” *American Journal for Nurse Practitioners* 15 (5/6): 30–36.

Sideli, L., H. Quigley, C. La Cascia, and R. M. Murray. 2020. “Cannabis Use and the Risk for Psychosis and Affective Disorders.” *Journal of Dual Diagnosis* 16 (1): 22–42.

Siu, Andrew M. H.. 2019. “Self-Harm and Suicide Among Children and Adolescents in Hong Kong: A Review of Prevalence, Risk Factors, and Prevention Strategies.” *Journal of Adolescent Health* 64 (6, Suppl): S59–S64.

Soares-Weiser, K., Y. B. Vergel, S. Beynon, G. Dunn, M. Barbieri, S. Duffy, J. Geddes, S. Gilbody, S. Palmer, and N. Woolacott. 2007. “A Systematic Review and Economic Model of the Clinical Effectiveness and Cost-Effectiveness of Interventions for Preventing Relapse in People with Bipolar Disorder.” *Health Technology Assessment* 11 (40): iii–119.

Soto-Sanz, V., P. Castellví, J. A. Piqueras, J. Rodríguez-Marín, T. Rodríguez-Jiménez, A. Miranda-Mendizábal, O. Parés-Badell, J. Almenara, I. Alonso, M. J. Blasco, A. Cebrià, A. Gabilondo, M. Gili, C. Lagares, M. Roca, and J. Alonso. 2019. “Internalizing and Externalizing Symptoms and Suicidal Behaviour in Young People: A Systematic Review and Meta-Analysis of Longitudinal Studies.” *Acta Psychiatrica Scandinavica* 140 (1): 5–19.

Souza, L. C., L. P. Galvão, H. S. Paiva, C. de Azevedo-Marques Périco, A. Ventriglio, J. Torales, J. M. Castaldelli-Maia, and A. S. Martins-Da-Silva. 2023. “Major Depressive Disorder as a Risk Factor for Suicidal Ideation for Attendees of Educational Institutions: A Meta-Analysis and Meta-Regression.” *Revista Paulista de Pediatria* 41.

Speckens, Anne E. M., and Keith Hawton. 2005. “Social Problem Solving in Adolescents with Suicidal Behavior: A Systematic Review.” *Suicide and Life-Threatening Behavior* 35 (4): 365–387.

Stanley, I. H., J. W. Boffa, M. L. Rogers, M. A. Hom, B. J. Albanese, C. Chu, D. W. Capron, N. B. Schmidt, and T. E. Joiner. 2018. “Anxiety Sensitivity and Suicidal Ideation/Suicide Risk: A Meta-Analysis.” *J Consult Clin Psychol* 86 (11): 946–960.

Stocchero, B. A., L. M. Rothmann, E. T. Portolan, T. G. Lopes, C. Ferraz-Rodrigues, M. G. Garcia, J. C. de Magalhães Narvaez, R. Grassi-Oliveira, and T. W. Viola. 2024. “The Consequences of Childhood Maltreatment on Dual-Diagnosis Psychiatric Conditions and Clinical Outcomes in Substance Use Disorders: A Systematic Review and Meta-Analysis.” *Child Abuse Negl* 158: 107085.

Suárez-Brito, P., C. Hausmann-Stabile, L. Polanco-Roman, A. Meca, and C. T. Ebrahimi. 2024. “The Role of Acculturation and Acculturation Stress in Suicidal Behaviors Among Latina Adolescents: A Systematic Review of Research.” *CHILDREN AND YOUTH SERVICES REVIEW* 163.

Tom, K., R. Shafran, D. Hargreaves, L. Muschialli, D. Linton, and S. Bennett. 2024. “Mental Health Clinical Pathways for Children and Young People with Long-Term Health Conditions: A Systematic Review.” *Journal of Psychosomatic Research* 181.

Tsapakis, E. M., F. Soldani, L. Tondo, and R. J. Baldessarini. 2008. “Efficacy of Antidepressants in Juvenile Depression: Meta-Analysis.” *Br J Psychiatry* 193 (1): 10–17.

Vaudreuil, C.. 2019. “45.4 Subthreshold Pediatric Bipolar Disorder Is a Highly Impairing Condition: A Systematic Literature Review and Meta-Analysis.” *Journal of the American Academy of Child and Adolescent Psychiatry* 58 (10): S368.

Velasco, A., A. Lengvenyte, J. Rodriguez-Revuelta, L. Jimenez-Treviño, P. Courtet, M. P. Garcia-Portilla, J. Bobes, and P. A. Sáiz. 2023. “Neutrophil-to-Lymphocyte Ratio, Platelet-to-Lymphocyte Ratio, and Monocyte-to-Lymphocyte Ratio in Depressed Patients with Suicidal Behavior: A Systematic Review.” *European Psychiatry* 67 (1).

Victor, S. E., and E. D. Klonsky. 2014. “Correlates of Suicide Attempts Among Self-Injurers: A Meta-Analysis.” *CLINICAL PSYCHOLOGY REVIEW* 34 (4): 282–297.

von Sydow, Kirsten, Stefan Beher, Jochen Schweitzer-Rothers, and Rudiger Retzlaff. 2006. “Systemic Family Therapy with Children and Adolescents as Index Patients. a Meta-Content Analysis of 47 Randomized Controlled Outcome Studies.” *Systemische Familientherapie bei Storungen des Kindesund Jugendalters. Eine Metainhaltsanalyse von 47 randomisierten Primarstudien.* 51 (2): 107–143.

Wang, M., Y. Zuo, Y. Tang, R. Zhang, Q. Lu, and B. Wang. 2025. “Anxiety and Depression Symptoms of Adolescents with Non-Suicidal Self-Injury: A Network Analysis Study.” *Journal of advanced nursing* .

Waraan, Luxsiya, Johan Siqveland, Ketil Hanssen-Bauer, Nikolai O. Czjakowski, Brynhildur Axelsdóttir, Lars Mehlum, and Marianne Aalberg. 2023. “Family Therapy for Adolescents with Depression and Suicidal Ideation: A Systematic Review and Meta–analysis.” *Clinical Child Psychology & Psychiatry* 28 (2): 831–849.

Wohlfarth, Tamar D., Barbara J. van Zwieten, Frits J. Lekkerkerker, Christine C. Gispen-de Wied, Jerry R. Ruis, Andre J. A. Elferink, and Jitschak G. Storosum. 2006. “Antidepressants Use in Children and Adolescents and the Risk of Suicide.” *European Neuropsychopharmacology* 16 (2): 79–83.

Wyatt, L. C., T. Ung, R. Park, S. C. Kwon, and C. Trinh-Shevrin. 2015. “Risk Factors of Suicide and Depression Among Asian American, Native Hawaiian, and Pacific Islander Youth: A Systematic Literature Review.” *J Health Care Poor Underserved* 26 (2 Suppl): 191–237.

Yiu, H. W., S. Rowe, and L. Wood. 2021. “A Systematic Review and Meta-Analysis of Psychosocial Interventions Aiming to Reduce Risks of Suicide and Self-Harm in Psychiatric Inpatients.” *Psychiatry Research* 305.

Yu, Minghua, Munila Muhetaer, Zepu Li, Jiaqi Zhu, Fernando Romero Menjivar, and Jiaqi Zhou. 2026. “Substance Use and Non-Suicidal Self-Injury Among Adolescents: A Meta-Analysis of Association Patterns and Moderating Factors.” *Journal of Youth & Adolescence* 55 (1): 27–45.

Zhou, X., T. Teng, Y. Zhang, C. Del Giovane, T. A. Furukawa, J. R. Weisz, X. Li, P. Cuijpers, D. Coghill, Y. Xiang, S. E. Hetrick, S. Leucht, M. Qin, J. Barth, A. V. Ravindran, L. Yang, J. Curry, L. Fan, S. G. Silva, A. Cipriani, and P. Xie. 2020. “Comparative Efficacy and Acceptability of Antidepressants, Psychotherapies, and Their Combination for Acute Treatment of Children and Adolescents with Depressive Disorder: A Systematic Review and Network Meta-Analysis.” *Lancet Psychiatry* 7 (7): 581–601.

Zuriaga, A., M. S. Kaplan, N. G. Choi, A. Hodkinson, D. Storman, N. I. Brudasca, S. P. Hirani, and S. Brini. 2021. “Association of Mental Disorders with Firearm Suicides: A Systematic Review with Meta-Analyses of Observational Studies in the United States.” *Journal of Affective Disorders* 291: 384–399.

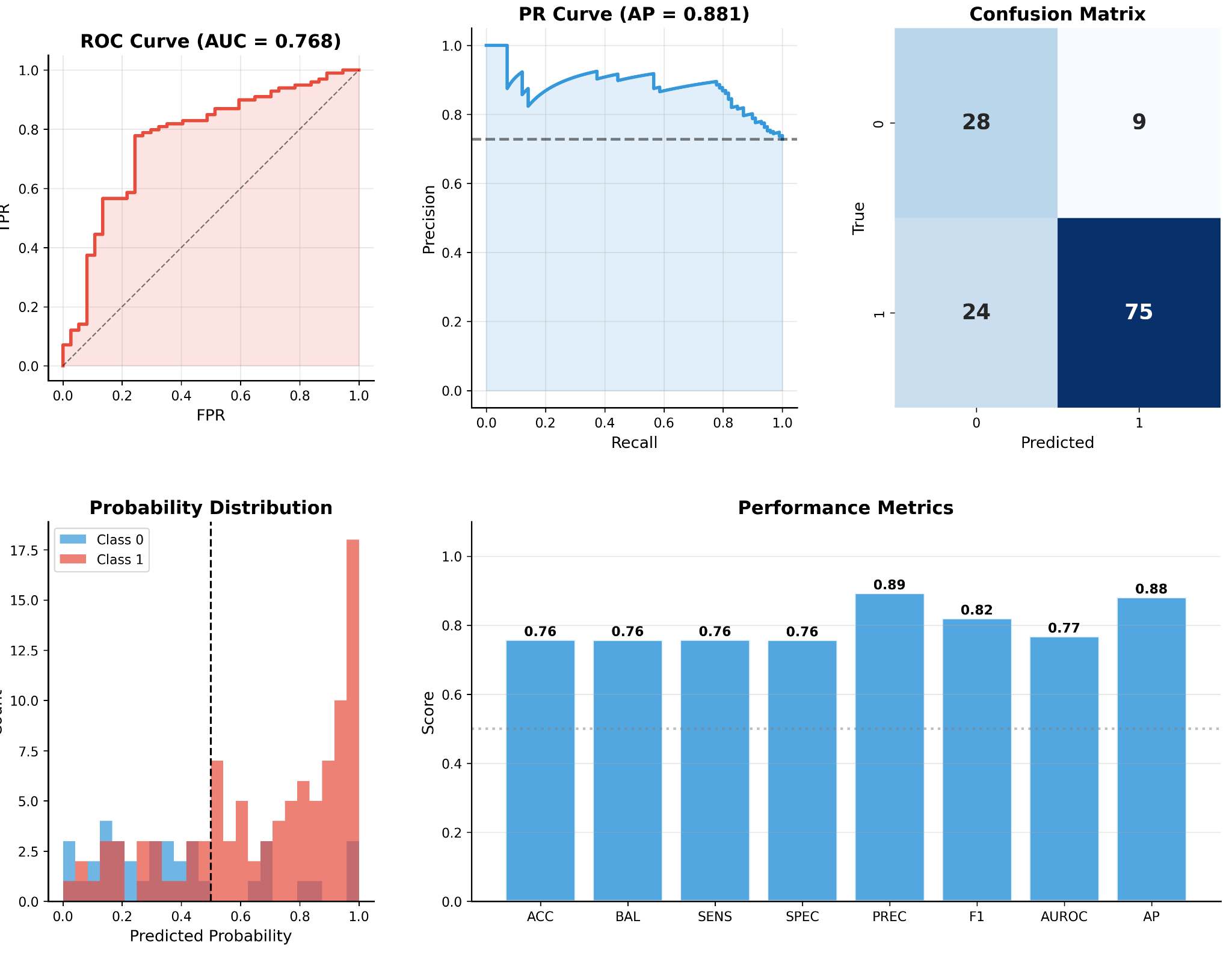

**Supplementary Figure 1.** Performance dashboard for the Evidence-Based AI LASSO model predicting suicidal ideation (MINI-KID item B1a) using leave-one-out cross-validation (N=136). The receiver operating characteristic (ROC) curve (top left) shows discrimination across all classification thresholds, with an area under the curve (AUROC) of 0.768 (95% CI 0.672 to 0.857). The curve rises steeply at low false positive rates, indicating that at conservative thresholds the model can identify a substantial fraction of positive cases while keeping the false positive rate relatively low. The precision-recall (PR) curve (top middle) is particularly informative given the class imbalance in this sample (99 positive, 37 negative). Average precision was 0.881 (95% CI 0.809 to 0.944), indicating that the model maintained high positive predictive value across most recall levels. The PR curve is a more sensitive evaluation metric than AUROC in imbalanced settings because it focuses on the model's ability to correctly identify the majority class without inflating performance estimates through the large number of true negatives.

The confusion matrix (top right) shows the classification outcome at a probability threshold of 0.5. The model correctly classified 75 of 99 participants reporting suicidal ideation (true positives) and 28 of 37 participants not reporting suicidal ideation (true negatives), yielding 9 false positives and 24 false negatives. The resulting sensitivity (0.758) and specificity (0.757) were closely balanced, which is noteworthy in the context of a 2.7-to-1 class ratio where models often achieve high sensitivity at the expense of specificity or vice versa.

The predicted probability distribution (bottom left) shows the histograms of model-assigned probabilities separated by true class membership. Participants who reported suicidal ideation (Class 1, red) were concentrated at higher predicted probabilities, with a pronounced peak near 1.0, while participants without suicidal ideation (Class 0, blue) were more diffusely distributed at lower probabilities. The overlap region between approximately 0.3 and 0.6 corresponds to the zone of greatest classification uncertainty and accounts for most of the observed misclassifications. The dashed vertical line at 0.5 indicates the classification threshold used throughout the analysis. The performance metrics summary (bottom right) shows all primary evaluation metrics at the 0.5 threshold. Precision (positive predictive value) was the highest individual metric at 0.893, indicating that when the model predicted suicidal ideation, it was correct in approximately 9 of 10 cases. The corresponding negative predictive value was 0.538 (not shown in this panel but reported in the main text), reflecting the difficulty of confidently ruling out suicidal ideation in this sample given the high base rate. All threshold-dependent metrics exceeded 0.75, and both threshold-independent metrics (AUROC and average precision) exceeded 0.76 and 0.88, respectively.

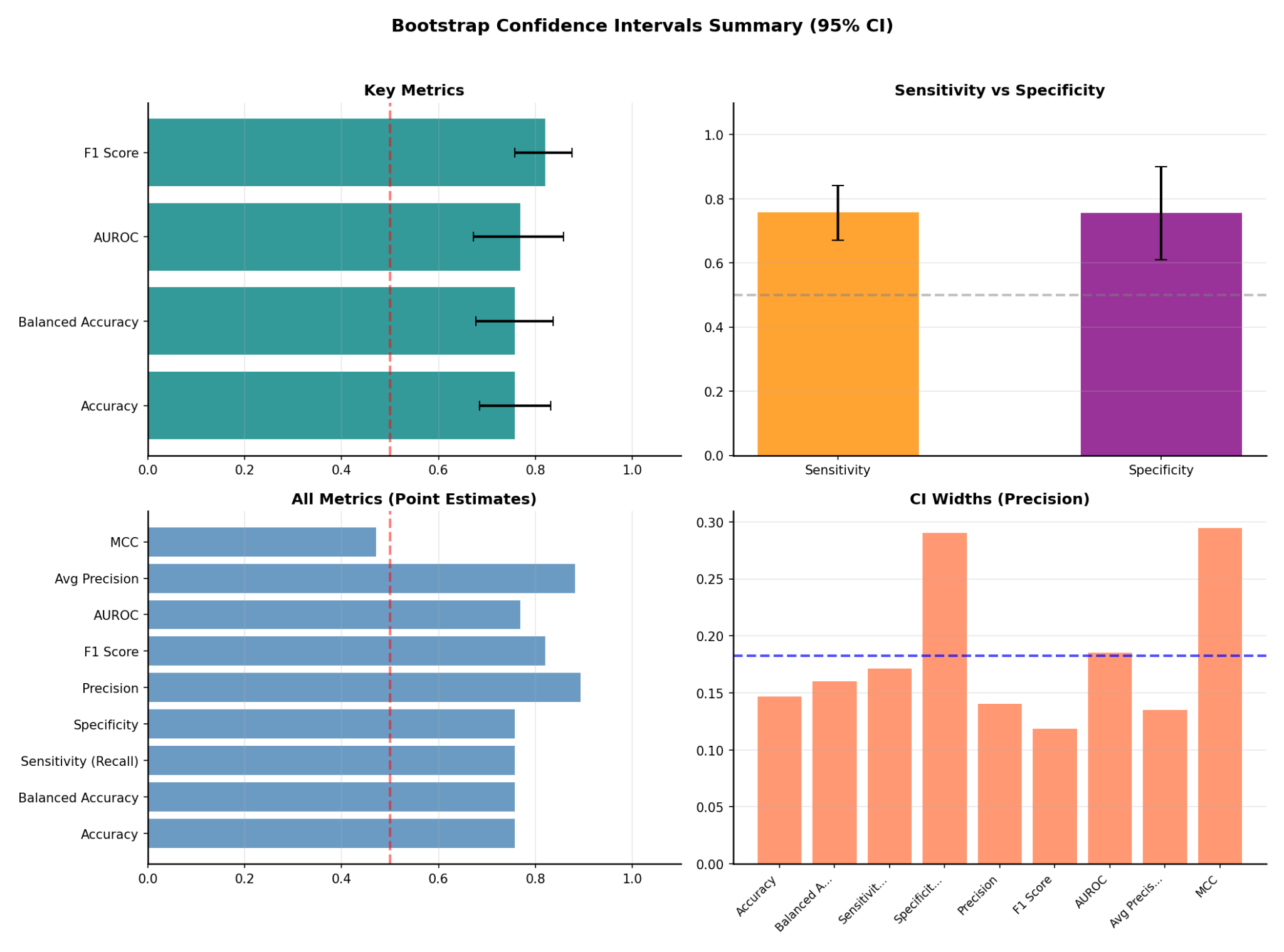

Supplementary Figure 2. Bootstrap confidence interval summary for the Evidence-Based AI LASSO model predicting suicidal ideation (MINI-KID item B1a, N=136). Confidence intervals were computed using 1,000 percentile bootstrap resamples of the 136 leave-one-out predicted probabilities.

The key metrics panel (top left) displays horizontal bars representing point estimates for F1 score, AUROC, balanced accuracy, and accuracy, with error bars indicating the corresponding 95% bootstrap confidence intervals. All four metrics exceeded the chance threshold of 0.5 (dashed vertical line), and the lower bound of each interval remained above 0.6, confirming that the model's above-chance performance is robust to sampling variability.

The sensitivity and specificity comparison (top right) shows that both metrics were closely matched at approximately 0.76, with overlapping confidence intervals. This balance is notable given the 2.7-to-1 class imbalance in the sample and indicates that the model does not achieve its overall accuracy by preferentially classifying participants into the majority class.

The full metrics panel (bottom left) displays point estimates for all nine evaluation metrics. Precision (positive predictive value, 0.893) and average precision (0.881) were the highest, reflecting the model's reliability when predicting suicidal ideation. Matthews correlation coefficient (0.471) was the lowest primary metric, which is expected given that MCC penalizes imbalance between false positive and false negative rates more heavily than accuracy-based measures.

The confidence interval width panel (bottom right) shows the precision of each bootstrap estimate, with narrower bars indicating more stable estimates. F1 score (width 0.118) and average precision (0.135) produced the narrowest intervals, consistent with their dependence on the larger positive class (n=99). Specificity (0.290) and MCC (0.295) produced the widest intervals, reflecting the smaller negative class (n=37) and the sensitivity of composite metrics to cell counts in the confusion matrix. The mean interval width across all metrics was 0.182 (dashed horizontal line).

**Supplementary Methods S1. Structured prompt template for penalty factor scoring**

The penalty scoring stage of the EBAL pipeline requires the language model to evaluate each candidate predictor and assign a numerical weight reflecting its evidence-based relevance to the outcome of interest. Because this judgment directly determines which predictors the LASSO model will favor or penalize during estimation, the instructions given to the language model must be precise, reproducible, and constrained against uncontrolled reasoning. The full prompt template used for penalty scoring in this study is reproduced below and is publicly available at <https://github.com/bmwas/LLM-Lasso/blob/main/prompts/pbd/pbd_normal.txt>.

The prompt was designed around three principles. The first was determinism. The model was instructed to assign integer penalty factors on a fixed ordinal scale from 2 (strongest relevance to suicidal ideation) to 5 (weakest or ambiguous relevance), with each tier defined by explicit criteria rather than left to the model's discretion. This integer scale was chosen to produce discrete, auditable penalty assignments that could be directly mapped to the weighted LASSO penalty structure, as opposed to continuous scores that would require additional calibration.

The second principle was input restriction. The prompt included a hard constraint requiring the model to base its scoring on only two sources of information, namely the predictor name and the retrieved evidence passages supplied through the retrieval-augmented generation system. The model was explicitly prohibited from citing external literature, inferring associations not present in the predictor name, assuming directionality of effects, or supplementing the provided context with prior clinical knowledge from its training data. This constraint was critical because the purpose of the pipeline is to channel curated, methodologically screened evidence into the penalty structure. Allowing the model to draw on unfiltered pretraining knowledge would undermine the quality control that the upstream evidence synthesis was designed to provide.

The third principle was transparency. The prompt required the model to produce a one-sentence rationale grounded in the predictor name for each penalty assignment, creating a per-variable audit trail that can be reviewed by domain experts. This requirement also served as a behavioral guardrail. By forcing the model to articulate a name-grounded justification, the prompt reduced the likelihood that the model would assign penalties based on latent associations from its training corpus that could not be traced to the curated evidence base.

The template contains a single placeholder variable, {factors}, which was populated at runtime with the list of 20 candidate predictor names. All other elements of the prompt, including the clinical context, the scoring rubric, and the output format, remained fixed across all scoring queries. The 20 predictors were processed in two sequential batches of 10 due to context window management, with the same prompt template applied to each batch.

We have a pediatric clinical dataset collected from children and adolescents diagnosed with pediatric bipolar disorder, with common comorbidities including ADHD and anxiety disorders (e.g., GAD, specific phobia, social anxiety), and depressive spectrum conditions (e.g., major depression, dysthymia). Our goal is to build a statistical Lasso (L1-regularized) classification model to predict whether a participant has **suicidal ideation** (SI+) versus **no suicidal ideation** (SI−), based on the **provided** clinical and psychosocial predictors.

\### HARD CONSTRAINT: USE ONLY PROVIDED CONTEXT (NO SPECULATION / NO EXTERNAL FACTS)

You MUST use **only**:

1) the information in this prompt, and

2) the predictor names provided in `{factors}`.

You must NOT:

- cite or rely on external literature, “known risk factors,” or general clinical evidence beyond what is explicitly stated here,

- infer what a predictor measures beyond what its name directly indicates,

- assume directionality (risk vs protective) unless explicit in the predictor name,

- add variables, mechanisms, or associations not present in `{factors}`,

- “fill in” missing details with guesses.

If a predictor name is ambiguous or not self-explanatory, treat it as **low-evidence** and penalize it more.

---

\## Task

Provide **penalty factors** (feature-specific Lasso weights) for each predictor provided. These penalty factors must be **integers between 2 and 5 (inclusive)**, where:

- **2** indicates the predictor name **explicitly** signals strong, direct relevance to suicidal ideation classification (e.g., the name contains terms like suicid*, suicidal ideation, self-harm, attempt, NSSI).

- **5** indicates the predictor name is unclear/ambiguous OR suggests minimal/indirect relevance based on the name alone.

The penalty factors must be listed in the **exact same order** as the predictors provided in `{factors}`.

---

\## Name-Only Scoring Rules (apply strictly)

Assign penalties using ONLY the predictor name:

- **Penalty = 2** (High specificity)

Predictor name explicitly references SI or self-harm concepts (e.g., suicidal ideation, suicid*, self-harm, attempt, NSSI).

- **Penalty = 3** (Moderate specificity)

Predictor name explicitly references severe mood/affective constructs in the prompt’s diagnostic scope (e.g., depression severity, dysthymia symptoms, bipolar episode severity) AND is clearly interpretable from the name alone.

- **Penalty = 4** (Low specificity)

Predictor name references broader clinical/psychosocial constructs in scope (e.g., anxiety, ADHD symptoms, functioning, sleep) but does not explicitly reference SI/self-harm.

- **Penalty = 5** (Unclear / ambiguous / distal)

Predictor name is ambiguous, administrative, demographic, unclear, or not interpretable without additional definitions.

---

\## Output Requirements

1) Preserve the **exact order** of predictors in `{factors}`.

2) For each predictor, output:

- Penalty (2–5)

- A brief rationale (≤ 1 sentence) grounded ONLY in the predictor name (no external claims).

**Output format (repeat for each predictor, in order):**

Predictor: <name> | Penalty: <2-5> | Rationale: <1 sentence max, name-grounded>

Do not include meta-commentary or disclaimers.

The list of predictors is: {factors}

**Supplementary Methods S2. Language model reasoning trace for penalty factor assignment**

A central design principle of the EBAL analytical pipeline is that the language model’s role in assigning penalty factors should be transparent and auditable rather than opaque. To support this, we provide the complete reasoning trace generated by the Qwen3-30B-A3B-Thinking model during penalty scoring of the 20 candidate predictors for the suicidal ideation outcome. This trace captures the model’s internal chain-of-thought reasoning in full, exactly as it was produced during the single deterministic scoring pass used in the analysis.

The reasoning model was instructed through a structured prompt (see above) to assign an integer penalty factor on a scale from 2 to 5 for each candidate predictor, where lower penalties correspond to stronger evidence-based relevance to suicidal ideation and higher penalties correspond to weaker or absent relevance. At each step, the model received the predictor name and the three most semantically relevant text segments retrieved from the curated evidence corpus, then evaluated the predictor against the scoring rubric before assigning a penalty. The rubric required the model to distinguish among four tiers of relevance. Predictors whose names explicitly referenced suicidal ideation or self-harm received a score of 2. Predictors referencing severe mood or affective constructs within the diagnostic scope (such as depressed or manic mood) received a score of 3. Predictors referencing broader clinical or psychosocial constructs (such as anxiety comorbidity or ADHD) received a score of 4. Predictors with demographic, administrative, or ambiguous names received a score of 5.

Several features of the reasoning trace are worth noting. First, the model systematically evaluated each scoring tier before arriving at a penalty, rather than producing scores without justification. For example, when scoring “current_mood_euthymic,” the model recognized that euthymic mood is a mood state but not a severe one, and explicitly distinguished it from depressed and manic mood before assigning a penalty of 4 rather than 3. Second, the model correctly declined to use clinical knowledge beyond the predictor name and retrieved evidence when instructed to do so. When scoring “currently_taking_lithium”, the model acknowledged the known suicide protective properties of lithium from the retrieved evidence but assigned a penalty of 5 because the predictor name itself did not reference suicidal ideation, consistent with the scoring rubric. Third, the model identified genuinely ambiguous predictors (such as “bis_bas_bis_parent” and “bis_bas_reward_parent”) and penalized them accordingly, noting that the names were not self-explanatory without additional context. These behaviors illustrate that the model functioned as a structured translator between the scoring rubric and the predictor list rather than as an unconstrained clinical judge.

The full trace is reproduced below without editing or reformatting. Predictors were processed in two sequential batches of 10.

================================================================================

LLM PENALTY FACTOR REASONING LOG

================================================================================

Generated: 2026-01-31 14:02:23

LLM Backend: vllm

Model: qwen3-thinking

Category: Suicidal Ideation

Features: 20

Prompt file: prompts/pbd/pbd_normal.txt

================================================================================

PENALTY FACTOR SUMMARY

------------------------------------------------------------

Feature	Penalty

------------------------------------------------------------

years_of_education	5.0

current_mood_depressed	3.0

current_mood_euthymic	4.0

current_mood_manic	3.0

lenght_illness_mood_disorder	4.0

comorbidities_generalized_anxiety_disorder	4.0

comorbidities_personality_disorder	4.0

comorbidities_agoraphobia	4.0

comorbidities_adhd	4.0

currently_taking_antidepressant	4.0

currently_taking_lithium	5.0

total_number_of_hospitalizations	5.0

zero_hospitalizations_due_to_mania	5.0

father_education_less_than_12_years	5.0

obesity	4.0

severe_obesity	4.0

hispanic_or_latino	5.0

bis_bas_bis_parent	5.0

bis_bas_reward_parent	5.0

birthweight_less_than_5.5_pounds	5.0

------------------------------------------------------------

Mean	4.35

Min	3.00

Max	5.00

Std	0.65

================================================================================

DETAILED LLM REASONING

================================================================================

--- BATCH 1 ---

We are given a list of predictors and must assign penalty factors (2-5) based solely on the predictor names, using the provided scoring rules.

Let's go through each predictor in the exact order:

1. `years_of_education`

- This is a demographic variable (education level). It does not reference any mental health condition, suicidal ideation, or related concepts.

- According to the rules: "Predictor name is ambiguous, administrative, demographic, unclear, or not interpretable without additional definitions." -> Penalty 5.

- Rationale: The name refers to a demographic factor without any direct link to suicidal ideation.

2. `current_mood_depressed`

- This name explicitly references "depressed" mood, which is a severe mood/affective construct. The prompt states that the dataset includes depressive spectrum conditions (e.g., major depression, dysthymia). The name directly indicates a current depressed mood state, which is a known risk factor for suicidal ideation.

- According to the rules: "Predictor name explicitly references severe mood/affective constructs in the prompt’s diagnostic scope" -> Penalty 3.

- Rationale: The name directly indicates a current depressed mood state, a known risk factor for suicidal ideation.

3. `current_mood_euthymic`

- This name refers to "euthymic" mood, which means a normal, stable mood (not depressed or manic). While it is a mood state, it is not a severe mood disorder state. The prompt does not indicate that euthymic mood is a risk factor for suicidal ideation (in fact, it's the opposite). However, the rule for penalty 3 requires "explicitly references severe mood/affective constructs". Euthymic is not severe; it's the absence of severe mood symptoms. Therefore, it does not meet the criteria for penalty 3.

- The rule for penalty 4: "Predictor name references broader clinical/psychosocial constructs in scope (e.g., anxiety, ADHD symptoms, functioning, sleep) but does not explicitly reference SI/self-harm." Euthymic is a mood state, but it's not a severe one. However, note that the rule for penalty 3 requires "severe mood/affective constructs". Since euthymic is not severe, it doesn't qualify for penalty 3. It is a mood state but not a severe one, so it falls under penalty 4 (low specificity) because it's a broader clinical construct (mood state) without explicit reference to SI or severe mood.

- Rationale: The name refers to a stable, non-depressed mood state, which is not a direct risk factor for suicidal ideation.

4. `current_mood_manic`

- This name explicitly references "manic" mood, which is a severe mood/affective construct (part of bipolar disorder). The prompt states the dataset is for pediatric bipolar disorder, and manic episodes are a core feature. Mania is associated with increased risk of suicidal behavior in some contexts (though complex), but the rule for penalty 3 is for "severe mood/affective constructs" and the name directly states "manic", which is a severe mood state.

- Rationale: The name directly indicates a current manic mood state, a severe mood disorder symptom in bipolar disorder.

5. `lenght_illness_mood_disorder`

- Note: There is a typo in the predictor name (should be "length" but written as "lenght"). The name refers to the "length of illness" for mood disorder. This is a duration measure, which is a clinical construct but not directly related to suicidal ideation. The prompt does not state that illness duration is a known risk factor for SI. It is a broader clinical construct (illness duration) without explicit reference to SI or severe mood.

- According to the rules: "Predictor name references broader clinical/psychosocial constructs in scope" -> Penalty 4.

- Rationale: The name refers to the duration of mood disorder, a clinical measure not directly linked to suicidal ideation in the name.

6. `comorbidities_generalized_anxiety_disorder`

- This name explicitly lists a comorbidity: generalized anxiety disorder (GAD). The prompt states that common comorbidities include anxiety disorders (e.g., GAD). However, the name does not explicitly reference suicidal ideation. The rule for penalty 3 requires "explicitly references severe mood/affective constructs" (which anxiety is, but note: the rule for penalty 3 is for mood/affective constructs, and anxiety is a separate category). The scoring rules state for penalty 3: "severe mood/affective constructs" (like depression, dysthymia, bipolar). Anxiety is a different category, but the prompt includes anxiety disorders as comorbidities. However, the rule for penalty 3 is specifically for mood/affective constructs. Anxiety is not mood (it's an anxiety disorder). Therefore, it does not qualify for penalty 3. It falls under penalty 4 because it's a broader clinical construct (anxiety disorder comorbidity) without explicit SI reference.

- Rationale: The name indicates a comorbidity of generalized anxiety disorder, an anxiety disorder not explicitly linked to suicidal ideation in the name.

7. `comorbidities_personality_disorder`

- This name refers to comorbid personality disorder. The prompt states that common comorbidities include personality disorders (as per the context: "comorbidities including ... personality disorders" is not explicitly stated in the prompt, but the predictor name includes it). However, the rule for penalty 3 requires "severe mood/affective constructs", and personality disorder is not a mood disorder. It is a broader clinical construct. The context document 14 mentions personality disorders (borderline, antisocial, avoidant) as increasing suicide risk, but the scoring rule says we must use only the predictor name. The name does not say "suicidal" or "mood", so it doesn't qualify for penalty 3. It is a comorbidity, but not a mood disorder. Therefore, penalty 4.

- Rationale: The name refers to a comorbidity of personality disorder, a clinical condition not explicitly linked to suicidal ideation in the name.

8. `comorbidities_agoraphobia`

- This name refers to comorbidity of agoraphobia (a specific anxiety disorder). Similar to the previous, it is an anxiety disorder comorbidity. The name does not reference suicidal ideation or severe mood. It is a specific anxiety disorder, so it falls under penalty 4 (broader clinical construct).

- Rationale: The name indicates a comorbidity of agoraphobia, an anxiety disorder not explicitly linked to suicidal ideation in the name.

9. `comorbidities_adhd`

- This name refers to comorbidity of ADHD. The prompt states that common comorbidities include ADHD. The name does not reference suicidal ideation. ADHD is a neurodevelopmental disorder, not a mood disorder. The context document 16 and 17 mention ADHD and suicide, but the scoring rule says we must use only the predictor name. The name does not contain any term related to SI or mood, so it is a broader clinical construct (ADHD comorbidity) -> penalty 4.

- Rationale: The name indicates a comorbidity of ADHD, a neurodevelopmental disorder not explicitly linked to suicidal ideation in the name.

10. `currently_taking_antidepressant`

- This name refers to current use of antidepressants. The prompt does not state that antidepressant use is a risk factor for SI (in fact, it's complex: antidepressants can have both risk and protective effects). However, the name does not explicitly reference suicidal ideation. The rule for penalty 3 is for mood/affective constructs, but antidepressant use is a treatment, not a mood state. It is a clinical intervention. The context document 19 and 20 mention antidepressants and suicidal behavior, but we cannot use that because the rule says "only the predictor name". The name does not contain "suicid" or "ideation", so it does not qualify for penalty 2 or 3. It is a broader clinical construct (treatment) -> penalty 4.

- Rationale: The name refers to current antidepressant use, a treatment factor not explicitly linked to suicidal ideation in the name.

Now, let's assign the penalties:

1. years_of_education -> 5 (demographic, not related to SI)

2. current_mood_depressed -> 3 (explicitly severe mood: depressed)

3. current_mood_euthymic -> 4 (mood state, but not severe; euthymic is the opposite of depressed, so not a risk factor and not severe)

4. current_mood_manic -> 3 (explicitly severe mood: manic)

5. lenght_illness_mood_disorder -> 4 (illness duration, broader clinical construct)

6. comorbidities_generalized_anxiety_disorder -> 4 (anxiety comorbidity, not mood)

7. comorbidities_personality_disorder -> 4 (personality disorder comorbidity, not mood)

8. comorbidities_agoraphobia -> 4 (anxiety comorbidity, not mood)

9. comorbidities_adhd -> 4 (ADHD comorbidity, not mood)

10. currently_taking_antidepressant -> 4 (treatment, not mood)

Note: We must be strict. The rule for penalty 3 requires "explicitly references severe mood/affective constructs". Only `current_mood_depressed` and `current_mood_manic` are severe mood states (depressed and manic). `current_mood_euthymic` is not severe (it's normal mood) so it gets penalty 4.

Let's double-check the scoring rules:

- Penalty 2: only if the name contains terms like "suicid*", "suicidal ideation", etc. None of the predictors have that.

- Penalty 3: explicitly references severe mood/affective constructs (depression, mania, etc.) -> `current_mood_depressed` and `current_mood_manic` qualify.

- Penalty 4: broader clinical/psychosocial constructs (anxiety, ADHD, etc.) -> the rest.

Therefore, the penalties are:

1. years_of_education: 5

2. current_mood_depressed: 3

3. current_mood_euthymic: 4

4. current_mood_manic: 3

5. lenght_illness_mood_disorder: 4

6. comorbidities_generalized_anxiety_disorder: 4

7. comorbidities_personality_disorder: 4

8. comorbidities_agoraphobia: 4

9. comorbidities_adhd: 4

10. currently_taking_antidepressant: 4

Now, we output in the exact order with the required format.

</think>

Predictor: years_of_education | Penalty: 5 | Rationale: The name refers to a demographic factor without any explicit link to suicidal ideation or mood disorders.

Predictor: current_mood_depressed | Penalty: 3 | Rationale: The name explicitly references a severe mood state (depressed) directly associated with suicidal ideation risk.

Predictor: current_mood_euthymic | Penalty: 4 | Rationale: The name indicates a stable mood state (euthymic), not a severe mood disorder, and lacks explicit suicidal ideation relevance.

Predictor: current_mood_manic | Penalty: 3 | Rationale: The name explicitly references a severe mood state (manic) directly associated with suicidal ideation risk in bipolar disorder.

Predictor: lenght_illness_mood_disorder | Penalty: 4 | Rationale: The name refers to illness duration, a clinical construct without explicit suicidal ideation linkage in the term.

Predictor: comorbidities_generalized_anxiety_disorder | Penalty: 4 | Rationale: The name indicates an anxiety comorbidity, a broader clinical construct not explicitly linked to suicidal ideation in the term.

Predictor: comorbidities_personality_disorder | Penalty: 4 | Rationale: The name refers to a personality disorder comorbidity, a clinical construct without explicit suicidal ideation reference.

Predictor: comorbidities_agoraphobia | Penalty: 4 | Rationale: The name specifies an anxiety disorder comorbidity (agoraphobia), not directly tied to suicidal ideation in the term.

Predictor: comorbidities_adhd | Penalty: 4 | Rationale: The name indicates an ADHD comorbidity, a neurodevelopmental condition without explicit suicidal ideation reference.

Predictor: currently_taking_antidepressant | Penalty: 4 | Rationale: The name refers to antidepressant use, a treatment factor without explicit suicidal ideation linkage in the term.

----------------------------------------

--- BATCH 2 ---

We are given a list of predictors and must assign penalty factors (2-5) based solely on the predictor names, using the provided scoring rules.

Let's go through each predictor one by one:

1. **currently_taking_lithium**

- The name "lithium" is a medication known to be used for bipolar disorder and has been studied for its antisuicidal effects (as seen in Document 3: "Lithium versus placebo" and its effect on reducing suicides). However, the scoring rule is based on the name alone. The name does not explicitly contain "suicid*", "suicidal ideation", etc. But note: the context provided in the documents (Document 3) does mention lithium's effect on suicide. However, the instruction says: "You must NOT cite or rely on external literature, 'known risk factors,' or general clinical evidence beyond what is explicitly stated here." The only explicit statement about lithium in the context is that it was studied for antisuicidal effects, but the predictor name itself does not contain the term "suicid*".

- According to the rules:

- Penalty 2: requires explicit reference to SI or self-harm concepts (e.g., "suicidal ideation").

- This name does not have that.

- It is a medication that is known in the context to be related to suicide prevention, but the name itself does not say that.

- The rule says: "assign penalties using ONLY the predictor name".

- Therefore, we cannot assume the medication's known effect.

- The name "currently_taking_lithium" is a clinical variable that might be associated with treatment, but it does not explicitly reference suicidal ideation.

- It is not a direct indicator of suicidal ideation (it's a treatment status).

- So, it does not meet the criteria for penalty 2.

- It is not a mood/affective construct (like depression severity) but a treatment.

- It is not a broader clinical construct (like anxiety) but a specific medication.

- However, note that the scoring rule for penalty 3 requires "explicitly references severe mood/affective constructs in the prompt’s diagnostic scope". The prompt's diagnostic scope includes bipolar disorder, but "lithium" is a treatment, not a mood construct.

- Therefore, it is not penalty 3.

- It is not a broader clinical/psychosocial construct (like anxiety) but a specific treatment.

- So, it falls under penalty 4 or 5?

- The rule for penalty 4: "references broader clinical/psychosocial constructs in scope (e.g., anxiety, ADHD symptoms, functioning, sleep) but does not explicitly reference SI/self-harm."

- "Lithium" is not a clinical construct in the sense of symptoms, but a treatment. It's not a symptom or a disorder.

- The rule for penalty 5: "predictor name is ambiguous, administrative, demographic, unclear, or not interpretable without additional definitions."

- The name "currently_taking_lithium" is clear: it means the patient is currently taking lithium. But it is not directly related to suicidal ideation in the name.

- However, note that the context provided in the documents (Document 3) does link lithium to suicide prevention. But we are not allowed to use that.

- Since the name does not explicitly reference SI, and it's not a mood construct (it's a treatment), and it's not a broader clinical construct (it's a specific medication), we must assign the highest penalty for being indirect?

- But note: the scoring rule for penalty 4 is for broader clinical constructs. "Lithium" is not a clinical construct (like depression) but a treatment.

- The rule for penalty 5 says "administrative, demographic, unclear, or not interpretable". This is interpretable (it's a medication status) but it's not directly related to SI.

- However, the rule says: "predictor name is ambiguous or not interpretable without additional definitions" — this one is interpretable, but it's not directly related.

- The rule for penalty 5 also includes "distal" (not directly related).

- Given that the name does not contain any term related to SI, and it's a treatment (which is not a symptom), it should be penalty 5?

- But wait: the scoring rule for penalty 4 says "broader clinical/psychosocial constructs in scope". The prompt's scope includes bipolar disorder and comorbidities, and lithium is a treatment for bipolar disorder. However, the name does not say "bipolar" or "mood disorder", it says "lithium".

- The rule for penalty 3 requires "explicitly references severe mood/affective constructs" — lithium does not reference a mood construct, it's a treatment.

- Therefore, it does not qualify for penalty 3.

- It is not a symptom (so not penalty 2 or 3) and it's not a broader clinical construct (like anxiety) but a specific treatment.

- The rule for penalty 4 is for constructs like anxiety, ADHD, etc. — which are symptoms or disorders. Lithium is not a symptom or disorder.

- So, it is not penalty 4.

- Then it must be penalty 5 because it's not directly related and the name doesn't signal relevance to SI.

- However, note that the context of the study (the prompt) is about pediatric bipolar disorder and suicidal ideation. But we are not allowed to use that context for the penalty assignment. We must use only the name.

- Therefore, penalty = 5.

But wait: let's check the exact rule for penalty 5: "predictor name is ambiguous, administrative, demographic, unclear, or not interpretable without additional definitions."

- The name is interpretable: it means the patient is taking lithium.

- However, the rule also says "or not interpretable without additional definitions" — but it is interpretable.

- But note: the rule for penalty 5 also includes "distal" (not directly related). The rule says "unclear / ambiguous / distal".

- The name is clear, but the relevance to SI is not direct.

- The scoring rules are based on the name's signal of relevance. Since the name does not contain any term related to SI, and it's a treatment (which is not a symptom), it is not a direct indicator.

- Therefore, we assign penalty 5.

2. **total_number_of_hospitalizations**

- This is a count of hospitalizations. It is a demographic/clinical administrative variable.

- It does not explicitly reference SI or self-harm.

- It is not a mood construct.

- It is a broader clinical construct? The rule for penalty 4: "broader clinical/psychosocial constructs in scope (e.g., anxiety, ADHD symptoms, functioning, sleep)".

- Hospitalizations are a clinical outcome, but they are not a symptom. They are a consequence of illness.

- However, the rule says "broader clinical constructs" — hospitalizations might be considered a clinical outcome, but the rule examples are symptoms (anxiety, ADHD symptoms).

- The name does not signal a direct link to SI.

- It is not a symptom of SI, but a general measure of illness severity.

- Therefore, it is not penalty 2 or 3.

- It is a clinical construct (in the sense of healthcare utilization) but not a symptom.

- The rule for penalty 4 includes "functioning" and "sleep", which are symptoms. Hospitalizations are not a symptom.

- So, it is not penalty 4.

- It is administrative (a count of events) and not directly related to SI.

- Therefore, penalty 5.

3. **zero_hospitalizations_due_to_mania**

- This is a binary indicator (0 or 1) for whether there were zero hospitalizations due to mania.

- It is a specific type of hospitalization (due to mania).

- The name does not contain "suicid*" or "SI".

- It is a clinical event (hospitalization due to mania) but not directly related to SI.

- It is not a mood construct (mania is a mood construct, but the name is about hospitalizations due to mania, not mania itself).

- The rule for penalty 3: "explicitly references severe mood/affective constructs" — the name does not reference mania (it references hospitalizations due to mania).

- The name does not say "mania", it says "hospitalizations due to mania".

- So, it does not explicitly reference a mood construct (it references a consequence of mania).

- Therefore, it is not penalty 3.

- It is not a symptom (it's a count of events).

- It is administrative (a count of hospitalizations).

- So, penalty 5.

4. **father_education_less_than_12_years**

- This is a demographic variable (father's education level).

- It is clearly demographic.

- The rule for penalty 5: "demographic" is explicitly mentioned.

- Therefore, penalty 5.

5. **obesity**

- This is a medical condition.

- The rule for penalty 4: "broader clinical/psychosocial constructs in scope (e.g., anxiety, ADHD symptoms, functioning, sleep)".

- Obesity is a medical condition, but it is not a symptom of mood disorder.

- However, note that in the context of the study (pediatric bipolar disorder), obesity might be a comorbidity.

- But the name does not reference SI.

- The rule for penalty 4 is for constructs that are in the scope of the prompt (which includes comorbidities like ADHD and anxiety). Obesity is a comorbidity that might be associated with mood disorders, but the name does not say that.

- The rule says: "references broader clinical/psychosocial constructs in scope" — obesity is a clinical construct (a medical condition) and is in the scope of the study (as a comorbidity).

- However, the rule examples are "anxiety, ADHD symptoms, functioning, sleep" — which are symptoms or disorders. Obesity is a physical condition.

- But note: the prompt says "common comorbidities including ADHD and anxiety disorders" — and obesity is not listed as a comorbidity in the prompt, but it is a condition that might be present.

- The rule for penalty 4 is for constructs that are in the scope (which includes comorbidities).

- However, the name "obesity" does not explicitly reference a mood disorder or SI.

- But the rule for penalty 4 does not require the construct to be directly related to SI, only that it is a broader clinical construct in scope.

- The prompt's scope includes "depressive spectrum conditions" and comorbidities. Obesity is a comorbidity that might be present.

- However, the rule says: "does not explicitly reference SI/self-harm".

- The name "obesity" does not reference SI, so it's not penalty 2.

- It is a clinical construct (a medical condition) and is in the scope of the study (as a potential comorbidity).

- Therefore, it should be penalty 4?

- But note: the rule for penalty 4 says "broader clinical/psychosocial constructs in scope (e.g., anxiety, ADHD symptoms, functioning, sleep)".

- Obesity is not listed in the examples, but it is a clinical condition.

- However, the examples are all mental health symptoms or disorders. Obesity is a physical condition.

- The prompt does not specify that obesity is a comorbidity in the study, but it is a common comorbidity in psychiatric disorders.

- But we are to judge by the name alone.

- The name "obesity" is a medical condition, and it is not a symptom of mood disorder.

- The rule for penalty 4 is for constructs that are in the scope of the prompt (which includes comorbidities).

- However, the rule does not say that the construct must be a mental health symptom. It says "broader clinical/psychosocial constructs".

- Obesity is a clinical construct.

- But note: the examples given are all mental health related (anxiety, ADHD, etc.).

- The rule says "in scope" — the prompt's scope includes comorbidities, and obesity is a comorbidity.

- Therefore, it is a broader clinical construct in scope.

- So, penalty 4.

6. **severe_obesity**

- Similar to obesity, but more severe.

- It is a more severe form of obesity.

- Same reasoning: it is a clinical construct (a medical condition) and in scope as a comorbidity.

- Therefore, penalty 4.

7. **hispanic_or_latino**

- This is a demographic variable (ethnicity).

- The rule for penalty 5: "demographic" is explicitly mentioned.

- Therefore, penalty 5.

8. **bis_bas_bis_parent**

- This is a complex name.

- "BIS" stands for Behavioral Inhibition System, and "BAS" for Behavioral Activation System.

- "BIS" is a temperament trait related to avoidance and fear, and "BAS" is related to approach and reward.

- The name "bis_bas_bis_parent" likely refers to the parent's BIS (Behavioral Inhibition System) score.

- The rule:

- Penalty 2: requires explicit reference to SI.

- This name does not have that.

- Penalty 3: requires explicit reference to severe mood/affective constructs.

- The BIS is a temperament trait that is associated with anxiety and depression, but the name does not say that.

- The name is "bis_bas_bis_parent" — it's a bit ambiguous.

- The rule says: "if a predictor name is ambiguous or not self-explanatory, treat it as low-evidence and penalize it more."

- The name "bis_bas_bis_parent" is not self-explanatory without knowing what BIS and BAS are.

- It is not a standard term that would be known to a clinician without context.

- Therefore, it is ambiguous.

- The rule for penalty 5: "ambiguous" is included.

- So, penalty 5.

9. **bis_bas_reward_parent**

- Similar to the above.

- "BAS_reward" likely refers to the reward sensitivity part of the Behavioral Activation System.

- Again, the name is not self-explanatory.

- It is ambiguous.

- Penalty 5.

10. **birthweight_less_than_5.5_pounds**

- This is a birth weight indicator (low birth weight).

- It is a demographic/medical history variable.

- The rule for penalty 5: "demographic" or "administrative".

- It is not related to SI in the name.

- Therefore, penalty 5.

Now, let's summarize:

1. currently_taking_lithium: penalty 5 (treatment status, not directly related to SI in name)

2. total_number_of_hospitalizations: penalty 5 (administrative, not directly related)

3. zero_hospitalizations_due_to_mania: penalty 5 (administrative, not directly related)

4. father_education_less_than_12_years: penalty 5 (demographic)

5. obesity: penalty 4 (broader clinical construct in scope as a comorbidity)

6. severe_obesity: penalty 4 (same as above, but more severe)

7. hispanic_or_latino: penalty 5 (demographic)

8. bis_bas_bis_parent: penalty 5 (ambiguous)

9. bis_bas_reward_parent: penalty 5 (ambiguous)

10. birthweight_less_than_5.5_pounds: penalty 5 (demographic/medical history)

But note: for obesity and severe_obesity, we assigned penalty 4. However, let's double-check the rule for penalty 4: "references broader clinical/psychosocial constructs in scope (e.g., anxiety, ADHD symptoms, functioning, sleep)".

- Obesity is not listed in the examples, but it is a clinical condition.

- The prompt says "common comorbidities including ADHD and anxiety disorders", and obesity is a common comorbidity in psychiatric disorders (though not explicitly listed).

- However, the rule says "in scope" — the scope of the study includes comorbidities, so obesity is in scope.

- But note: the examples are all mental health symptoms. Obesity is a physical condition.

- The rule does not specify that the construct must be mental health. It says "broader clinical/psychosocial constructs".

- Clinical can include physical conditions.

- Therefore, we'll keep penalty 4 for obesity and severe_obesity.

However, note that the rule for penalty 4 says "does not explicitly reference SI/self-harm". The name "obesity" does not reference SI, so it's not penalty 2. And it is a clinical construct in scope, so penalty 4.

But wait: the rule for penalty 3 is for "explicitly references severe mood/affective constructs". Obesity is not a mood construct. So it doesn't qualify for penalty 3. And it's not penalty 2. So penalty 4 is the next.

Now, let's write the output in the exact order.

Note: The rule says "if a predictor name is ambiguous or not self-explanatory, treat it as low-evidence and penalize it more." For bis_bas_bis_parent and bis_bas_reward_parent, we are penalizing with 5 because they are ambiguous.

Let's write the output:

Predictor: currently_taking_lithium | Penalty: 5 | Rationale: Name does not explicitly reference suicidal ideation or self-harm concepts.

Predictor: total_number_of_hospitalizations | Penalty: 5 | Rationale: Administrative count of hospitalizations, not directly related to suicidal ideation.

Predictor: zero_hospitalizations_due_to_mania | Penalty: 5 | Rationale: Administrative indicator of hospitalizations due to mania, not directly related to suicidal ideation.

Predictor: father_education_less_than_12_years | Penalty: 5 | Rationale: Demographic variable (father's education level).

Predictor: obesity | Penalty: 4 | Rationale: Broader clinical construct (comorbidity) in scope of the study.

Predictor: severe_obesity | Penalty: 4 | Rationale: Broader clinical construct (comorbidity) in scope of the study.

Predictor: hispanic_or_latino | Penalty: 5 | Rationale: Demographic variable (ethnicity).

Predictor: bis_bas_bis_parent | Penalty: 5 | Rationale: Ambiguous name not self-explanatory without context.

Predictor: bis_bas_reward_parent | Penalty: 5 | Rationale: Ambiguous name not self-explanatory without context.

Predictor: birthweight_less_than_5.5_pounds | Penalty: 5 | Rationale: Demographic/medical history variable.

We must be careful: the rationale must be one sentence and grounded in the name.

Let's write them as required.

</think>

Predictor: currently_taking_lithium | Penalty: 5 | Rationale: Name does not explicitly reference suicidal ideation or self-harm concepts.

Predictor: total_number_of_hospitalizations | Penalty: 5 | Rationale: Administrative count of hospitalizations, not directly related to suicidal ideation.

Predictor: zero_hospitalizations_due_to_mania | Penalty: 5 | Rationale: Administrative indicator of hospitalizations due to mania, not directly related to suicidal ideation.

Predictor: father_education_less_than_12_years | Penalty: 5 | Rationale: Demographic variable (father's education level).

Predictor: obesity | Penalty: 4 | Rationale: Broader clinical construct (comorbidity) in scope of the study.

Predictor: severe_obesity | Penalty: 4 | Rationale: Broader clinical construct (comorbidity) in scope of the study.

Predictor: hispanic_or_latino | Penalty: 5 | Rationale: Demographic variable (ethnicity).

Predictor: bis_bas_bis_parent | Penalty: 5 | Rationale: Ambiguous name not self-explanatory without context.

Predictor: bis_bas_reward_parent | Penalty: 5 | Rationale: Ambiguous name not self-explanatory without context.

Predictor: birthweight_less_than_5.5_pounds | Penalty: 5 | Rationale: Demographic/medical history variable.

----------------------------------------

**Supplementary Data S3. Candidate predictor list, final EBAL coefficients, leave-one-out coefficient summaries, and selection frequencies**

```
**Supplementary Data S3. Candidate predictor list, final EBAL coefficients, leave-one-out coefficient summaries, and selection frequencies**

{
  "feature_names": [
    "years_of_education",
    "current_mood_depressed",
    "current_mood_euthymic",
    "current_mood_manic",
    "lenght_illness_mood_disorder",
    "comorbidities_generalized_anxiety_disorder",
    "comorbidities_personality_disorder",
    "comorbidities_agoraphobia",
    "comorbidities_adhd",
    "currently_taking_antidepressant",
    "currently_taking_lithium",
    "total_number_of_hospitalizations",
    "zero_hospitalizations_due_to_mania",
    "father_education_less_than_12_years",
    "obesity",
    "severe_obesity",
    "hispanic_or_latino",
    "bis_bas_bis_parent",
    "bis_bas_reward_parent",
    "birthweight_less_than_5.5_pounds"
  ],
  "final_model": {
    "coefficients": {
      "years_of_education": 0.0,
      "current_mood_depressed": 0.64752467499242,
      "current_mood_euthymic": -0.11553325279769112,
      "current_mood_manic": -0.17786302657927702,
      "lenght_illness_mood_disorder": 0.5822606217158244,
      "comorbidities_generalized_anxiety_disorder": 0.5105040896143158,
      "comorbidities_personality_disorder": 0.15569724128676862,
      "comorbidities_agoraphobia": 0.0,
      "comorbidities_adhd": -0.0822110996438035,
      "currently_taking_antidepressant": 0.0,
      "currently_taking_lithium": 0.0,
      "total_number_of_hospitalizations": 0.0,
      "zero_hospitalizations_due_to_mania": 0.27509997717228163,
      "father_education_less_than_12_years": 0.0,
      "obesity": 0.21406138873232253,
      "severe_obesity": 0.0,
      "hispanic_or_latino": 0.0,
      "bis_bas_bis_parent": 0.21239072438724269,
      "bis_bas_reward_parent": -0.05732696034507967,
      "birthweight_less_than_5.5_pounds": 0.0
    },
    "intercept": 0.48887418672307886,
    "n_nonzero": 11
  },
  "loo_aggregated": {
    "mean_coefficients": {
      "years_of_education": 0.020760884154003765,
      "current_mood_depressed": 0.9721835611812497,
      "current_mood_euthymic": -0.06467396251035631,
      "current_mood_manic": -0.2171066582460771,
      "lenght_illness_mood_disorder": 0.9243997773097226,
      "comorbidities_generalized_anxiety_disorder": 0.6411831996358998,
      "comorbidities_personality_disorder": 0.28012078903494914,
      "comorbidities_agoraphobia": 0.2711907192539787,
      "comorbidities_adhd": -0.28427631330620573,
      "currently_taking_antidepressant": -0.03646948940040371,
      "currently_taking_lithium": 0.0036249321058805972,
      "total_number_of_hospitalizations": -0.031973389387111265,
      "zero_hospitalizations_due_to_mania": 0.38025543012648155,
      "father_education_less_than_12_years": -0.08795180861958617,
      "obesity": 0.35053784382290354,
      "severe_obesity": 0.12493438040560155,
      "hispanic_or_latino": 0.13013250721846265,
      "bis_bas_bis_parent": 0.49188604589641044,
      "bis_bas_reward_parent": -0.47135020468407374,
      "birthweight_less_than_5.5_pounds": 0.16333218810047026
    },
    "std_coefficients": {
      "years_of_education": 0.054019050593347676,
      "current_mood_depressed": 0.16901326100063158,
      "current_mood_euthymic": 0.09066999832490205,
      "current_mood_manic": 0.0865403006511445,
      "lenght_illness_mood_disorder": 0.2108059872669199,
      "comorbidities_generalized_anxiety_disorder": 0.117083515261159,
      "comorbidities_personality_disorder": 0.15471800603589642,
      "comorbidities_agoraphobia": 0.1391366120470717,
      "comorbidities_adhd": 0.11136498295022855,
      "currently_taking_antidepressant": 0.0632502910309045,
      "currently_taking_lithium": 0.018490928841778036,
      "total_number_of_hospitalizations": 0.06791635476415954,
      "zero_hospitalizations_due_to_mania": 0.14259374767727262,
      "father_education_less_than_12_years": 0.10894201823990131,
      "obesity": 0.1335016038974101,
      "severe_obesity": 0.09251842800029568,
      "hispanic_or_latino": 0.11550148121393986,
      "bis_bas_bis_parent": 0.1977187697346742,
      "bis_bas_reward_parent": 0.24808738930046678,
      "birthweight_less_than_5.5_pounds": 0.12731217920938168
    },
    "selection_frequency": {
      "years_of_education": 0.3161764705882353,
      "current_mood_depressed": 1.0,
      "current_mood_euthymic": 0.5220588235294118,
      "current_mood_manic": 1.0,
      "lenght_illness_mood_disorder": 1.0,
      "comorbidities_generalized_anxiety_disorder": 1.0,
      "comorbidities_personality_disorder": 0.9705882352941176,
      "comorbidities_agoraphobia": 0.9705882352941176,
      "comorbidities_adhd": 0.9852941176470589,
      "currently_taking_antidepressant": 0.4117647058823529,
      "currently_taking_lithium": 0.14705882352941177,
      "total_number_of_hospitalizations": 0.2867647058823529,
      "zero_hospitalizations_due_to_mania": 1.0,
      "father_education_less_than_12_years": 0.5882352941176471,
      "obesity": 1.0,
      "severe_obesity": 0.8455882352941176,
      "hispanic_or_latino": 0.7720588235294118,
      "bis_bas_bis_parent": 1.0,
      "bis_bas_reward_parent": 0.9485294117647058,
      "birthweight_less_than_5.5_pounds": 0.8235294117647058
    },
    "mean_intercept": 0.9332623546960878,
    "std_intercept": 0.253964953934502
  }
}
```

**Supplementary Data S4. Per-subject leave-one-out predicted probabilities for suicidal ideation (MINI-KID item B1a, N = 136).**

*Subjects are numbered 1 to 136. Original label is the B1a outcome (1 = reported suicidal ideation, 0 = no report). Predicted probabilities are from nested leave-one-out cross-validation; classifications use a 0.5 threshold. Values reproduce the reported metrics exactly for both models. EBAL: AUROC 0.768, balanced accuracy 0.757, sensitivity 0.758, specificity 0.757. Standard LASSO: AUROC 0.760, balanced accuracy 0.715, sensitivity 0.727, specificity 0.703.*

| **Subject** | **Original label (B1a)** | **EBAL predicted probability** | **Standard LASSO predicted probability** |
| --- | --- | --- | --- |
| 1 | 1 | 0.924656 | 0.944198 |
| 2 | 1 | 0.480752 | 0.457447 |
| 3 | 0 | 0.191836 | 0.187603 |
| 4 | 1 | 0.609072 | 0.606418 |
| 5 | 0 | 0.033596 | 0.028525 |
| 6 | 1 | 0.788667 | 0.819149 |
| 7 | 0 | 0.151540 | 0.123654 |
| 8 | 1 | 0.943584 | 0.950860 |
| 9 | 1 | 0.164026 | 0.161678 |
| 10 | 1 | 0.998090 | 0.998965 |
| 11 | 1 | 0.269315 | 0.253904 |
| 12 | 1 | 0.981875 | 0.974147 |
| 13 | 1 | 0.379117 | 0.379259 |
| 14 | 0 | 0.641009 | 0.624104 |
| 15 | 0 | 0.338519 | 0.468784 |
| 16 | 0 | 0.398770 | 0.509127 |
| 17 | 1 | 0.998466 | 0.998949 |
| 18 | 1 | 0.511388 | 0.486362 |
| 19 | 0 | 0.222865 | 0.153366 |
| 20 | 1 | 0.564098 | 0.446755 |
| 21 | 1 | 0.830374 | 0.793730 |
| 22 | 1 | 0.582030 | 0.685241 |
| 23 | 1 | 0.540921 | 0.242844 |
| 24 | 0 | 0.359462 | 0.524434 |
| 25 | 1 | 0.940041 | 0.963176 |
| 26 | 1 | 0.672636 | 0.641865 |
| 27 | 0 | 0.162515 | 0.144265 |
| 28 | 1 | 0.504789 | 0.543543 |
| 29 | 1 | 0.922836 | 0.848590 |
| 30 | 1 | 0.999083 | 0.999302 |
| 31 | 1 | 0.271046 | 0.346661 |
| 32 | 0 | 0.291971 | 0.267041 |
| 33 | 0 | 0.017766 | 0.015432 |
| 34 | 1 | 0.481866 | 0.444084 |
| 35 | 1 | 0.611006 | 0.636224 |
| 36 | 1 | 0.967965 | 0.989754 |
| 37 | 1 | 0.608976 | 0.618051 |
| 38 | 1 | 0.908118 | 0.916725 |
| 39 | 1 | 0.999568 | 0.999723 |
| 40 | 1 | 0.995760 | 0.996582 |
| 41 | 1 | 0.940778 | 0.941878 |
| 42 | 1 | 0.512340 | 0.526175 |
| 43 | 1 | 0.857724 | 0.928342 |
| 44 | 0 | 0.855637 | 0.859138 |
| 45 | 1 | 0.437477 | 0.442689 |
| 46 | 1 | 0.078724 | 0.089268 |
| 47 | 0 | 0.245653 | 0.183176 |
| 48 | 0 | 0.294410 | 0.289800 |
| 49 | 1 | 0.464140 | 0.439278 |
| 50 | 1 | 0.633953 | 0.622989 |
| 51 | 1 | 0.137638 | 0.121872 |
| 52 | 1 | 0.884274 | 0.896066 |
| 53 | 1 | 0.979253 | 0.987786 |
| 54 | 0 | 0.161912 | 0.200427 |
| 55 | 1 | 0.565677 | 0.587644 |
| 56 | 1 | 0.998415 | 0.998017 |
| 57 | 1 | 0.844426 | 0.875943 |
| 58 | 1 | 0.767509 | 0.827846 |
| 59 | 1 | 0.509615 | 0.508821 |
| 60 | 1 | 0.979912 | 0.986556 |
| 61 | 1 | 0.937001 | 0.959062 |
| 62 | 1 | 0.675303 | 0.696230 |
| 63 | 1 | 0.082488 | 0.074548 |
| 64 | 0 | 0.680161 | 0.698709 |
| 65 | 1 | 0.993241 | 0.995657 |
| 66 | 0 | 0.315381 | 0.378296 |
| 67 | 1 | 0.516451 | 0.548426 |
| 68 | 1 | 0.966647 | 0.973949 |
| 69 | 1 | 0.851444 | 0.922355 |
| 70 | 1 | 0.734815 | 0.730643 |
| 71 | 1 | 0.418009 | 0.472679 |
| 72 | 1 | 0.877027 | 0.977442 |
| 73 | 1 | 0.532579 | 0.750039 |
| 74 | 1 | 0.907841 | 0.875034 |
| 75 | 1 | 0.915537 | 0.913702 |
| 76 | 1 | 0.308409 | 0.327044 |
| 77 | 1 | 0.934400 | 0.947549 |
| 78 | 1 | 0.999144 | 0.999064 |
| 79 | 1 | 0.707432 | 0.705661 |
| 80 | 0 | 0.098153 | 0.102741 |
| 81 | 0 | 0.205265 | 0.183807 |
| 82 | 0 | 0.146635 | 0.195161 |
| 83 | 1 | 0.875212 | 0.904345 |
| 84 | 1 | 0.902447 | 0.884580 |
| 85 | 1 | 0.182916 | 0.193809 |
| 86 | 0 | 0.446139 | 0.441306 |
| 87 | 0 | 0.703022 | 0.671200 |
| 88 | 1 | 0.319500 | 0.319393 |
| 89 | 0 | 0.810283 | 0.827427 |
| 90 | 1 | 0.157408 | 0.130154 |
| 91 | 1 | 0.953074 | 0.990652 |
| 92 | 0 | 0.169795 | 0.158916 |
| 93 | 1 | 0.765178 | 0.758778 |
| 94 | 1 | 0.794615 | 0.774775 |
| 95 | 1 | 0.971817 | 0.955622 |
| 96 | 1 | 0.859887 | 0.874116 |
| 97 | 1 | 0.996050 | 0.996736 |
| 98 | 1 | 0.995868 | 0.996882 |
| 99 | 1 | 0.775105 | 0.659181 |
| 100 | 1 | 0.846170 | 0.857177 |
| 101 | 1 | 0.103919 | 0.103050 |
| 102 | 1 | 0.804204 | 0.839198 |
| 103 | 0 | 0.683045 | 0.685252 |
| 104 | 1 | 0.637491 | 0.648542 |
| 105 | 1 | 0.929470 | 0.942652 |
| 106 | 1 | 0.208327 | 0.148575 |
| 107 | 1 | 0.983888 | 0.988243 |
| 108 | 1 | 0.748934 | 0.821773 |
| 109 | 1 | 0.825240 | 0.828300 |
| 110 | 0 | 0.478678 | 0.476459 |
| 111 | 1 | 0.830973 | 0.812340 |
| 112 | 0 | 0.060761 | 0.044646 |
| 113 | 1 | 0.583817 | 0.568817 |
| 114 | 1 | 0.429139 | 0.457970 |
| 115 | 0 | 0.435765 | 0.401411 |
| 116 | 1 | 0.303875 | 0.218532 |
| 117 | 1 | 0.173710 | 0.169129 |
| 118 | 0 | 0.422646 | 0.435948 |
| 119 | 0 | 0.982274 | 0.970630 |
| 120 | 0 | 0.276800 | 0.282042 |
| 121 | 0 | 0.995912 | 0.996406 |
| 122 | 0 | 0.394223 | 0.390179 |
| 123 | 1 | 0.259590 | 0.259631 |
| 124 | 0 | 0.104297 | 0.094094 |
| 125 | 0 | 0.979826 | 0.996940 |
| 126 | 1 | 0.957185 | 0.956619 |
| 127 | 1 | 0.995123 | 0.993930 |
| 128 | 1 | 0.337346 | 0.341994 |
| 129 | 1 | 0.739179 | 0.734514 |
| 130 | 1 | 0.790777 | 0.808020 |
| 131 | 1 | 0.034748 | 0.036693 |
| 132 | 0 | 0.038776 | 0.020535 |
| 133 | 0 | 0.348506 | 0.370284 |
| 134 | 1 | 0.735148 | 0.719661 |
| 135 | 1 | 0.616077 | 0.710728 |
| 136 | 1 | 0.821262 | 0.818459 |
